# Detailed stratified GWAS analysis for severe COVID-19 in four European populations

**DOI:** 10.1101/2021.07.21.21260624

**Authors:** Frauke Degenhardt, David Ellinghaus, Simonas Juzenas, Jon Lerga-Jaso, Mareike Wendorff, Douglas Maya-Miles, Florian Uellendahl-Werth, Hesham ElAbd, Malte C Rühlemann, Jatin Arora, Onur Özer, Ole Bernt Lenning, Ronny Myhre, May Sissel Vadla, Eike M Wacker, Lars Wienbrandt, Aaron Blandino Ortiz, Adolfo de Salazar, Adolfo Garrido Chercoles, Adriana Palom, Agustín Ruiz, Alba-Estela Garcia-Fernandez, Albert Blanco-Grau, Alberto Mantovani, Alberto Zanella, Aleksander Rygh Holten, Alena Mayer, Alessandra Bandera, Alessandro Cherubini, Alessandro Protti, Alessio Aghemo, Alessio Gerussi, Alfredo Ramirez, Alice Braun, Almut Nebel, Ana Barreira, Ana Lleo, Ana Teles, Anders Benjamin Kildal, Andrea Biondi, Andrea Caballero-Garralda, Andrea Ganna, Andrea Gori, Andreas Glück, Andreas Lind, Anja Tanck, Anke Hinney, Anna Carreras Nolla, Anna Ludovica Fracanzani, Anna Peschuck, Annalisa Cavallero, Anne Ma Dyrhol-Riise, Antonella Ruello, Antonio Julià, Antonio Muscatello, Antonio Pesenti, Antonio Voza, Ariadna Rando-Segura, Aurora Solier, Axel Schmidt, Beatriz Cortes, Beatriz Mateos, Beatriz Nafria-Jimenez, Benedikt Schaefer, Björn Jensen, Carla Bellinghausen, Carlo Maj, Carlos Ferrando, Carmen de la Horra, Carmen Quereda, Carsten Skurk, Charlotte Thibeault, Chiara Scollo, Christian Herr, Christoph D Spinner, Christoph Gassner, Christoph Lange, Cinzia Hu, Cinzia Paccapelo, Clara Lehmann, Claudio Angelini, Claudio Cappadona, Clinton Azuure, COVICAT study group, Covid-19 Aachen Study (COVAS), Cristiana Bianco, Cristina Cea, Cristina Sancho, Dag Arne Lihaug Hoff, Daniela Galimberti, Daniele Prati, David Haschka, David Jiménez, David Pestaña, David Toapanta, Eduardo Muñiz-Diaz, Elena Azzolini, Elena Sandoval, Eleonora Binatti, Elio Scarpini, Elisa T Helbig, Elisabetta Casalone, Eloisa Urrechaga, Elvezia Maria Paraboschi, Emanuele Pontali, Enric Reverter, Enrique J Calderón, Enrique Navas, Erik Solligård, Ernesto Contro, Eunate Arana-Arri, Fátima Aziz, Federico Garcia, Félix García Sánchez, Ferruccio Ceriotti, Filippo Martinelli-Boneschi, Flora Peyvandi, Florian Kurth, Francesco Blasi, Francesco Malvestiti, Francisco J Medrano, Francisco Mesonero, Francisco Rodriguez-Frias, Frank Hanses, Fredrik Müller, Georg Hemmrich-Stanisak, Giacomo Bellani, Giacomo Grasselli, Gianni Pezzoli, Giorgio Costantino, Giovanni Albano, Giulia Cardamone, Giuseppe Bellelli, Giuseppe Citerio, Giuseppe Foti, Giuseppe Lamorte, Giuseppe Matullo, Guido Baselli, Hayato Kurihara, Holger Neb, Ilaria My, Ingo Kurth, Isabel Hernández, Isabell Pink, Itziar de Rojas, Iván Galván-Femenia, Jan Cato Holter, Jan Egil Afset, Jan Heyckendorf, Jan Kässens, Jan Kristian Damås, Jan Rybniker, Janine Altmüller, Javier Ampuero, Javier Martín, Jeanette Erdmann, Jesus M Banales, Joan Ramon Badia, Joaquin Dopazo, Jochen Schneider, Jonas Bergan, Jordi Barretina, Jörn Walter, Jose Hernández Quero, Josune Goikoetxea, Juan Delgado, Juan M Guerrero, Julia Fazaal, Julia Kraft, Julia Schröder, Kari Risnes, Karina Banasik, Karl Erik Müller, Karoline I Gaede, Koldo Garcia-Etxebarria, Kristian Tonby, Lars Heggelund, Laura Izquierdo-Sanchez, Laura Rachele Bettini, Lauro Sumoy, Leif Erik Sander, Lena J Lippert, Leonardo Terranova, Lindokuhle Nkambule, Lisa Knopp, Lise Tuset Gustad, Lucia Garbarino, Luigi Santoro, Luis Téllez, Luisa Roade, Mahnoosh Ostadreza, Maider Intxausti, Manolis Kogevinas, Mar Riveiro-Barciela, Marc M Berger, Marco Schaefer, Mari EK Niemi, María A Gutiérrez-Stampa, Maria Carrabba, Maria E. Figuera Basso, Maria Grazia Valsecchi, María Hernandez-Tejero, Maria JGT Vehreschild, Maria Manunta, Marialbert Acosta-Herrera, Mariella D’Angiò, Marina Baldini, Marina Cazzaniga, Marit M Grimsrud, Markus Cornberg, Markus M Nöthen, Marta Marquié, Massimo Castoldi, Mattia Cordioli, Maurizio Cecconi, Mauro D’Amato, Max Augustin, Melissa Tomasi, Mercè Boada, Michael Dreher, Michael J Seilmaier, Michael Joannidis, Michael Wittig, Michela Mazzocco, Michele Ciccarelli, Miguel Rodríguez-Gandía, Monica Bocciolone, Monica Miozzo, Natale Imaz Ayo, Natalia Blay, Natalia Chueca, Nicola Montano, Nicole Braun, Nicole Ludwig, Nikolaus Marx, Nilda Martínez, Norwegian SARS-CoV-2 Study group, Oliver A Cornely, Oliver Witzke, Orazio Palmieri, Pa COVID-19 Study Group, Paola Faverio, Paoletta Preatoni, Paolo Bonfanti, Paolo Omodei, Paolo Tentorio, Pedro Castro, Pedro M Rodrigues, Pedro Pablo España, Per Hoffmann, Philip Rosenstiel, Philipp Schommers, Phillip Suwalski, Raúl de Pablo, Ricard Ferrer, Robert Bals, Roberta Gualtierotti, Rocío Gallego-Durán, Rosa Nieto, Rossana Carpani, Rubén Morilla, Salvatore Badalamenti, Sammra Haider, Sandra Ciesek, Sandra May, Sara Bombace, Sara Marsal, Sara Pigazzini, Sebastian Klein, Serena Pelusi, Sibylle Wilfling, Silvano Bosari, Sonja Volland, Søren Brunak, Soumya Raychaudhuri, Stefan Schreiber, Stefanie Heilmann-Heimbach, Stefano Aliberti, Stephan Ripke, Susanne Dudman, Tanja Wesse, Tenghao Zheng, The Humanitas COVID-19 Task Force, The Humanitas Gavazzeni COVID-19 Task Force, Thomas Bahmer, Thomas Eggermann, Thomas Illig, Thorsten Brenner, Tomas Pumarola, Torsten Feldt, Trine Folseraas, Trinidad Gonzalez Cejudo, Ulf Landmesser, Ulrike Protzer, Ute Hehr, Valeria Rimoldi, Valter Monzani, Vegard Skogen, Verena Keitel, Verena Kopfnagel, Vicente Friaza, Victor Andrade, Victor Moreno, Wolfgang Albrecht, Wolfgang Peter, Wolfgang Poller, Xavier Farre, Xiaoli Yi, Xiaomin Wang, Yascha Khodamoradi, Zehra Karadeniz, Anna Latiano, Siegfried Goerg, Petra Bacher, Philipp Koehler, Florian Tran, Heinz Zoller, Eva C Schulte, Bettina Heidecker, Kerstin U Ludwig, Javier Fernández, Manuel Romero-Gómez, Agustín Albillos, Pietro Invernizzi, Maria Buti, Stefano Duga, Luis Bujanda, Johannes R Hov, Tobias L Lenz, Rosanna Asselta, Rafael de Cid, Luca Valenti, Tom H Karlsen, Mario Cáceres, Andre Franke

**Author notes:** **[CORRESPONDING AUTHORS]** Dr. Frauke Degenhardt, Address: Institute of Clinical Molecular Biology and University Hospital of Schleswig-Holstein, Christian-Albrechts-University, Rosalind-Franklin-Str. 12, D-24105 Kiel, Germany, Mail; Prof. Dr. Andre Franke, Address: Institute of Clinical Molecular Biology and University Hospital of Schleswig-Holstein, Christian-Albrechts-University, Rosalind-Franklin-Str. 12, D-24105 Kiel, Germany, Mail. The authors wish it to be known that, in their opinion, the first 6 authors should be regarded as joint First Authors. The authors wish it to be known that, in their opinion, the last 7 authors should be regarded as joint Last Authors.

## Abstract

Given the highly variable clinical phenotype of Coronavirus disease 2019 (COVID-19), a deeper analysis of the host genetic contribution to severe COVID-19 is important to improve our understanding of underlying disease mechanisms. Here, we describe an extended GWAS meta-analysis of a well-characterized cohort of 3,260 COVID-19 patients with respiratory failure and 12,483 population controls from Italy, Spain, Norway and Germany/Austria, including stratified analyses based on age, sex and disease severity, as well as targeted analyses of chromosome Y haplotypes, the human leukocyte antigen (HLA) region and the SARS-CoV-2 peptidome. By inversion imputation, we traced a reported association at 17q21.31 to a highly pleiotropic ∼0.9-Mb inversion polymorphism and characterized the potential effects of the inversion in detail. Our data, together with the 5^th^ release of summary statistics from the COVID-19 Host Genetics Initiative, also identified a new locus at 19q13.33, including *NAPSA*, a gene which is expressed primarily in alveolar cells responsible for gas exchange in the lung.

## Introduction

In the past year, Coronavirus disease 2019 (COVID-19), caused by the severe acute respiratory syndrome coronavirus 2 (SARS-CoV-2), has evolved into a global pandemic with more than 312 million confirmed cases and 5.5 million COVID-19 related deaths worldwide (frequencies reported by the World Health Organization, January 12th, 2022). The clinical manifestations of COVID-19 are variable and range from complete absence of symptoms to severe respiratory failure and death. Severe COVID-19 requires intensive medical care with respiratory support and can result in long-term damages detrimental to the individual. The pathogenesis of severe COVID-19 is, however, still poorly understood. This condition has been associated with clinical risk factors such as old age, male sex and comorbidities including diabetes, active cancer, hypertension and coronary artery disease as well as solid organ transplant or other conditions that promote an immunosuppressive state.(1–4)

Different studies, including the first genetic analysis of our Severe COVID-19 GWAS study group, have shown that genetic predisposition plays a role in COVID-19 susceptibility and severity.(5–7) In particular, we reported significant associations between genetic variants at loci 3p21.31 (around *LZTF1*) and 9q34.2 (*ABO* blood group locus) to severe respiratory failure and SARS-CoV-2 infection in 1,980 severe COVID-19 patients and 2,160 population controls of European ancestry. Since then, these loci have been replicated in subsequent studies and extended also to non-European cohorts.(8–10) 11 additional genome-wide significant loci, associated with SARS-CoV-2 infection or COVID-19 manifestations have been reported by various studies, including the Genetics Of Mortality In Critical Care (GenOMICC) Initiative and, more recently, the largest genetic consortium for COVID-19, the COVID-19 Host Genetics initiative (HGI), which contains 3,815 individuals from the Severe COVID-19 GWAS study group.(6,7) Six of these loci have been linked to critical illness by COVID-19, and include genes that were previously associated with pulmonary or autoimmune and inflammatory diseases.(7)

Since our primary publication, the Severe COVID-19 GWAS study group dataset has been extended to 3,260 severe COVID-19 patients with respiratory failure(5) and 12,483 controls with unknown COVID-19 status from 30 study centers across Italy, Spain, Norway and Germany/Austria (**Supplementary Table 1a**). Further, detailed information on age, sex and comorbidities as hypertension, diabetes, and coronary artery disease is available in this dataset. The control cohort included predominantly population controls recruited at the respective centers as blood or bone marrow donors with unknown or negative COVID-19 status (**Supplementary Table 1c**). With this unique resource at hand, we performed an extended genome-wide association study (GWAS) and meta-analysis of severe COVID-19 as well as a meta-analysis with the COVID-19 HGI release 5 GWAS summary statistics, followed by stratified analyses of age, sex and disease and a detailed analysis of ABO secretor status. Using Y-chromosomal genotype calls from the Global Screening Array (GSA), we additionally performed an imputation and disease association of chromosome Y haplotypes. Compared to our first study, we also performed an even more detailed analysis of the HLA which includes classical fine-mapping of the HLA region based on local imputation of SNP, amino acid and classical allele information, as well as a broad range of other approaches, including a peptidome-wide association study (PepWAS)(11) computational prediction of SARS-CoV-2 peptide presentation, HLA class I supertype association analysis, and tests for heterozygote advantage, divergent allele advantage and molecular mimicry. Finally, by inversion imputation, we present a functional analysis of the established 17q21.31 locus(7), which has been described by COVID-19 HGI but not characterized in depth.

## Results

### GWAS meta-analyses for severe COVID-19 with respiratory failure

Genotyping was carried out using Illumina’s GSA, followed by genotype quality control (QC) analysis and TOPMed genotype imputation (**Online Methods, Supplementary Figure 1**, patient numbers before and after QC are shown in **Supplementary Table 1**). As before, respiratory failure was defined as respiratory support with supplemental oxygen [class 1] or non-invasive and invasive ventilation [classes 2 and 3, respectively], or by extracorporeal membrane oxygenation (ECMO) [class 4]).^5^ Analogously to the COVID-19 HGI(7), we conducted two GWAS discovery meta-analyses for two main categories of COVID-19 disease state: First, “hospitalization with respiratory support” (respiratory support classes 1-4 with a total of 3,260 patients and 12,483 controls; first analysis) and second, a more stringent definition of severe COVID-19 “hospitalization with mechanical ventilation” (classes 2-4 with 1,911 critically ill individuals and 12,226 controls; second analysis). Compared to our previous study, we increased the number of patients and controls by approximate factors of 2 and 5.5 respectively (from 1,610 and 2,205). Details of per cohort patient numbers are shown in **Supplementary Table 1e**. The characteristics of patients and controls included in these analyses are shown in **Table 1** and **Supplementary Table 2**. After imputation, we carried out a GWAS of 9,223,806 high-quality genetic variants (imputation R^2^≥0.6 and minor allele frequency (MAF) ≥1%) stratified by ancestry (Italy, Spain, Norway and Germany/Austria) using logistic mixed model analysis as implemented in SAIGE(12), followed by fixed-effect inverse variance meta-analysis using METAL(13) (low genomic inflation of 1.017; **Supplementary Figures 2-3**). Genome-wide comparison of the case-control frequencies (conservatively adjusted for age, sex, age*age, sex*age and top 10 principal components from principal component analysis (PCA) as employed by the COVID-19 HGI; **Online Methods**) revealed genome-wide significant associations (P<5×10^−8^; **Supplementary Table 3**) at four known loci: chr3:45848457:C:T (rs35731912) within *LZTFL1* at the 3p21.31 locus, chr9:133261703:A:G (rs687289) within *ABO* at 9q34.2, chr19:10351837:C:T (rs687289) within *TYK2* at 19p13.2 and chr19:4717660:A:G (rs12610495) within *DPP9* at 19p13.3 in the first or second analysis. Variants at four loci (*PCDH7* at 4p15.1; *FREM1* at 9p22.3; *MAPT* at 17q21.31; *DPP9* at 19p13.3) had suggestive significance (*P*<10^−6^) in our first analysis, and an additional five loci (*OLMF4* at 13q21.1; *TTC7B* at 14q32.11; *CPD* at 17q11.2; *PTPRM* at 18p11.23; *IFNAR2* at 21q22.11) had suggestive significance in the second analysis (**Figure 1**; **Supplementary Table 3**, regional association plots shown in **Supplementary Figures 4-5**). 3 loci not included in the list above had evidence for excessive heterogeneity (P <10^−5^). Of note, associations at 17q21.31 (*MAPT*), 19p13.3 (*DPP9*) and 21q22.11 (*IFNAR2*) have been previously reported by the COVID-19 HGI.(7) The respective lead risk variants (this study vs. COVID-19 HGI) at reported loci are linked with r^2^ or D^’^ >0.9 across our study cohorts (**Supplementary Table 4**). Hierarchical mixture model analysis with MAMBA(14) (**Online Methods**) showed a high posterior probability of replication (PPR>0.8) of consistent effect sizes across all analyzed cohorts for all but chr14:90679345:C:T at 14q32.11 (*TTC7B*, PPR=0.005) and chr17:30409086:A:T at 17q11.2 (CPD, PPR=0.038) (**Supplementary Table 3**). The chr17:45933112:G:A *MAPT* variant (rs8065800), showed higher replicability in our first analysis compared to the second analysis (*MAPT*, PPR=0.93/PPR=0.2). None of the novel and replicable associations at *PCDH7* at 4p15.1; *FREM1* at 9p22.3; *OLMF4* at 13q21.1 and *PTPRM* at 18p11.23 had suggestive associations in the COVID-19 HGI 5 release dataset, which is why we did not consider them further. Future analyses will explore whether these are attributable to artefacts in the cohorts of this study or maybe a too high heterogeneity of cohorts and data sets included in the COVID-19 HGI analyses.

**Table 1.**
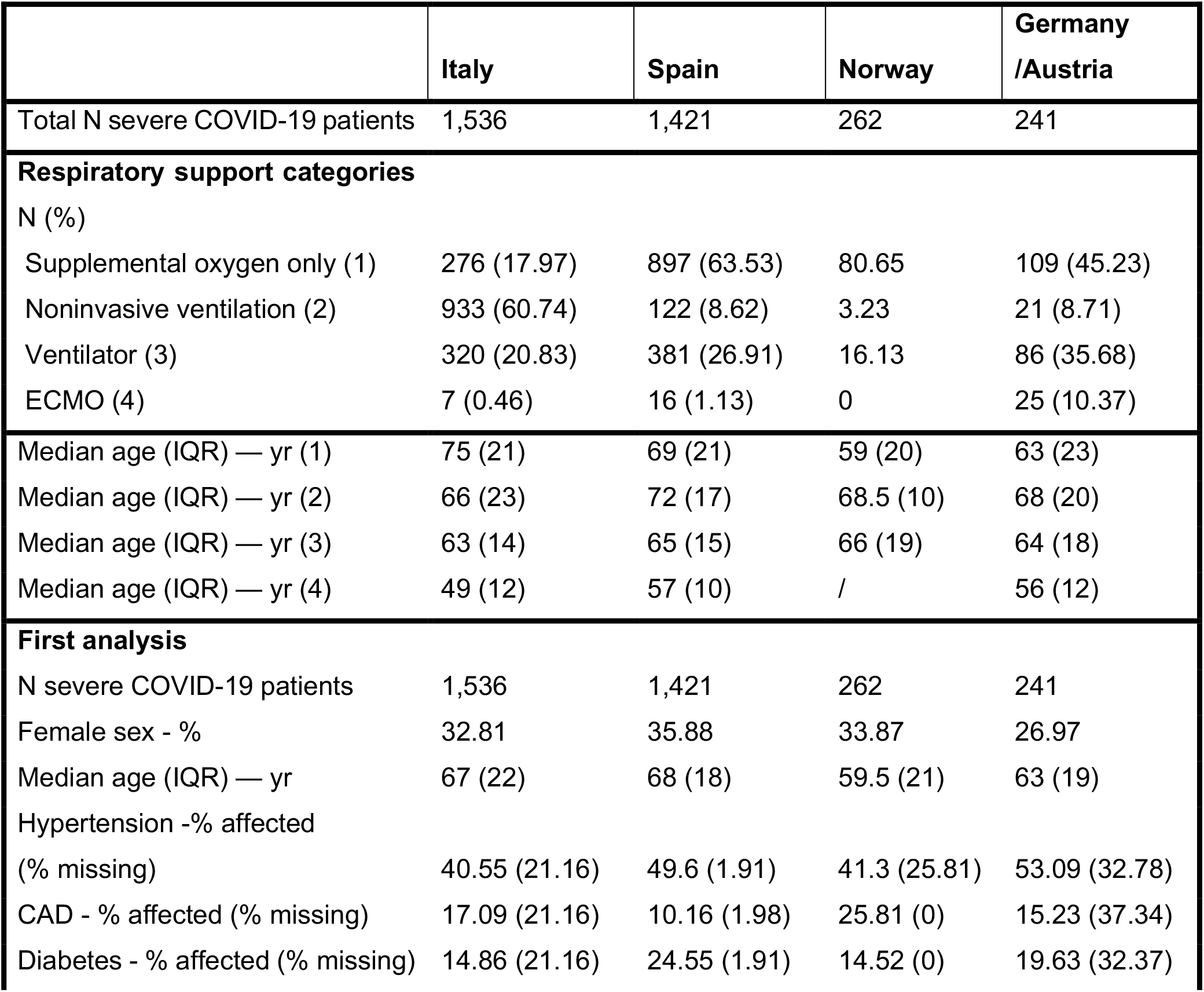
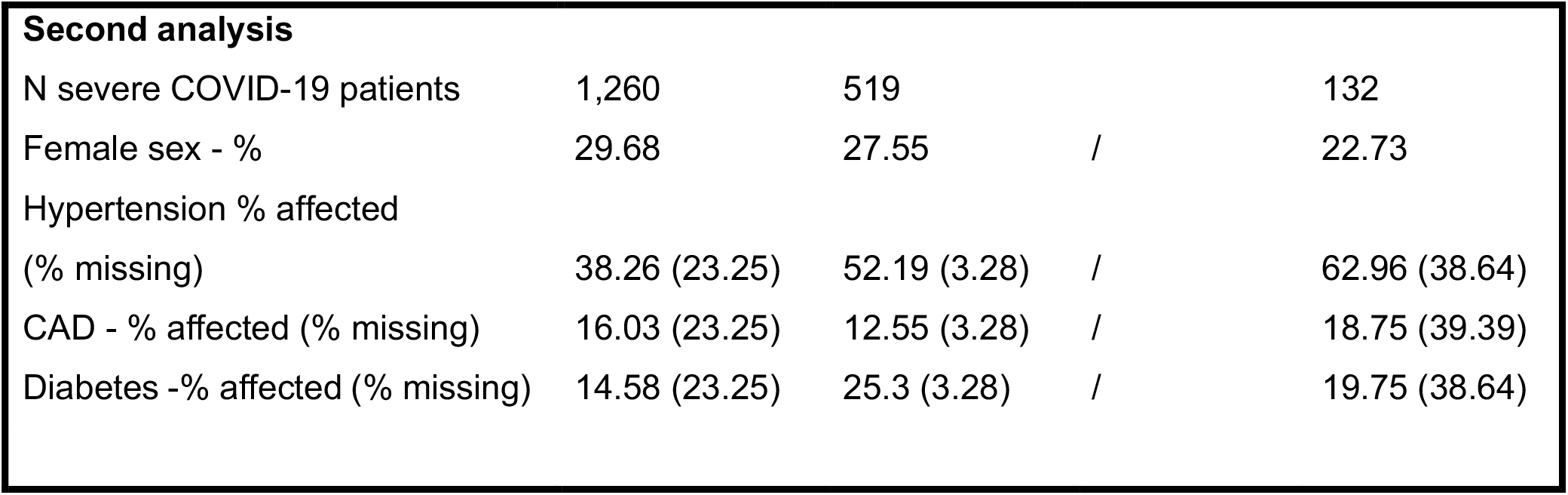
Overview of patients included in the genome-wide discovery analysis. Overview of patients included in our first analysis (3,260 patients) and second analysis (1,911 patients). Individuals of the Italian, Spanish, Norwegian and German/Austrian cohorts were recruited at five, seven, eight and ten different hospitals/centers, respectively. Shown are respiratory support status groups 1-4, age and median age across all individuals as well as within each respiratory support group, percentage of females within each cohort, as well as percentage of individuals affected by known comorbidities of COVID-19. Commonly reported comorbidities in COVID-19 are shown, hypertension, coronary artery disease (CAD) and diabetes. Characteristics of control individuals are shown in **Supplementary Table 2**.

|  | Italy | Spain | Norway | Germany /Austria |
| --- | --- | --- | --- | --- |
| Total N severe COVID-19 patients | 1,536 | 1,421 | 262 | 241 |
| **Respiratory support categories** |  |  |  |  |
| N (%) |  |  |  |  |
| Supplemental oxygen only (1) | 276 (17.97) | 897 (63.53) | 80.65 | 109 (45.23) |
| Noninvasive ventilation (2) | 933 (60.74) | 122 (8.62) | 3.23 | 21 (8.71) |
| Ventilator (3) | 320 (20.83) | 381 (26.91) | 16.13 | 86 (35.68) |
| ECMO (4) | 7 (0.46) | 16 (1.13) | 0 | 25 (10.37) |
| Median age (IQR) — yr (1) | 75 (21) | 69 (21) | 59 (20) | 63 (23) |
| Median age (IQR) — yr (2) | 66 (23) | 72 (17) | 68.5 (10) | 68 (20) |
| Median age (IQR) — yr (3) | 63 (14) | 65 (15) | 66 (19) | 64 (18) |
| Median age (IQR) — yr (4) | 49 (12) | 57 (10) | / | 56 (12) |
| **First analysis** |  |  |  |  |
| N severe COVID-19 patients | 1,536 | 1,421 | 262 | 241 |
| Female sex - % | 32.81 | 35.88 | 33.87 | 26.97 |
| Median age (IQR) — yr | 67 (22) | 68 (18) | 59.5 (21) | 63 (19) |
| Hypertension -% affected |  |  |  |  |
| (% missing) | 40.55 (21.16) | 49.6 (1.91) | 41.3 (25.81) | 53.09 (32.78) |
| CAD - % affected (% missing) | 17.09 (21.16) | 10.16 (1.98) | 25.81 (0) | 15.23 (37.34) |
| Diabetes - % affected (% missing) | 14.86 (21.16) | 24.55 (1.91) | 14.52 (0) | 19.63 (32.37) |
| **Second analysis** |  |  |  |  |
| N severe COVID-19 patients | 1,260 | 519 |  | 132 |
| Female sex - % | 29.68 | 27.55 | / | 22.73 |
| Hypertension % affected |  |  |  |  |
| (% missing) | 38.26 (23.25) | 52.19 (3.28) | / | 62.96 (38.64) |
| CAD - % affected (% missing) | 16.03 (23.25) | 12.55 (3.28) | / | 18.75 (39.39) |
| Diabetes -% affected (% missing) | 14.58 (23.25) | 25.3 (3.28) | / | 19.75 (38.64) |

**Figure 1.**
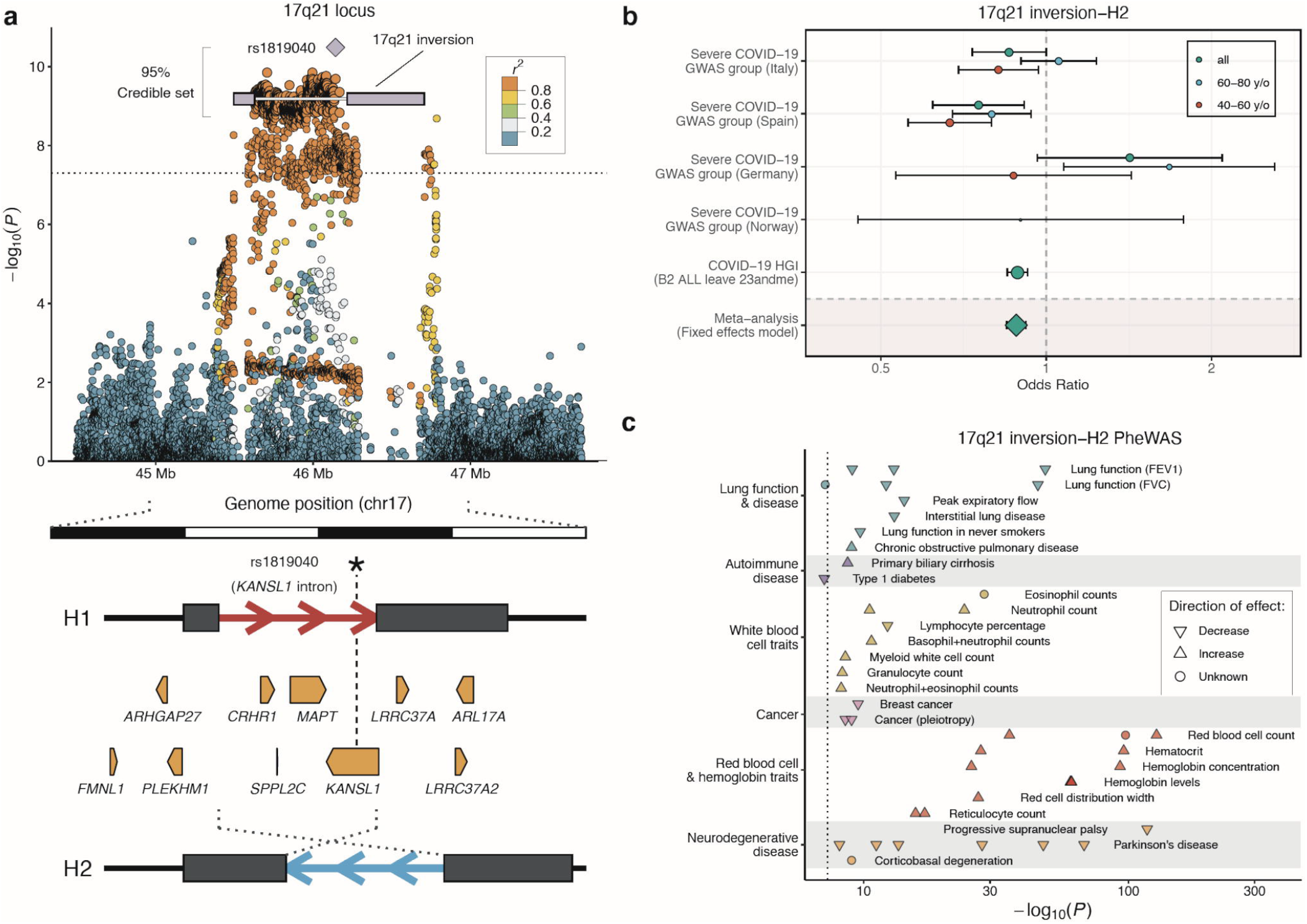
Association of the 17q21.31 locus with severe COVID-19 with respiratory failure. **(a)** Regional association plot showing the variant most strongly associated with severe COVID-19 (rs1819040, purple diamond) a ∼0.9-Mb inversion polymorphism at 17q21.21(56) (white line with blue rectangles representing the variable segmental duplication (SD) blocks at the breakpoints), and the large credible set obtained by statistical fine-mapping including 2,178 SNPs in high LD (median(LD)=0.97) with the inversion (**Supplementary Table 7**). Pairwise LD values (*r*^2^) with lead variant rs1819040 were calculated from merged Italian, Spanish, German/Austrian and Norwegian GWAS discovery datasets. The dotted line indicates the genome-wide significance threshold (P=5×10^−8^). Below, organization of the 17q21.31 inversion genomic region, with the extended haplotypes associated with each orientation (H1 and H2) shown as red and blue arrows, respectively, and breakpoint SDs as dark rectangles. Protein-coding genes for which the inversion is a lead eQTL in at least one GTEx tissue are shown as pointed rectangles indicating the direction of transcription. **(b)** Forest plot and extended meta-analysis of our first discovery analysis and the COVID-19 HGI release 5 analysis B2 dataset (**Online Methods**) of the association between severe COVID-19 and the 17q21.31 inversion haplotype H2. Shown are OR and 95% CIs of the main (all), age-stratified (41-60 and 61-80 years (y/o)) across all analyzed cohorts. **(c)** Phenome-wide association study (PheWAS) results for the 17q21.31 inversion showing only potentially COVID-19 related phenotypes from the GWAS Catalog (*P*=10^−7^) grouped by disease categories using different colors. The effect direction of known SNP-trait associations is shown using triangles pointing upward (increase) and downward (decrease), whereas dots represent unknown effect direction. Phenotypes shown were selected according to previously reported COVID-19 links with lung damage, blood cell alterations and exacerbated immune response, as well as some potential co-morbidities. The whole list of phenotypic associations is included in **Supplementary Table 10**.

Next, we performed a replication analysis of the 13 variants reported to be genome-wide significantly associated with COVID-19 by the COVID-19 HGI using only individuals from the Severe COVID-19 GWAS group not included in the COVID-19 HGI release 5 datasets (**Supplementary Methods**). We replicated 8 of the 13 variants reported as genome-wide significant by the COVID-19 HGI(7) at least at nominal P-value of 0.05 (located near *LZTFL1, ABO, TYK2, DDP9, IFNAR2*, MAPT/*KANSL1, OAS1*) **Supplementary Table 5**).

Subsequently, we performed a fixed-effect inverse variance meta-analysis using METAL(13) across the first analysis and COVID-19 HGI B2 statistics as well as the second analysis and COVID-19 A2 statistics, resulting in a total of 725,601 (6,526 cases and 719,075 controls) and 1,320,760 (14,467 and cases and 1,306,293 controls) analyzed individuals in the respective analyses as well as 9,163,456 and 9,309,373 high quality variants (imputation R^2^≥0.6 in our cohorts and overall minor allele frequency (MAF) ≥1%; **Supplementary Figures 6-7**). This analysis prioritized rs1819040 at the 17q21.31 locus within the *KANSL1* gene as the most strongly associated variant in this region (P=3.27×10^−11^ for rs1819040; OR=0.88 for minor allele T; 95%CI=0.84-0.92) over rs8065800 from the first analysis (**Supplementary Tables 3**,**4**,**6**). The meta-analysis of our second discovery cohort (critically ill only) with the COVID-19 HGI summary statistics revealed an additional genome-wide significant locus not previously associated with severe COVID-19, chr19:50379362:T:C (rs1405655, P=3.25×10^−8^; OR=1.09; 95%CI=1.06-1.1) located in the *NAPSA* gene at 19q13.33; (regional association plot is shown in **Supplementary Figure 8, Supplementary Table 6**). Bayesian fine-mapping analysis of this locus with FINEMAP(15) (**Online Methods**) identified a total of 15 (log^10^(Bayes factor)=5.95) variants that belong to the 95% most likely to be causal (**Figure 1, Supplementary Table 7**).

### In-depth stratified analysis of lead variants

To estimate differences in genetic effects from age, sex and disease severity groups, we performed an in-depth stratified analysis for our discovery populations of the four replicable genome-wide significant hits from our first and second analyses, all of the 13 (including different variants at the aforementioned four loci) variants from the COVID-19 HGI release 5 analysis(7) and the novel association, chr19:50379362:T:C, at the 19q13.33 locus (**Supplementary Methods**). We additionally investigated association of these variants to known comorbidities such as hypertension, coronary artery disease and diabetes in cases only (**Supplementary Table 8**) and performed sex-stratified analysis within age and severity groups. We compared sex-specific and age-specific 95%CIs of ORs to determine statistical significance when CIs did not overlap. We confirmed our previously described association with age and disease severity at 3p21.31 for chr3:45848457:C:T (first analysis; ages 41-60; P=2.15×10^−23^, OR=2.20, 95%CI=1.88-2.57; ages 61-80; P=6.41×10^−6^; OR=1.49, 95%CI=1.25-1.77; severity: P=9.73×10^−7^, OR=1.65; 95%CI=1.35-2.03), which is also discussed in detail in a study by Nakanishi *et al*.(16) In addition, the chr17:46142465:T:A the (rs1819040) variant from the COVID-19 HGI study was associated with a protective OR that was consistently observed across the Italian, Spanish, and German/Austrian study cohorts in the 41-60 years age group (first analysis: P=4.59×10^−7^, OR=0.74, 95%CI=0.66-0.83), while no association was observed in the equally powered 61-80 years age group (P=0.491, OR=0.96, 95%CI=0.86-1.07, **Supplementary Table 8, Supplementary Figures 9-10**). This was especially pronounced in the male individuals. Non-significant (here comparing the overlap of the 95%CIs of the ORs), strong trends for association with age were observed for chr19:4717660:A:G (rs12610495; *DPP9*) at the 19p13.3 locus (first analysis: ages 41-60; P=3.8×10^−7^; OR=1.33, 95%CI=1.19-1.48; ages 61-80; P=0.0137; OR=1.15, 95%CI=1.03-1.28) and chr21:33242905:T:C (rs13050728; *IFNAR2*) which was significantly associated with severe COVID-19 only in younger individuals (first analysis; age <=60; P=5.01×10^−7^; OR=1.27, 95%CI=1.16-1.4; age > 60: P=0.109 OR=1.08; 95%CI=0.98-1.18). We did not observe any statistically significant associations with age, sex, severity or comorbidities for the remaining analyzed lead variants.

The genome-wide significant association near *LTZF1, TYK2, DDP9* and *ABO* have been followed up in detail before. Therefore, we next focused on the novel association at the 19q13.33 (*NAPSA*) locus from the meta-analysis with COVID-19 HGI release 5 summary statistics and an in-depth analysis of the 17q21.31 (*MAPT, KANSL1*) locus associated with age.

The 17q21.31 locus has been previously reported to be linked to a common inversion polymorphism.(17) Indeed, Bayesian fine-mapping(15) at this locus shows that the 95% credible set includes 1,530 variants in high linkage disequilibrium (LD; *r*^*2*^) (mean(*r*^*2*^)=0.997 and min(*r*^*2*^)=0.954) (among these chr17:45933112:G:A (rs8065800) from our initial discovery analysis and chr17:46142465:T:A (rs1819040) from the COVID-19 HGI analysis with certainty <0.3%, indicating that the individual SNP associations are only proxy variants for the likely causal variant, the inversion polymorphism (**Figure 1a, Supplementary Table 9**). We therefore imputed the inversion haplotypes H1 and H2 for our cohorts by genotype imputation with IMPUTE2(18), employing as reference 109 individuals from the 1000 Genomes Project for which 17q21.31 inversion genotypes were obtained experimentally by FISH and droplet digital PCR (**Online Methods**). LD between the chr17:46142465:T:A (rs1819040) variant, which was prioritized over chr17:45933112:G:A (rs8065800) in the meta-analysis with the COVID-19 HGI summary statistics, and the inversion in our cohorts is near perfect (*r*^2^=0.98, D’=0.99). Genome-wide significant association with severe respiratory COVID-19 for the inversion was confirmed using logistic regression followed by meta-analysis across this study’s discovery and replication panels from the COVID-19 HGI (meta-analysis first discovery panel and COVID-19 HGI release 5 B2: P=7.61×10^−10^, OR=0.89; 95%CI=0.84-0.92; meta-analysis second discovery panel and COVID-19 HGI release 5 A2: P=1.5×10^−4^, OR=0.90; 95%CI= 0.85-0.95; **Figure 1b, Supplementary Table 9**). Using stratified analysis, we also confirmed the age effect for the inversion (**Supplementary Table 7 and Figure 1b**). We did not observe any association of the inversion with disease severity.

### Functional analysis of 17q21.31 and 19q13.33 using publicly available datasets

We next performed several follow-up analyses to understand better possible functional implications of the associations at the 17q21.31 inversion and at the novel 19q13.33 locus, including a phenome-wide association study (PheWas) and exploratory gene expression analysis.

We queried variants in high LD (r^2^>0.9) with chr19:50379362:T:C (rs1405655) or the 17q21.31 inversion, using a wide range of phenotypes from the NHGRI GWAS Catalog (**Online Methods**).(19) While no known phenotypes were found to be linked to chr19:50379362:T:C (rs140565) or its proxy variants, 161 GWAS associations were identified for variants in high LD (r^2^>0.9) with the 17q21.31 inversion, illustrating its pleiotropic effects (**Supplementary Table 10**). These associations included several traits potentially related to COVID-19 pathology, such as blood and immune cell composition or lung function (**Figure 1c**).

Variants within the credible set from Bayesian fine mapping at 19q13.33 overlap with several genes, including for instance *NAPSA, NR1H2* and *KCNC3*, while the 17q21.31 inversion spans multiple genes including *MAPT, KANSL1, FMNL1* and *CRHR1* (**Supplementary Figures 4, 5 and 8** and **Figure 1a**). To identify possible target genes relevant in COVID-19 disease pathology, we performed an exploratory gene expression analysis using several publicly available datasets to: 1) Examine the direct effect of both loci on gene expression by using expression and splicing quantitative trait loci (eQTL and sQTL), 2) Identify in which tissues or cell types our candidate genes are expressed by analyzing their RNA expression at bulk and single-cell levels; and; 3) Infer the possible contribution of these genes to COVID-19 pathology by looking at their expression patterns in a) monocytes exposed to different viral and non-viral immune stimulators; b) organoids infected with COVID-19 and c) single-cell RNA-seq of lung and other tissues from patients who died after experiencing a SARS-CoV-2 severe disease (**Online Methods**). Results of these analyses are displayed in **Figure 2**.

**Figure 2.**
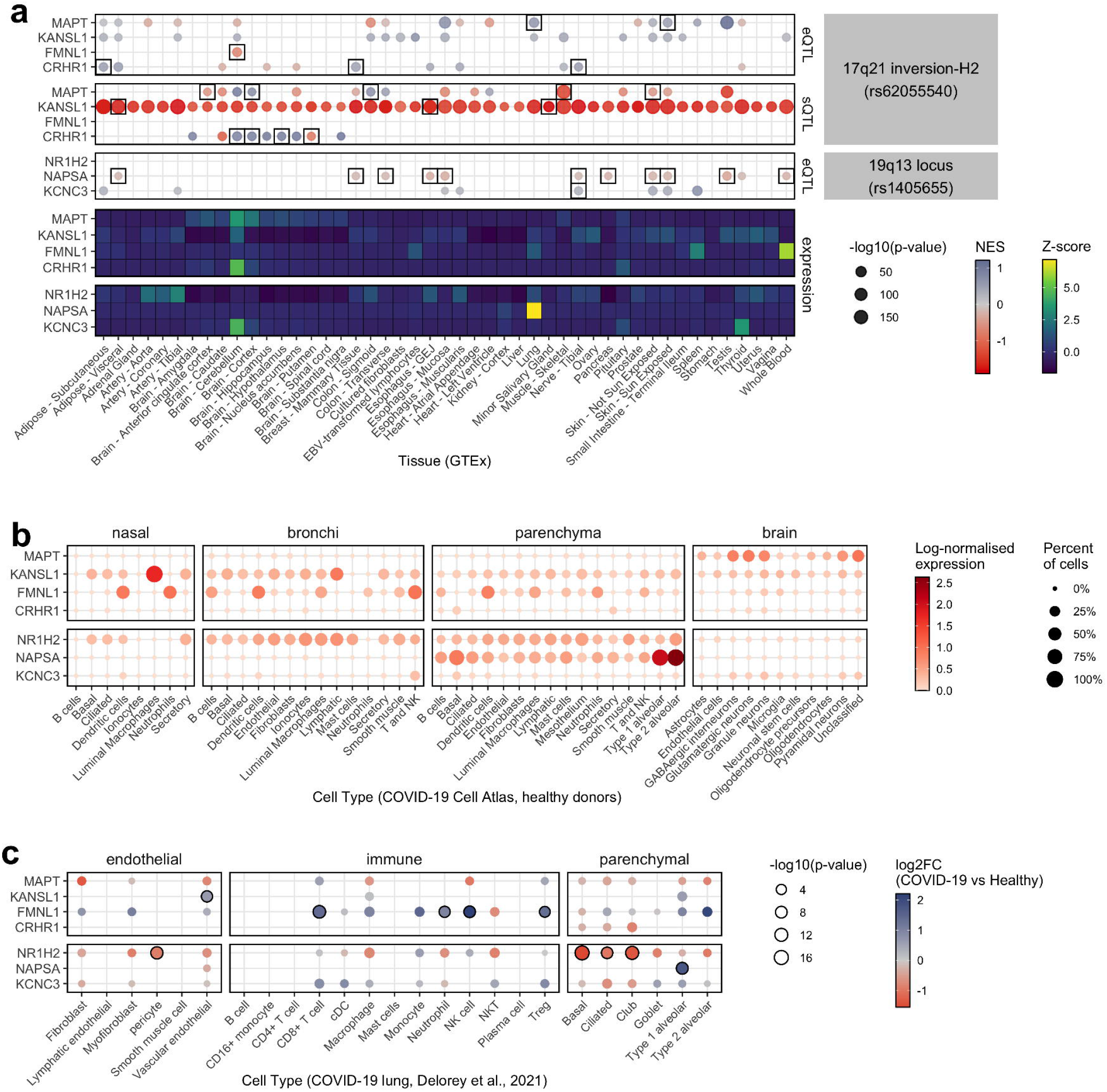
Expression analysis of the most plausible candidate genes associated with the 17q.21.31 and 19q.13.33 loci in organ tissues and COVID-19 relevant cell types. **(a)** GTEx tissue-specific expression QTL (eQTL, upper panel) and splicing QTL (sQTL, middle panel) effects of the 17q21.31 and 19q.13.33 loci on selected candidate genes as well as expression of these genes in GTEx(20) tissues (lower panel). Direction of normalized eQTL and sQTL effect size (NES) is represented by color intensities, and statistical significance by dot size. Effects are calculated on the 17q21.31 inversion haplotype H2 (here using the tag SNP rs62055540-C, LD (r^2^)=1), rs1405655-T or variants in high LD (r^2^>0.9) with either variant. Black rectangles indicate genes for which these variants or proxy variants in high LD (r^2^) are lead QTL variants in that tissue. If no box is shown other variants (i.e. not rs62055540-C, rs1405655-T or their proxy variants) are lead QTLs. Heatmap displays gene-wise centered median by tissue expression values (represented by color intensities), showing in which tissues candidate genes are mostly enriched. **(b)** Expression levels of candidate genes in scRNA-seq datasets from healthy upper airways (nasal, bronchi) and lung (parenchyma) cells(47) and adult human brain cells from recently deceased, non-diseased donors.(48) Figure displays log-normalized mean expression (represented by color) and fraction of cells expressing those genes (indicated by dot size). Processed and cell-type annotated gene expression levels from studies were retrieved from COVID-19 Cell Atlas.(49) **(c)** The figure shows differential expression of candidate genes in lung cells of COVID-19 patients compared to healthy controls. Log^2^ fold change (log2FC) values are presented as color gradient. Nominal p-values in - log^10^ scale are shown proportionally to dot size. Black-bordered circles indicate significantly differentially expressed genes after FDR correction. Results were obtained from pseudo-bulk differential expression analysis by Delorey *et al*.(21) More detailed figures are shown in **Supplementary Figures 11**,**12** and **15**.

The potential functional role of the 17q21.31 inversion is supported by the identification of 2,902 linked variants (*r*^*2*^>0.9) that are already reported as eQTL or sQTL in the GTEx Project.(20) The inversion is in LD with lead eQTLs and sQTLs (i.e. displaying the strongest association with target), for 24 (eQTL) and 7 (sQTL) genes, respectively (**Supplementary Table 11, Supplementary Figure 11**). Expression patterns of coding genes affected by the inversion are shown in **Supplementary Figure 12**, with the best candidates to play a role in the effects of the inversion on COVID-19, including *MAPT, KANSL1, FMNL1* and *CRHR1*, being summarized in **Figure 2**. Many of genes the coding genes affected by the inversion are highly expressed in neural and male-specific tissues (i.e. testis, **Supplementary Figure 12**). However, several of them are also expressed in major immune cell types in COVID-19 relevant tissues (**Figure 2b; Supplementary Figure 12a/b**), such as *KANSL1*, which is expressed in lung tissue-resident alveolar macrophages, or *FMNL1* (**Figure 2b, Supplementary Figure 12b**). These two genes especially show significantly higher expression in different lung cell types in COVID-19 patients versus healthy controls(21) (**Figure 2c, Supplementary Figure 12c**). Moreover, RNA-seq data of monocytes under different bacterial and viral stimuli(22) show expression changes in several genes affected by the inversion (**Supplementary Figure 13, Supplementary Table 12**). For example, *KANSL1*, which could have anti-inflammatory effects, shows higher expression in H2 carrier monocytes stimulated with Influenza A virus infection-like conditions, which appears to be related to a significant increase of coding and non-coding isoforms (**Supplementary Figure 14**). Similarly, in SARS-CoV-2 infected brain organoids, the expression of *MAPT* was significantly downregulated in premature and mature neuronal cells (**Supplementary Figure 15, Supplementary Table 13**).

In the case of the 19q13.33 locus, expression of candidate genes shows high tissue specificity, with *NAPSA* mRNA being specific to lung and lung parenchyma and *KCNC3* being highly expressed in brain and thyroid tissue, while *NR1H2* is more broadly expressed among human tissues, including many immune cell types (**Figure 2b, Supplementary Figure 12**). Of those candidates, expression of *KCNC3* and especially *NAPSA* appears to be clearly affected by the rs1405655 lead SNP (or variants on high LD, r^2^>0.9) (**Figure 2a**). Single SNP Mendelian randomization analysis using cis-eQTL data and assessment of effect heterogeneity identified higher expression of *NAPSA* as a potentially causal protective factor for severe COVID-19 (Beta=-0.39; P=1.26×10^−7^) (**Supplementary Methods, Supplementary Table 14**). Notably, *NAPSA* shows significantly increased expression in type 1 alveolar cells of COVID-19 patients as compared to healthy controls(21) (**Figure 2, Supplementary Figure 12**). On the other hand, in COVID-19 patients, the *NR1H2* gene is significantly down-regulated in some parenchymal (basal, ciliated, club) and endothelial (pericyte) lung cells and is up-regulated in monocytes compared to healthy controls (**Figure 2c, Supplementary Figure 12**).(21)

### Association analysis of specific candidate genomic regions: ABO locus, HLA locus & Y-chromosome haplogroups

The ABO locus is one of the most significant associations with COVID-19 infection, as highlighted in several publications.(5,7) In our initial publication, we showed that the genetic association at the ABO locus could be explained by differences in the actual ABO blood type. ABO secretor status, i.e. secretion of ABO antigens into body fluids, is determined by genetic variation in the *FUT2* gene. Recent studies suggest that A secretors have a higher risk of COVID-19 susceptibility.(23) Here we sought to replicate this finding (**Online Methods**) and compared 95%CIs for secretors and non-secretors of the A and B subtypes. 81% of all individuals were secretors irrespective of blood group on average across all control cohorts. In our study, though not statistically significant (95%CIs overlap), a clear trend was observed for increased risk of COVID-19 infection for A secretors (A secretors: OR=1.32; 95%CI=1.20-1.46; A non-secretors: OR=1.08; 95%CI=0.91-1.28) (**Supplementary Table 15**). Neither blood type B nor B secretion were significantly associated with COVID-19 susceptibility. When performing the stratified analysis as described above on the ABO locus, we saw a strong tendency for a more severe disease course in female A non-secretors in the Italian and Spanish cohorts, and a more severe disease course for males with blood type O, which did however not replicate in the comparably smaller German cohort. This should be followed-up in future studies.

The HLA fine-mapping approach yielded no association at the genome-wide or nominal (P<10^−5^) significance threshold, neither in the overall meta-analysis across the four cohorts, nor within the separate cohorts (**Supplementary Table 16, Supplementary Figure 16**). Furthermore, we found no significant association for any HLA-presented viral peptide in a so-called PepWAS approach (**Supplementary Table 17, Supplementary Figures 17-18**), where associations between HLA-presented peptides and disease is unravelled by integrating similarities and differences in peptide binding among HLA alleles across patients nor robust statistical associations with any of the other tested HLA parameters (**Supplementary Table 18**).

With male sex identified as a risk factor for severe COVID-19 and COVID-19-related death(3), we explored possible associations between genetic variants on the Y chromosome and the risk of developing severe COVID-19 and COVID-19 mortality in males.(3) Variations on the Y chromosome describe so-called Y-chromosome haplogroups with letters A-Z (defined by the Y Chromosome Consortium)(24) and follow a pattern of ancestral population migrations in Europe and on a global scale. Results of the Y-chromosome haplogroup analysis are shown in **Supplementary Table 19**. We did not observe any statistically significant or consistent results for Y chromosomal haplotype association with COVID-19 across the Spanish, Italian and German populations in our first or second analysis after correction of P-values for multiple testing using Bonferroni. We found that cohort-specific suggestive (P<0.05) associations within the Italian population for the R1b haplogroup were partially driven by single genotyping batches with diverging haplogroup frequencies (i.e. high coefficient of variation between control batches). COVID-19 related mortality showed a strong trend for association with haplogroup R1b1a2a1 (U106) (P=9.49×10^−3^, OR=2.8, 95%CI=1.29-6.2) in both the Spanish and Italian populations. This association remained after adjusting for the comorbidities hypertension, coronary artery disease and diabetes. COVID-19 related death remains, however, a challenging endpoint influenced by many factors.

## DISCUSSION

We here present a large collaborative COVID-19 genetics study of different centers from Italy, Spain, Norway and Germany/Austria. With our clearly defined phenotype of severe respiratory COVID-19, a centralized genotyping and rigorous quality control, we have generated a valuable resource for further COVID-19 related genetic studies. By conservatively including only severely affected COVID-19 patients in our analysis we aimed to balance out a potential bias from selection of controls from the general population, which is however general practice in GWA analyses. Our study replicated known associations with COVID-19 from our previous and other studies and suggest associations at four novel loci, *PCDH7* at 4p15.1; *FREM1* at 9p22.3; *OLMF4* at 13q21.1 and *PTPRM* at 18p11.23, which showed a high level of replicability across our cohorts but could not be replicated in the COVID-19 HGI release 5 statistics. Future studies in independent cohorts need to show whether these signals are true. Genome-wide meta-analysis of COVID19 HGI release 5 summary statistics with our data revealed a so-far unreported association the 19q13.33 locus, near the gene *NAPSA*. Our in-depth stratified analysis showed age-associations or trends for an association for *LZTFL1* at the 3p21.31 locus, *DPP9* at 19p13, the inversion at 17q21.31 and *IFNAR2* at 21q22.11. Here younger age groups (<= 60 years old; or aged 41-60) carrying the minor allele of variants associated at these loci, were more susceptible to COVID-19 than older age groups (>60 years old; or aged 61-80) carrying this allele. Effects of age in genetic variation are frequently observed in genetic studies, however reasons for this are not clear. Possible models for this are discussed in detail in Jiang *et al*.(25)

Our functional analysis focused on two loci of interest associated with severe COVID-19, the previously known 17q21.31 inversion and the 19q13.33 locus, including the *NAPSA* gene.

For novel association at the 19q13.33 locus, additional analyses provided first hints for a functional involvement in COVID-19 through its regulation of the *NAPSA* gene. *NAPSA* encodes for a protease highly expressed in Type 1 (AT1) and Type 2 (AT2) alveolar cells, two cell types required for the gas exchange at the lung surface and the secretion of surfactant proteins as well as immunomodulatory factors (AT2).(26) Complementary findings were observed in another recent study dissecting the lung transcriptome of COVID-19 infected patients in which *NAPSA* expression is increased in AT1 cells. This study also linked *NAPSA* to the marker gene expression signature of “damage-associated transient progenitors” (DAMPs), an intermediate cell state between AT1 and AT2 cells promoted by inflammation, characterized by a failure of AT2 cells to differentiate to AT1 cells.(27) Thus, given that the NAPSA protein is involved in lung surfactant production, which is dysregulated in COVID-19(28), the fact that the COVID-19 risk allele is associated with decreased *NAPSA* expression while increased expression of *NAPSA* is a protective factor for severe COVID-19, suggests a potential role of *NAPSA* in susceptibility for severe COVID-19. These findings are in line with a recent report in which *NAPSA* has been identified as a candidate gene for COVID-19 severity through cross methylome omnibus test combined with S-PrediXcan analyses in blood tissue and integrative multiomics(29), as well as with another recent report in which summary based Mendelian randomization analysis showed an inverse correlation between the expression of *NAPSA* in blood and COVID-19 severity.(30)

Moreover, we detected a clear association of the well-known and pleiotropic 17q21.31 inversion polymorphism, which is linked to many traits potentially relevant to COVID-19 outcome. For instance, the inverted haplotype H2 was previously associated with higher number of red blood cells and hemoglobin levels, whereas each haplotype correlates with different proportions of lymphocytes and granulocytes, which could potentially modulate the immune response during SARS-CoV-2 infection.(31) In addition, the H2 protective allele is also associated with decreased lung function and increased risk of chronic obstructive pulmonary disease but protects against development of pulmonary fibrosis(32), and is associated with higher ventilatory response to corticosteroids in individuals with asthma(33), showing potential trade-offs and shared pathways that may be important in lung health. This variant has been proposed to be under positive selection in Europeans through its effect on fertility.(17) Our results point to a role of this polymorphism in immunity and virus infection defence as well. Interestingly, inversion effects were found to be stronger in the younger age group in both severity classes, which could explain the weaker association in the HGI more severe A2 phenotype due likely to a larger proportion of older individuals.(7) The inversion probably affects COVID-19 disease course through its large effects on gene expression shown by us and others.(7) Although the function of many of the affected genes are not well known, the inversion acts as an eQTL and sQTL of several interesting candidate genes for severe COVID-19. In particular, there are several genes potentially associated with immune function and immune response. For example, *KANSL1*, involved in histone acetylation, is broadly expressed in many types of immune cells in upper airways and lung tissue (**Figure 2**) and has been proposed to play a role in the macrophage transition to an anti-inflammatory phenotype in mice.(34) Here, we have found that its expression decrease in infection-like stimulated monocytes is partially compensated in homozygotes for the H2 inversion haplotype. *CHRH1*, associated to higher expression in the H2 haplotype in several tissues (**Figure 2**), encodes a receptor that binds to corticotropin-releasing hormones, which are major regulators of the hypothalamic-pituitary-adrenal axis, and regulates immune and inflammatory responses.(35) Finally, *FMNL1*, which is also located in the inversion locus, shows high expression levels in macrophages, dendritic cells and B and T lymphocytes in different COVID-19 related tissues (**Figure 2**) and it is involved in cell motility and T cell trafficking.(36) In addition, many of the genes in the 17q21.31 inversion regions show a predominant expression in brain tissues, which could also play an important role in COVID-19. The clearest example is *MAPT*, which is downregulated in SARS-CoV-2 infected neuronal cells (**Supplementary Figure 15**) and lung-related cells and tissues. The H2 haplotype is linked to increased expression of *MAPT* in the lung and its *MAPT* exon 3 in brain tissues(37), which could compensate for the downregulation during viral infection and have a protective effect against COVID-19. However, despite the potential implication of these and other genes, it is not possible to single out just one as the most likely candidate. In fact, inversions are well-known for keeping together a combination of alleles from different genes that generate complex phenotypic traits in different organisms.(38,39) Our hypothesis-driven analysis of associations in the HLA, as well as COVID-19 specific PepWAS analyses yielded no significant results, indicating no major role for HLA variability in mediating the severity of COVID-19 in our cohorts. These results are in line with a recent, and so-far the largest, HLA analysis from Shachar et *al*.(40) Within our analysis of Y-chromosome haplogroups, none of the results remain significant after correction for multiple testing, though single haplogroups showed trends for association (**Supplementary Text**) and larger study samples are necessary to obtain reliable conclusions. For individual haplogroups, we observed different frequencies between batches that may also arise from different versions of the same genotyping platform. In general, the analysis of Y-chromosomal disease-relevant SNPs is mainly neglected in GWAS, hence the curation of Y-chromosomal SNPs on genotyping arrays is potentially also a confounding factor. Therefore, to gain more knowledge regarding the potential role of the Y-chromosome haplogroups in disease in general, this should be investigated in more depth.

In summary, our findings add to the number of genome-wide significant hits for COVID-19 – totalling now 14 independent loci – and provide new insights to the molecular basis of COVID-19 severity that could potentially trigger subsequent and more targeted experiments to develop therapies for severe COVID-19.

## METHODS

### Study Participants and Recruitment

We recruited 5,228 patients with mild to severe COVID-19, which was defined as hospitalization only (mild) or with respiratory failure (severe) with a confirmed SARS-CoV-2 viral RNA polymerase-chain-reaction (PCR) test from nasopharyngeal swabs or other relevant biologic fluids, cross sectionally, from intensive care units and general wards at different hospitals from Italy (4 centers, N=1,857), Spain (6 centers, N=2,795), Norway (7 centers, N=127) and Germany/Austria (8 German, 1 Austrian center, N=449). For comparison, we included 13,705 control participants from Italy (4 centers, N=5,247), Spain (3 centers, N=4,552), Norway (1 center, N=288) and Germany (1 center, N=3,582). Details on the centers and origin of the control panels are shown in **Supplementary Table 1a/b**. Though all patient samples that were sent to our study center were processed, only the severe COVID-19 individuals were analyzed in this study (**Supplementary Table 1e)**. Respiratory failure was defined in the simplest possible manner to ensure feasibility: the use of oxygen supplementation or mechanical ventilation, with severity graded according to the maximum respiratory support received at any point during hospitalization (1: supplemental oxygen therapy only, 2: noninvasive ventilatory support, 3: invasive ventilatory support, or 4: extracorporeal membrane oxygenation).(5)

### Recruiting Centers and Ethics Committee Approval IDs

The project protocol involved the rapid recruitment of patient-participants and no additional project-related procedures (we primarily used material from clinically indicated venipunctures) and afforded anonymity, owing to the minimal dataset collected. Differences in recruitment and consent procedures among the centers arose because some centers integrated the project into larger COVID-19 biobanking efforts, whereas other centers did not, and because there were differences in how local ethics committees provided guidance on the handling of anonymization or deidentification of data as well as consent procedures. Written informed consent was obtained, sometimes in a delayed fashion, from the study patients at each center when possible. In some instances, informed consent was provided verbally or by the next of kin, depending on local ethics committee regulations and special policies issued for COVID-19 research. For some severely ill patients, an exemption from informed consent was obtained from a local ethics committee or according to local regulations to allow the use of completely anonymized surplus material from diagnostic venipuncture. Centers from which samples were obtained are listed together with their ethics approval reference numbers from each ethic committee in **Supplementary Table 1b**.

### Sample Processing, Genotyping, Quality control, and Imputation

Detailed description on sample processing, genotyping, genotype quality control and genotype imputation can be found in the **Supplementary Methods**. In brief: The majority of samples were processed at the DNA laboratory of the Institute of Clinical Molecular Biology (Christian-Albrechts-University of Kiel, Germany), with a subset of the German samples (BoSCO study) processed and typed at the Genomics Department of Life&Brain Center, Bonn, and another subset (COMRI study) prepped at the Technical University Munich, Munich, Germany and genotyped at the Genotyping laboratory of the Institute for Molecular Medicine Finland FIMM Technology Centre, University of Helsinki, Finland. The German control samples were prepped at the Institute of Clinical Molecular Biology and genotyped at the Regeneron Genetics Center, U.S.A. In brief, prepped DNA extracts or non-prepped whole blood, buffy coat samples (and for an exceedingly small subset also saliva) genotyped on Illumina’s Global Screening Array (GSA), version 1.0 (German controls), 2.0 or 3.0 (BoSCO study, COMRI study and 378 cases and 1,180 controls from the Italian cohort) with a SNP coverage of 700,078 to 730,059 variants. We performed a uniform sample and single-nucleotide polymorphism (SNP) quality control across the German/Austrian, Italian, Norwegian and Spanish populations respectively. After the exclusion of samples during quality control (the majority due to population outliers) (**Supplementary Table 1**), the final case–control datasets comprised 1,536, patients and 4,759 control participants from Italy, 1,421 patients and 4,377 control participants from Spain, 62 patients and 262 controls from Norway and 241 patients and 3,110 control participants from Germany. The number of SNPs pre-imputation were 664,969 for the Italian cohort, 669,359 for the Spanish cohort, 663,411 for the Norwegian cohort and 568,542 for the German/Austrian cohort. To maximize genetic coverage, we performed SNP imputation on genome build GRCh38 using the Michigan Imputation Server and 194,512 haplotypes generated by the Trans-Omics for Precision Medicine (TOPMed) program (freeze 5)(41) (**Supplementary Methods**). The number of SNPs with a post-imputation score (R^2^) of > 0.1 were 77,767,912 for the Italian cohort, 72,504,622 for the Spanish cohort 19,466,514, for the Norwegian cohort and 51,399,774 for the German/Austrian cohort.

### Statistical Analysis

#### Genome-wide association analysis

To take imputation uncertainty into account, we tested for phenotypic associations with allele dosage data separately for the Italian, Spanish, German/Austrian, and Norwegian case-control data. We carried out a logistic regression analysis corrected for potential population stratification, age and sex bias using the SAIGE software(12) (**Supplementary Methods**). An inverse-variance weighted fixed-effects meta-analysis was conducted with the meta-analysis tool METAL(13) on the first discovery cohort including 3,260 cases and 12,483 population controls of unknown COVID-19 status from Italy, Spain, Norway and Germany/Austria and the second discovery cohort including 1,911 critically ill cases and 12,483 population controls (**Supplementary Table 1**. Only variants that were common to at least 2 datasets with respective post-imputation R^2^ ≥0.6 and that had an overall minor allele frequency (MAF) of ≥1% were considered in the analysis. For each variant, we computed across-cohort heterogeneity P-values and I^2^ values using METAL. We used a significance threshold of P<10^−6^ for joint P-values to determine suggestive statistical significance and the common threshold of P<5×10^−8^ to determine genome-wide significance. Meta-Analysis Model-based Assessment of replicability(14) (MAMBA) was used (**Supplementary Methods**) to assess replicability at each locus.

To replicate 13 genome-wide significant associations reported by the COVID-19 HGI(7), we excluded 3,837 individuals overlapping between our current study and COVID-19 HGI release 5. We subsequently carried our logistic regression and meta-analysis as described above in 1,579 cases and 10,372 population controls for the first analysis and 944 cases and 10,065 population controls for the second analysis (**Supplementary Table 1e**). A threshold of 0.05 was used to define replicability of the results. Meta-analysis with the COVID-19 HGI release 5 summary statistics included up to 12,888 hospitalized cases (including 5,582 critically ill cases) and 1,295,966 population controls of the COVID-19 HGI. To unify case definitions by our study and the COVID-19 HGI, we performed meta-analysis of summary statistics from our first analysis with COVID-19 release 5 A2 summary statistics and meta-analysis of our second analysis and COVID-19 release 5 B2 summary statistics. We used the common threshold of P<5×10^−8^ to determine genome-wide significance

Fine mapping of candidate SNPs was performed using Bayesian fine mapping with the tool FINEMAP(15) (**Supplementary Methods**).

#### Association analysis of candidate SNPs

We performed age- and sex-stratified and severity analyses on 4 candidate SNPs from our first and second discovery cohorts, and 13 candidate SNPs from the COVID-19 HGI analysis.(7) We additionally analysed comorbidities hypertension, coronary artery disease and diabetes in cases only. We carried out a logistic regression analysis corrected for potential population stratification, age and sex bias using the software R version 3.6.2 (**Supplementary Methods**). The inverse-variance weighted fixed-effects meta-analysis was conducted using the R-package metafor(42) including only statistics from cohorts with N^Case^ and N^Control^ > 50. Sub-analyses in age groups of >60, <=60, 20-40, 41-60, 61-80 and > 80 years were carried out, with the highest sample numbers and statistical power in the age groups of 41-60 and 61-80 years and the binarized age groups of <=60 and >60 (**Supplementary Table 1e**).

#### Association analysis of the 17q21.31 inversion

17q21.31 inversion genotypes were imputed using IMPUTE v2.3.2(43) from quality controlled SNP data (first and second analysis) based on experimentally-validated inversion genotypes from 109 individuals from the 1000 Genomes Project(44) or from P-values (COVID-19 HGI A2/B2 release 5 data) using Fast and accurate P-value Imputation for genome-wide association study (FAPI) (**Supplementary Methods**).(45) The inversion was coded as 0 for the major allele H1 and 1 for the minor allele H2. All association analyses were carried out as described above on the minor allele H2.

#### Functional analysis

Phenotype associations with the different variants were obtained from the NHGRI-EBI GWAS Catalog(19). We queried GWAS hits in high LD (r^2^ > 0.9) with the inversion as well as hits in high LD (*r*^2^ > 0.9) with rs1405655 (19q13.33). Similarly, tissue-specific expression or splicing effects were obtained by searching for SNPs in high LD (*r*^2^>0.9) with the inversion or rs1405655 that have been already identified as expression quantitative trait loci (eQTLs) or splicing quantitative trait loci (sQTLs) in *cis* by the GTEx Project (GTEx Analysis Release v8).(20) Candidate coding genes were selected based on their inclusion in the GWAS credible sets and/or if any of the variants had been identified as lead eQTL or sQTL (**Supplementary Table 19)**. Exploratory gene expression analysis of selected candidates was performed on publicly available pre-processed RNA-seq datasets generated from organ tissues (GTEx Analysis Release v8 immune cell types (BLUEPRINT(46)), as well as respiratory tract(47) and brain cells(48) (COVID-19 Cell Atlas).(49) Differential expression of candidate genes in COVID-19 infected lung cells were obtained from pseudo-bulk differential expression analysis performed by Delorey *et al*.(21) Since several candidate genes (including *MAPT, CRHR1* and *KANSL1*) were highly expressed in the neural system, differential gene expression was also analyzed on single-cell RNA-seq dataset of COVID-19 infected brain organoid cells from Song *et al*.(50) (obtained upon request). The analysis was carried out using hurdle modeling, implemented in the R package MAST.(51) Finally, to check the effect of the 17q21.31 inversion on monocytes stimulated by infection-like conditions we also performed a differential expression analysis in the RNA-seq data from Quach *et al*.(22) by imputing the inversion genotypes with IMPUTE v2.3.2(18) and quantifying gene and transcript expression differences with Kallisto v0.46.0(52) and QTLtools v1.1.(53) For detailed description, refer to **Supplementary Methods**.

#### Analysis and fine mapping of the HLA

Detailed descriptions of this analysis can be found in the **Supplementary Methods**. In brief, quality-controlled genotypes at the HLA region (chr6:29-34Mb) were extracted. HLA allele, amino acid, and SNP imputation was performed using the random-forest based HLA genotype imputation with attribute bagging (HIBAG) and applying specially tailored as well as publicly available reference panels.(5,54,55) The resulting data were used as a basis for several subsequent analyses, including: 1) basic association analysis (fine mapping) as described in the section **Statistical Analysis** and the **Supplementary Methods**, 2) a peptidome-wide association study(11) (pepWAS), to screen for disease-relevant peptides from SARS-CoV-2, that may present a possible functional link between severe COVID-19 and variation at classical HLA loci, 3) quantitative HLA analyses directed at the number of peptides bound by an HLA allele, as well as 4) an analysis of HLA-presentation of shared peptides (‘molecular mimicry’).

#### Analysis of the Y-chromosome haplotypes

First, we produced high quality Y-chromosome genotypes by manually calling and visually inspecting Y-chromosome SNPs in the male fraction of the cohorts only. Next, we used 22 Y-chromosome SNPs to distinguish known Y-chromosome haplogroups as described in the **Supplementary Methods** at different haplogroup resolutions. We here focused on haplogroup R, the most prevalent Y-chromosome haplogroup across Europe. Association analysis was carried out as described in the Section **Statistical Analysis – Association analysis of candidate SNPs** on Y-chromosome haplogroups coded as absent (0) or present (1). We additionally analyzed the end-point mortality (**Supplementary Methods**). The coefficient of variation (CV) across frequencies was calculated for Y-chromosome haplogroups between individual batches (i.e. contributing center/hospital or different versions of the GSA). Here only frequencies that were observed in more than 100 individuals were considered.

#### Analysis of the ABO secretor status

ABO blood group typing was performed as described by Ellinghaus *et al*.(5) Briefly, genotypes of the SNPs rs8176747 (c.803C>T, *ABO*B*), rs41302905 (c.802G>A, *ABO*O2)* and rs8176719 (c.261delG, *ABO*O1*) were extracted from the imputed data (R2=1 for all SNPs and cohorts) and used to infer the A, B and O blood types. The ABO-”secretor” status was inferred from the genotypes of the rs601338 SNP (c.428G>A, *FUT2*01N.02*) at the *FUT2* gene, located at 19q13.33, extracted from the imputed data (R2=0.98-0.99 for all cohorts). Individuals carrying genotypes GA or GG were assigned secretor status and individuals carrying genotype AA were assigned non-secretor status based on the genotype dosages, ranging from 0 to 2, retrieved from the imputed data. Individuals with allelic dosages 1.3-1.7 were called as “no call”, individuals with dosages ≤1.3 were called “secretors” and individuals with dosages ≥1.7 were called “non-secretors”. Association analysis was carried out as described in the Section **Statistical Analysis – Association analysis of candidate SNPs** on blood type or blood type secretor status coded as absent (0) or present (1).

#### Availability of Summary Statistics

Genome-wide summary statistics of our analyses will be made available upon acceptance of this manuscript.

## Data Availability

Additional details of the analyses may be requested from the corresponding authors and data will be shared per the authors approval.

## CONTRIBUTIONS

Contributions are shown in the **Supplementary Note**.

## ACKNOWLEDGEMENTS

We would like to thank the COVID-19 Human Genetics Initiative (HGI) for collecting and openly sharing extensive summary statistics with the community.

For the cohort from Milan Fondazione Ca’ Granda we would like to acknowledge specifically Rossana Carpani. A full list of the investigators who contributed to the generation of the GCAT data is available from www.genomesforlife.com. We thank Dr. Luis Puig and Vanessa Pleguezuelos on behalf of the Blood and Tissue Bank from Catalonia (BST) who collaborated in the GCAT/COVICAT recruitment, and all the GCAT volunteers that participated in the study. We would also like to acknowledge the Task Force Humanitas and the Task Force Humanitas Gavazzeni Castelli (**Supplementary Note**). We would like to acknowledge all the participants from the “Grupo de Trabajo en Medicina Personalizada contra el COVID-19 de Andalucia”. We also thank “Consejeria de Salud y Familias” and “Junta de Andalucía” for their support and funding that is currently supporting the COVID-19-GWAS and the COVID-PREMED initiatives. We are indebted to the HCB-IDIBAPS Biobank for the biological human samples and data procurement and to the Fundació Glòria Soler for its support to the COVIDBANK collection. We acknowledge the work of the Norwegian SARS-CoV-2 Study group (**Supplementary Note**). We also acknowledge Benedicte A. Lie at the Department of Immunology, Oslo University Hospital and the Norwegian Bone Marrow Donor Registry are acknowledged for providing healthy controls. We thank the support of the German COVID-19 multiomics initiative (decoi.eu). We thank the members of the COVID-19 use and access committee (Prof. Reinhold Förster, Prof. Christine Falk, Prof. Thomas Fühner, Prof. Christoph Höner zu Siederdissen, Prof. Marius Höper, Prof. Thomas Friedrich Schulz, Prof. Markus Cornberg, Prof. Thomas Illig and Prof. Michael Manns), as well as the study nurses, medical doctors and biobank staff members involved in the project. For funding we thank the Ministry of Science and Culture (MWK) of Lower-Saxony. We thank everybody involved in the COVID-19 registry at the University Hospital Regensburg (COVUR). Genotyping of the German control dataset was performed at the Regeneron Genetics Center. Genotyping of the COMPRI study was performed by the Genotyping laboratory of Institute for Molecular Medicine Finland FIMM Technology Centre, University of Helsinki. We are very grateful to Professor Akiko Iwasaki and Eric Song for generously sharing pre-processed single-cell RNA-seq data generated from SARS-CoV-2 infected brain organoids.

## ABBREVIATIONS

95%CI: 95% confidence interval
AT1: Type 1 alveolar cells
AT2: Type 2 alveolar cells
CAD: Coronary artery disease
CHRH1: Corticotropin Releasing Hormone Receptor 1
COVID-19: Coronavirus disease 2019
COVID-19 HGI: COVID-19 Host Genetics Initiative
DAMPs: Damage-associated transient progenitors
DNA: Deoxyribonucleic acid
ECMO: Extracorporeal membrane oxygenation
eQTL: Expression quantitative trait loci
FISH: Fluorescence in situ hybridization
FMNL1: Formin Like 1
GenOMICC: Genetics Of Mortality In Critical Care Initiative
GSA: Global Screening Array
GTEx: Genotype-Tissue Expression project
GWAS: Genome-wide association analysis
HLA: Human Leukocyte Antigen
KANSL1: KAT8 Regulatory NSL Complex Subunit 1
KCNC3: Potassium Voltage-Gated Channel Subfamily C Member 3
LD: Linkage disequilibrium
MAF: Minor allele frequency
MAPT: Microtubule Associated Protein Tau
Mb: Megabase pairs
NAPSA: Napsin A Aspartic Peptidase
NES: normalized effect size
NR1H2: Nuclear Receptor Subfamily 1 Group H Member 2
OR: Odds ratio
P: P-value of association
PCA: Principal component analysis
PCR: Polymerase-chain reaction
PepWAS: Peptidome-wide association study
PheWAS: Phenome wide association study
PPR: Posterior probability of replication
QC: Quality control
RNA: Ribonucelic acid
RNA-seq: RNA sequencing
scRNA-seq: Single sell RNA-seq
SNP: Single nucleotide Polymorphism
sQTL: Quantitative splicing trait loci

