## Supplement for "Detailed stratified GWAS analysis for severe COVID-19 in four European populations"

### Supplementary Appendix

This appendix has been provided by the authors to give readers additional information about their work.

#### Table of Contents

***Supplementary Note*** ... ***1***

**Author contributions** ... **1**

**Additional authors from study groups** ... **2**

Task Force COVID-19 Humanitas ... 2

TASK FORCE COVID-19 HUMANITAS GAVAZZENI & CASTELLI ... 3

COVICAT Study Group ... 5

Pa COVID-19 Study Group ... 5

Covid-19 Aachen Study (COVAS) ... 5

Norwegian SARS-CoV-2 Study group ... 6

***Supplementary Methods*** ... ***8***

**DNA extraction and SNP genotyping** ... **8**

DNA extraction Institute of Clinical Molecular Biology ... 8

DNA extraction for the GCAT cohort ... 9

DNA extraction Genomics Department of Life&Brain Center, Bonn ... 9

DNA extraction COMRI cohort ... 9

Genotyping at the Institute of Clinical Molecular Biology ... 9

Genotyping at the Life&Brain Center, Bonn, Germany ... 10

Genotyping at the Regeneron Genetics Center ... 10

Genotyping at the Genotyping laboratory of Institute for Molecular Medicine Finland FIMM Technology Centre, University of Helsinki ... 10

**Genotype calling, quality control and imputation** ... **11**

Genotype calling ... 11

SNP and sample quality control and principal component analysis ... 11

SNP genotype imputation ... 12

Imputation of the ~0.9-Mb inversion polymorphism at 17q21.31 ... 12

***REFERENCES*** .......... *Error! Bookmark not defined.*

#### Supplementary Note

##### *Author contributions*

T.H.K. conceived and initiated the project. T.H.K. and A.F. jointly designed, provided infrastructure to and jointly supervised the project. F.D., D.E., S.J, M.C., J.L-J., T.L.L., M.W., H.EA., O.O., J.A., L.V., R.A., R.d.C., D.M-M. and A.F. wrote the first draft of the manuscript. A.F., D.E. F.D., K.L. and E.C.S vouche for the genetic data. F.D., D.E. (genetics), S.J., M.C., J.L-J. (expression and inversion) and M.We. and T.L.L. (HLA) coordinated and performed the statistical analyses with contributions from L.W., F.U-W., E.M.W (genetics), H.EA., J.A., O.Ö. (HLA), R.M., O.B.L., M.S.V. (Y-Chromosome analysis), M.Wi (ABO analysis) and M.C.R. (Mendelian Randomization). A.Al., A.La., B.H., E.C.S., F.T., H.Z., J.C.H., J.Fe., J.H., J.R.H., K.U.L., L.B., L.V., M.Bu., M.Rom., P.Ba., P.I., P.K., R.A., R.d.C., S.Dug., S.G., T.H.K organised and supervised patient inclusion, vouche for the clinical data, and provided input to study design and the manuscript. A.Ag., A.B.K., A.B.O., A.Ban., A.Bar., A.Bi., A.Bl., A.Br., A.C.N., A.Cab., A.Cav., A.Ch., A.d.S., A.G.C., A.Gan., A.Gar., A.Ge., A.Gl., A.Go., A.H., A.J., A.L.F., A.Li., A.Ll., A.M.D., A.Man., A.May., A.Mu., A.N., A.Pa., A.Pesc., A.Pese., A.Po., A.Pr., A.R.H., A.Ram., A.Ran., A.Rue., A.Rui., A.S., A.Ta., A.Te., A.V., A.Z., B.C., B.J., B.M., B.N., B.S., C.An., C.Az., C.Be., C.Bi., C.Ca., C.Ce., C.D.S., C.d., C.F., C.G., C.He., C.Hu., C.La., C.Le., C.M., C.P., C.Q., C.Sa., C.Sc., C.Sk., C.T., D.A., D.G., D.H., D.J., D.Pe., D.Pr., D.T., E.Ar., E.Az., E.B., E.C., E.J.C., E.M.P., E.M., E.N., E.P., E.R., E.Sa., E.Sc., E.So., E.T.H., E.U., F.A., F.B., F.C., F.G.S., F.G., F.H., F.J.M., F.K., F.Mal., F.Mar., F.Me., F.Mü., F.P., F.R., G.A., G.Ba., G.Be., G.Ca., G.Ci., G.Co., G.F., G.G., G.H., G.L., G.M., G.P., H.E., H.K., H.N., I.d.R., I.G., I.H., I.K., I.M., I.P., J.Al., J.Am., J.Ba., J.Be., J.De., J.Do., J.E.A., J.E., J.Fa., J.G., J.H.Q., J.K.D., J.Kä., J.Kr., J.M.B., J.M.G., J.M., J.R.B., J.R., J.Schn., J.Schr., J.W., K.B., K.E.M., K.G., K.I.G., K.R., K.T., L.E.S., L.G., L.H., L.I., L.J.L., L.K., L.N., L.R.B., L.R., L.Sa., L.Sc., L.Su., L.T.G., L.Té., L.Te., M.A.G., M.Ac., M.Au., M.Ba., M.Boa., M.Boc., M.Car., M.Cas., M.Caz., M.Ce., M.Ci., M.Cord., M.Corn., M.D'Am., M.D'An., M.Dr., M.E., M.E.N., M.G.V., M.H., M.I., M.J.S., M.J.V., M.J., M.K., M.M.B., M.M.G., M.M.N., M.Man., M.Mar., M.Maz., M.Mi., M.O., M.Ri., M.Rod., M.S., M.T., M.Wi., N.Bl., N.Br., N.C., N.I.A., N.L., N.Mart., N.Marx., N.Mo., O.A.C., O.P., O.W., P.Bo., P.C., P.F., P.H., P.M.R., P.O., P.P.E., P.P., P.R., P.Sc., P.Su., P.T., R.B., R.C., R.d.P., R.F., R.Ga., R.Gu., R.Mo., R.N., S.Al., S.An., S.Ba., S.Bom., S.Bos., S.Br., S.C., S.Dud., S.Ha., S.He., S.K., S.Mar., S.May., S.Pe., S.Pi., S.Ra., S.Ri., S.Ro., S.S., S.W., T.Ba., T.Br., T.E., T.Fe., T.Fo., T.G.C., T.I., T.P., T.W., T.Z., U.H.,

U.L., U.P., V.A., V.F., V.Ke., V.Ko., V.Mon., V.Mor., V.R., V.S., W.A., W.Pe., W.Po., X.D., X.F., X.W., X.Y., Y.K., Z.K. provided samples, processed samples, provided phenotypic data or intellectual input to the manuscript. All authors revised and edited the manuscript for critical content and approved of the final version to be published.

***Additional authors from study groups***

***Task Force COVID-19 Humanitas***

**IRCCS Humanitas Research Hospital, Rozzano, Milan, Italy**

Accornero Stefano, Alfarone Ludovico, Ali Hussam, Aloise Monia, Anfray Clement, Angelini Claudio, Arcari Ivan, Arosio Paola, Azzolini Elena, Baccarin Alessandra, Badalamenti Salvatore, Baggio Sara, Balzarini Luca, Barbagallo Michela, Barberi Caterina, Barbieri Viviana, Barbone Alessandro, Basciu Alessio, Belgiovine Cristina, Benvenuti Chiara, Bertocchi Alice, Bianchi Ilaria, Bocciolone Monica, Bombace Sara, Bonifacio Cristiana, Borea Federica, Borroni Mario, Brescia Paola, Bresciani Gianluigi, Brunetta Enrico, Bulletti Cinzia, Cadonati Cristina, Calabro’ Lorenzo, Calatroni Marta, Calcaterra Francesca, Caltagirone Giuseppe, Calvetta Albania Antonietta, Calvi Michela, Cancellara Assunta, Cannata Francesco, Canziani Lorenzo, Capogreco Antonio, Capretti Giovanni Luigi, Capucetti Arianna, Carenza Claudia, Carlani Elisa, Carloni Sara, Carnevale Silvia, Carrone Flaminia, Casana Maddalena, Castelli Alice, Castelnuovo Elena, Cazzetta Valentina, Ceribelli Angela, Ceriotti Carlo, Chiarito Manuel, Ciccarelli Michele, Cimino Matteo, Citterio Gianluigi, Ciuffini Leonardo, Colaizzi Chiara, Colapietro Francesca, Costa Guido, Cozzi Ottavia, Craviotto Vincenzo, Crespi Chiara, Crippa Massimo, Da Rio Leonardo, Dal Farra Sara, D'antonio Federica, D'antuono Felice, Darwich Abbass, De Ambroggi Guido, De Donato Massimo, De Lucia Francesca, De Nittis Pasquale, De Paoli Federica, De Santis Maria, Delle Rose Giacomo, Desai Antonio, Di Donato Rachele, Di Pilla Marina, Digifico Elisabeth, Dipaola Franca, Dipasquale Andrea, Dipasquale Angelo, D'orazio Federico, Droandi Ginevra, Durante Barbara, Farina Floriana Maria, Favacchio Giuseppe, Fazio Roberta, Fedeli Carlo, Ferrante Giuseppe, Ferrara Elisa Chiara, Ferrari Valentina, Ferrari Matteo Carlo, Ferri Sebastian, Folci Marco, Foresti Sara, Fornasa Giulia, Franchi Eloisa, Franzese Sara, Fraolini Elia, Furfaro Federica, Furlan Raffaello, Galimberti Paola, Galtieri Alessia, Gardini Maria, Gavazzi Francesca, Generali Elena, Giannitto Caterina, Gil Gomez Antonio, Giorgino Massimo Giovanni, Giugliano Silvia, Goletti Benedetta, Gomes Ana Rita, Guarino Elisabetta, Guerrini jacopo, Guidelli Giacomo, Jacobs Flavia, Kurihara Hayato, Lagioia Michele, Lania

Andrea Gherardo, Lanza Ezio, Lavezzi Elisabetta, Libre' Luca, Lizier Michela, Lleo Ana, Lo Cascio Antonino, Loiacono Ferdinando, Loy Laura, Lughezzani Giovanni, Lutman Fabio, Maccallini Marta, Magnoni Paola, Maiorino Alfonso Francesco, Mantovani Riccardo, Marchettini Davide, Marinello Arianna, Markopoulos Nikolaos, Marrano Enrico, Masetti Chiara, Mazziotti Gherardo, Melacarne Alessia, Milani Angelo, Mirani Marco, Morelli Paola, Motta Francesca, Mozzarelli Alessandro, Mrakic Sposta Federica, Mundula Valeria, My Ilaria, Nasone Irene, Nigro Mattia, Omodei Paolo, Oresta Bianca, Ormas Monica, Pagliaro Arianna, Paiardi Silvia, Paliotti Roberta, Pasqualini Fabio, Pasto' Anna, Pavesi Alessia, Pedale Rosa, Pedicini Vittorio, Pegoraro Francesco, Pelamatti Erica, Pellegatta Gaia, Pellegrino Marta, Perucchini Chiara, Pestalozza Alessandra, Petriello Gennaro, Piccini Sara, Pivato Giorgio, Pocaterra Daria, Poliani Laura, Poretti Dario, Pozzi Chiara, Preatoni Paoletta, Procopio Fabio, Profili Manuel, Puggioni Francesca, Pugliese Luca, Pugliese Nicola, Racca Francesca, Randazzo Michele, Regazzoli Lancini Damiano, Reggiani Francesco, Rimoldi Valeria, Rimoldi Monica, Ripoll Pons Marta, Rodolfi Stefano, Ronzoni Giulia, Ruongo Lidia, Sacco Clara, Sagasta Michele, Sandri Maria Teresa, Sarra Giuseppe, Savi Marzia, Scarfo' Iside, Scarpa Alice, Shiffer Dana, Sicoli Federico, Silvestri Alessandra, Sironi Marina, Solano Simone, Solitano Virginia, Spadoni Ilaria, Spano' Salvatore, Spata Gianmarco, Stainer Anna, Stella Matteo Carlo, Strangio Giuseppe, Supino Domenico, Taormina Antonio, Tentorio Paolo, Teofilo Francesca Ilaria, Testoni Lucia, Tordato Federica, Torrisi Chiara, Trabucco Angela, Ulian Luisa, Ummarino Aldo, Valentino Rossella, Valentino Sonia, Valeriano Chiara, Vena Walter, Verlingieri Simona, Vespa Edoardo, Voza Antonio, Voza Giuseppe, Zaghi Elisa, Zanon Veronica, Zanuso Valentina, Zilli Alessandra, Zumbo Aurora.

***TASK FORCE COVID-19 HUMANITAS GAVAZZENI & CASTELLI***

**Humanitas Gavazzeni-Castelli, Bergamo, Italy**

Abati Elena Maria, Abualkheir Mohammed, Agradi Sergio, Albano Giovanni, Alioto Giuseppina, Allegrini Davide, Altieri Vincenzo Maria, Angeli Enzo, Arduini Mario, Assisi Alberto, Baldi Marzia, Barbagallo Lucia, Barzaghi Maria Elena, Basili Luca Manfredi, Beretta Alessandra, Beretta Natascia, Beretta Giordano, Beretta Simone, Bertella Erika, Bettoni Elisabetta, Biazzo Alessio, Bombardieri Emilio, Bonacina Manuela, Bonfanti Riccardo, Bordoni Mariagrazia, Bordoni Luca, Borghesi Maria Lucia, Bortolotti Luigi, Bravi Marco, Brena Federica, Bruno Simona, Camozzini Valentina, Canziani Lorenzo Maria, Caporarello Salvatore, Cappelleri Gianluca, Caputo Rosilde, Carrara Alfonso, Carriero Federica, Castoldi Massimo, Castriota Fausto, Catellani Francesco, Catenacci Alberto, Celotti Simona, Cereda

Marco Angelo, Ceresoli Giovanni Luca, Cesari Eugenio, Chiesa Giuseppe, Colli Alessandro, Corapi Antonio, Cordua Nadia, Cortina Gabriele, Coscione Andrea Vittorio, Costa Maria Concetta, Cremonesi Alberto, Dalto Serena, D'Aquino Nicola, D'Aveni Alessandro, De Amicis Francesco, De Filippis Costantino Nicandro, Delalio Elena, Dell'Era Valentina, Di Cintio Davide, Di Noia Vincenzo Pio, Esposito Giovanni, Fedele Isabella, Ferrari Elisa, Filippi Claudia, Finamora Ilaria, Fiorentino Gennaro, Fortino Olga, Franceschini Grisolia Enrico, Frongillo Elisabettamaria, Fumagalli Miriam, Gabuda Marian Svyatoslavovich, Gaffuri Nicola, Galbussera Maurizio, Galdamez Carcamo Shintia Coralia, Galli Sara, Gentinetta Franco, Gerometta Piersilvio, Ghilardi Carlo Guardo, Ghirardelli Paolo, Ghirardi Guido Luciano, Ghisi Patrizia Chiara, Giambartino Sebastiano, Giofre' Fabrizio, Giroletti Laura, Goletti Orlando, Graniero Ascanio, Grassi Massimo Maria, Grazioli Valentina, Guanella Giovanni Battista, Guidotti Eugenio, Intelisano Antonio, Lanzone Alberto Maria, Ledda Giovanna Franca, Licini Gloria, Liguori Alessia, Lisanti Rocca Carmela, Lleshaj Eliona, Loffreno Antonella, Lopresti Ennio, Lucca Elena, Macca Claudio, Macchini Daniele, Maggi Lamberto, Magni Luca, Mancin Annalisa, Manfredini Fabio, Marangoni Silvia, Marchese Stefano, Marzano Bernardo, Mascioli Giosue', Masia Antonio Francesco, Mazzocchi Claudia, Mazzoni Maurizio Giovanni, Meco Massimo, Mela Federico, Meroni Roberta, Micciche' Eligio, Mingone Daniela, Molteni Mattia, Monti Cinzia, Moretti Antonio, Nerla Roberto, Nicoletti Alessandro, Nicoli Flavia, Orlandi Roberto, Paleologo Claudia, Passaretti Bruno Maria, Pedrigi Maria Cristina, Pesenti Nicola, Polidoro Roberto, Poloni Camillo Luca, Previtali Ilaria, Quartierini Giorgio, Rabbolini Giovanni, Re Chiara, Rea Bruna, Ricciardelli Gabriella, Rizzardi Giovanna, Ronzoni Annalisa, Roscitano Claudio, Rossi Daniela, Rota Stefano, Ruggiero Perrino Vincenzo, Salvini Piermario, Santoro Franco, Sauta Maria Grazia, Schifilliti Daniela, Scrofani Amalia, Selmi Carlo, Setti Lucia, Smarrelli Davide, Solinas Costantino, Spreafico Andrea Giovanni, Squadroni Michela, Stagno Maria Francesca, Stelian Edmond Jean, Stratta Gregorio, Tarantino Luca, Tessa Lorenzo, Testa Amidio, Testa Rosa Miranda, Traini Mariaemilia, Trovati Serena, Uccelli Fara Margerita Letizia, Usai Luca, Valenti Francesco, Vandenbulcke Filippo, Vavassori Vittorio Luigi, Vernile Laura, Viganò Luca, Villa Clara, Villa Giambattista, Villari Nicola, Vismara Alberto Carlo, Zambarbieri Giulia, Zanello Alessandro, Zerbini Graziano, Zotti Mariacamilla, Zumbo Aurora.

***COVICAT Study Group***

Gemma Castaño-Vinyals, Carlota Dobaño, Judith Garcia-Aymerich, Ximena Goldberg, Cathryn Tonne.

***Pa COVID-19 Study Group***

**Charité Universitätsmedizin Berlin, Berlin, Germany**

Stefan Hippenstiel, Sascha S. Haenel, Mirja Mittermaier, Fridolin Steinbeis, Tilman Lingscheid, Bettina Temmesfeld-Wollbrück, Martin Witzenrath, Thomas Zoller, Holger Müller-Redetzky, Alexander Uhrig, Daniel Grund, Christoph Ruwwe-Glösenkamp, Miriam S. Stegemann, Katrin M. Heim, Ralf H. Hübner, Bastian Opitz, Kai-Uwe Eckardt, Martin Möckel, Felix Balzer, Claudia Spies, Steffen Weber-Carstens, Frank Tacke, Chantip Dang-Heine, Michael Hummel, Georg Schwanitz, Uwe D. Behrens, Maria Rönnefarth, Sein Schmidt, Alexander Krannich Christof von Kalle, Linda Jürgens, Malte Kleinschmidt, Sophy Denker, Moritz Pfeiffer, Belén Millet-Pascual-Leone, Luisa Mrziglod, Felix Machleidt, Sebastian Albus, Felix Bremer, Jan-Moritz Doehn, Tim Andermann, Carmen Garcia, Philipp Knape, Philipp M. Krause, Liron Lechtenberg, Yaosi Li, Panagiotis Pergantis, Till Jacobi, Teresa Ritter, Berna Yedikat, Lennart Pfannkuch, Christian Zobel, Ute Kellermann, Susanne Fieberg, Laure Bosquillon de Jarcy, Anne Wetzel, Christoph Tabeling, Markus Brack, Moritz Müller-Plathe, Jan M. Kruse, Daniel Zickler, Andreas Edel, Britta Stier, Roland Körner, Nils B. Mueller, and Philipp, Paula Stubbemann, Nadine Olk, Willi M. Koch, Alexandra Horn, Katrin K. Stoyanova, Saskia Zvorc, Lucie Kretzler, Lil A. Meyer-Arndt, Linna Li, and Isabelle Wirsching, Denise Treue, Dana Briesemeister, Jenny Schlesinger, Birgit Sawitzki, Lara Bardtke, Kai Pohl, Philipp Georg, Daniel Wendisch, Anna L. Hiller, Sophie Brumhard, Marie Luisa Schmidt, Leonie Meiners, and Patricia Tscheak

***Covid-19 Aachen Study (COVAS)***

Paul Balfanz[1], Peter Boor[2], Ralf Hausmann[3], Hannah Kuhn[4], Susanne Isfort[5], Julia Carolin Stingl[3], Günther Schmalzing[3], Christiane K Kuhl[6], Rainer Röhrig[7], Gernot Marx[8], Stefan Uhlig[9], Edgar Dahl[10],[11], Dirk Müller-Wieland[12], Michael Dreher[13], Nikolaus Marx[12]

[1] Department of Cardiology, Angiology and Intensive Care Medicine, University Hospital RWTH Aachen, Germany.

[2] Institute of Pathology & Department of Nephrology, RWTH Aachen University, Aachen, Germany.

[3] Institute of Clinical Pharmacology, Uniklinik RWTH Aachen University, Aachen, Germany.

[4] Institute for Biology I, RWTH Aachen University, Aachen, Germany.

[5] Department of Hematology, Oncology, Hemostaseology and Stem Cell Transplantation, Medical School, RWTH Aachen University, Aachen, Germany.
[6] Department of Diagnostic and Interventional Radiology, RWTH Aachen University Hospital, Aachen, Germany.
[7] Institute of Medical Informatics, RWTH Aachen University Hospital, Aachen, Germany.
[8] Department of Intensive Care, Uniklinik RWTH Aachen, Aachen, Germany.
[9] Institute of Pharmacology and Toxicology, Medical Faculty Aachen, RWTH Aachen University, Aachen, Germany.
[10] Molecular Oncology Group, Institute of Pathology, Medical Faculty, RWTH Aachen University, Aachen, Germany.
[11] RWTH centralized Biomaterial Bank (RWTH cBMB) of the Medical Faculty, RWTH Aachen University, Aachen, Germany.
[12] Department of Internal Medicine I, RWTH Aachen University Hospital, Aachen, Germany.
[13] Department of Pneumology and Intensive Care Medicine, University Hospital Aachen.

***Norwegian SARS-CoV-2 Study group***

Synne Jenum (Department of Infectious diseases, Oslo University Hospital, Oslo, Norway), Børre Fevang (Section of Clinical Immunology and Infectious Diseases, Oslo University Hospital, Oslo, Norway), Birgitte Stiksrud (Department of Infectious diseases, Oslo University Hospital, Oslo, Norway), Else Quist-Paulsen (Department of Microbiology, Oslo University Hospital, Oslo, Norway), Simreen Kaur Johal, (Department of Microbiology, Oslo University Hospital, Oslo, Norway and Institute of Clinical Medicine, University of Oslo, Oslo, Norway), Anne Steffensen (Institute of Clinical Medicine, University of Oslo, Oslo, Norway and Department of Microbiology, Oslo University Hospital, Oslo, Norway), Liv Hesstvedt (Department of Infectious diseases, Oslo University Hospital, Oslo, Norway), Dag Henrik Reikvam (Department of Infectious diseases, Oslo University Hospital, Oslo, Norway and Institute of Clinical Medicine, University of Oslo, Oslo, Norway), Frank Pettersen (Department of Infectious diseases, Oslo University Hospital, Oslo, Norway), Vidar Ormaasen (Department of Infectious diseases, Oslo University Hospital, Oslo, Norway), Erik Egeland Christensen (Department of Infectious diseases, Oslo University Hospital, Oslo, Norway and Institute of Clinical Medicine, University of Oslo, Oslo, Norway), and Kjerstin Røstad (Department of Infectious diseases, Oslo University Hospital, Oslo, Norway), Linda Skeie (Department of Infectious diseases, Oslo University Hospital, Oslo, Norway), Marthe Jøntvedt Jørgensen (Department of Infectious diseases, Oslo University Hospital, Oslo, Norway and Institute of Clinical Medicine, University of Oslo, Oslo, Norway), Sarah Nur (Department of Infectious diseases, Oslo University Hospital, Oslo, Norway), Gry Klouman Bekken (Departmentt of Internal Medicine, Drammen Hospital, Vestre Viken Hospital Trust, Drammen, Norway), Anne Hermann, (Departmentt of Internal Medicine, Drammen Hospital, Vestre Viken Hospital Trust, Drammen, Norway), Hanne Opsand (Departmentt of Internal Medicine, Drammen Hospital, Vestre Viken Hospital Trust, Drammen, Norway), Bjørn Martin Woll (Departmentt of Internal Medicine, Drammen Hospital, Vestre Viken Hospital Trust, Drammen, Norway), Mette Bogen (Departmentt of Laboratory Medicine , Drammen Hospital, Vestre Viken Hospital Trust, Drammen, Norway), Cathrine Austad (Departmentt of Rheumatology, Drammen Hospital, Vestre Viken Hospital Trust, Drammen, Norway), Garth Daryl Tylden (Department of Microbiology and Infection Control, University Hospital of North Norway, Tromsø, Norway and Department of Medical Biology, Faculty of Health Sciences, UIT The Arctic University of Norway), Berit Gravrok (Department of Clinical Research, University Hospital of North Norway, Tromsø, Norway), Waleed Ghanima (Departments of Hemato-oncology and Research, Østfold Hospital Trust, Grålum, Norway and Department of Hematology, Oslo University Hospital and Institute of Clinical Medicine, University of Oslo, Oslo, Norway), Anne Marie Halstensen (Department of Research, Østfold Hospital Trust, Grålum, Norway), Jorunn Brynildsen (Department of Research, Østfold Hospital Trust, Grålum, Norway), Jeanette Aarem (Center for Laboratory Medicine, Østfold Hospital Trust, Grålum, Norway), Saad Aballi (Department of Infectious Diseases, Østfold Hospital Trust, Grålum, Norway), Siri Øverstad (Department of Internal Medicine, Østfold Hospital Trust, Grålum, Norway), Kristine Marie Aarberg Lund (Department of Infectious Diseases, Østfold Hospital Trust, Grålum, Norway), Åse-Berit Mathisen (Center for Laboratory Medicine, Østfold Hospital Trust, Grålum, Norway).

#### Supplementary Methods

##### *DNA extraction and SNP genotyping*

DNA extraction was either performed at the Institute of Clinical Molecular Biology (Christian-Albrechts-University of Kiel, Germany) or the respective study centers (**Supplementary Table 1a**). Genotyping for most samples was performed at the Institute of Clinical Molecular Biology. Samples from the BoSCO study were genotyped at the Genomics Department of Life&Brain Center, Bonn, Germany. Samples from the COMRI study were genotyped by the Genotyping laboratory of Institute for Molecular Medicine Finland FIMM Technology Centre, University of Helsinki, Finland. The ITM cohort was typed at the Regeneron Genetics Center, U.S.A, DNA extraction for this cohort was performed at the Institute of Clinical Molecular Biology.

##### *DNA extraction Institute of Clinical Molecular Biology*

DNA extraction was performed by the DNA laboratory of the Institute of Clinical Molecular Biology (Christian-Albrechts-University of Kiel, Germany) using a Chemagic 360 from PerkinElmer (Waltham, Massachusetts, U.S.) with the low volume kit cmg 1491 and the buffy coat kit cmg-714 (Chemagen, Baesweiler, Germany) according to the manufacturer's protocol. The Chemagen chemistry for DNA extraction from whole blood and buffy coat is based on the use of magnetic beads. Up to 400 µl whole blood or 300 µl buffy coat were used for isolation, depending on the shipped material of the different centers. In a first step, the cell lysis for protein degradation by protease is performed. The isolation of the DNA is achieved by capturing the DNA with polyvinyl alcohol magnetic beads (M-PVA Magnetic Beads). The DNA is bound to the surface coating of these beads. These beads, together with the bound DNA, are attracted by the magnetized metal rods, which can then transfer the DNA from one wash buffer to another. After deactivation of the electromagnet, the particles are resuspended in the solution. Finally, beads were transferred into 100-250 µl elution buffer, which inactivates the interaction between the beads and the DNA. The magnetic beads are removed, leaving the isolated DNA in suspension. The concentration was measured in a Dropsense96 (Unchained Labs, Pleasanton, CA, U.S.A.) spectrophotometer.

##### *DNA extraction for the GCAT cohort*

DNA extraction of the GCAT cohort(1) was performed by the GCAT laboratory of the Institute Germans Trias i Pujol (IGTP) using the automated ReliaPrep Large Volume HT gDNA Isolation System (Promega Biotech Ibérica S.L., Spain) following their own standard described procedures. The ReliaPrep chemistry for DNA extraction from whole blood and buffy coat is based on the use of magnetic beads with the HSM 2.0 Instrument automated on a robotic liquid-handling workstation. DNA yield and purity was determined by spectrophotometry using the Trinean Dropsense96 system (Unchained Labs, U.S.A.) and the integrity of a representative subsample was determined using the 2200 TapeStation System (Agilent Technologies Inc., U.S.A.)

##### *DNA extraction Genomics Department of Life&Brain Center, Bonn*

The Bonn study of COVID genetics (BoSCO) comprises two arms: (i) population-based recruitment of mild and asymptomatic infected participants, and (ii) clinic-based recruitment of moderate to severely affected individuals. Population-based participants provided saliva samples via the self-collection kit (Oragene OG-500, DNA Genotek), and DNA was extracted at the Department of Genomics, Life&Brain Center Bonn, using standard procedures as described by the manufacturer. For individuals recruited from wards and clinics, blood samples were retrieved during clinical care and DNA was extracted at each hospital, according to respective standard procedures. DNA was shipped to Bonn for subsequent genotyping.

##### *DNA extraction COMRI cohort*

DNA extraction was performed at the Institute for Virology (Technical University Munich, Munich, Germany) from 200 µl EDTA blood using the NucleoSpin Blood QuickPure Kit (Machery-Nagel, Düren, Germany) according to the manufacturer’s instructions. DNA concentrations were measured using NanoDrop One/OneC Microvolume UV Spectrophotometer (Thermo Fisher Scientific, Waltham, MA, U.S.A).

##### *Genotyping at the Institute of Clinical Molecular Biology*

Genotyping was conducted by the Institute of Clinical Molecular Biology’s DNA Laboratory and Genotyping Core Facilities (Christian-Albrechts-University of Kiel, Germany) employing Illumina’s (Illumina Inc., San Diego, U.S.) Global Screening Array-24 Multi Disease (GSA) Version 2.0 B1 and GSA Version 3.0. A1 following the Illumina(R) Infinium HTS Assay Auto 3-day Workflow (Document #15045738v0). In brief, initial DNA quantification, amplification and incubation for 24 hours was performed according to protocol on day 1. Next, we performed enzymatic DNA fragmentation, followed by 2-propanol precipitation, resuspension and overnight hybridization of DNA to the BeadChip on day 2. Finally, BeadChip washing removing unhybridized and non-specifically hybridized DNA, extension adding labeled nucleotides to extend primers hybridized to the sample, staining of primers and final imaging using the Infinium LCG scan setting was performed following the manufacturer's protocol on day 3. The genome-wide content of the GSA was selected by the vendor for high imputation accuracy at minor allele frequencies (MAF) >1% across all twenty-six 1,000 Genomes Project populations(2). The clinical research content includes 712,189 (GSA Version 2.0) variants or 730,059 variants (GSA Version 3.0) with established disease associations, relevant pharmacogenomics markers, and curated exonic content based on ClinVar, NHGRI, PharmGKB, and ExAC databases.

***Genotyping at the Life&Brain Center, Bonn, Germany***

Genotyping of all samples of the BoSCO study was conducted using Illumina's (Illumina Inc., San Diego, U.S.) Global Screening Array-24 Multi Disease (GSA) Version 3.0.

***Genotyping at the Regeneron Genetics Center***

Genotyping was conducted employing Illumina's (Illumina Inc., San Diego, U.S.) Global Screening Array-24 Multi Disease (GSA) Version 1.0.

***Genotyping at the Genotyping laboratory of Institute for Molecular Medicine Finland FIMM Technology Centre, University of Helsinki***

Genotyping was conducted employing Illumina's (Illumina Inc., San Diego, U.S.) Global Screening Array-24 Multi Disease (GSA) Version 3.0. DNA quantity and/or quality the samples were measured with Qubit (ThermoFisher) and Nanodrop (ThermoFisher) and sometimes run on FlashGel™ (Lonza) to verify the concentration and detect possible contaminants or fragmentation of the DNA. Processing of the samples followed the infinium-hts-assay-reference-guide-15045738-04. The samples were processed either by hand or using Viaflo 96 (Integra) and Tecan/Infinium automated system pipetting platforms. The arrays were scanned with Illumina iScan instrument.

##### *Genotype calling, quality control and imputation*

##### *Genotype calling*

Initial genotype calling extracting GSA genotyped data from intensity data files was performed with the Illumina GenomeStudio Version 2.0 software with the cluster definition files (GSAMD24v2-0_20024620_A1-762Samples-LifeBrain (GSA Version 2.0), GSAMD-24v3-0-EA_200034606_A1 (GSA Version 3.0) and GSAMD-24v1-0-A_4349HNR_Samples (GSA vs. 1.0). The calling of Y-Chromosomal SNPs was performed on males only. Finally, we had 712,189 SNPs before genotype quality control (QC) on the GSA Version 2.0, 730,059 SNPs before QC on the GSA Version 3.0 and 700,078 SNPs before QC on the GSA Version 1.0 After genotyping a total of 4,067 German/Austrian (449 cases/3,618 controls), 7,347 Spanish (2,795 cases/4,552 controls), 415 Norwegian (127 cases/288 controls) and 7,114 Italian (1,857 cases/5,247 controls) were available with non-missing core-phenotype (COVID-19 case-control status, respiratory support status and sex) information. For individuals with missing sex information, sex was inferred from the genotypic sex if possible.

##### *SNP and sample quality control and principal component analysis*

Based on initial genotype data, we removed samples with <90% call rate using PLINK.(3, 4) We additionally removed individuals with non-matching genotypic and phenotypic sex. After genotype calling, a QC procedure was carried out for the Spanish, Italian, Norwegian and German/Austrian case-controls GWAS datasets respectively. Variants that had >2% missing data, a MAF<0.1% in disease sets or in controls, different missing genotype rates in affected and unaffected individuals ($P_{Fisher}<10^{-5}$) or deviated from Hardy-Weinberg equilibrium (with a false discovery rate (FDR) threshold of $10^{-5}$ in controls) (a) across the entire collection with at most one batch being removed or (b) falling below in two single batches, were excluded. Samples that had overall increased/decreased heterozygosity rates (i.e. ±5 SD away from the sample mean) were removed. For robust duplicate/relatedness testing (IBS/IBD estimation) and population structure analysis, we used a linkage disequilibrium (LD)-pruned subset of SNPs on the basis of a set of independent (MAF≥5%) SNPs excluding X- and Y-chromosomes, SNPs in LD (leaving no pairs with $r^2>0.2$), and 11 high-LD regions as described by Price *et al.*(5) Pairwise percentage IBD values were computed using PLINK. By definition, Z0: P(IBD=0), Z1: P(IBD=1), Z2: P(IBD=2), Z0+Z1+Z2=1, and PI_HAT: P(IBD=2) + 0.5 * P(IBD=1) (proportion IBD). One individual (the one showing greater missingness) from each pair with PI_HAT>0.1875 was removed. A value of 0.1875 (proportion IBD) corresponds to a theoretical relationship of halfway between the expected IBD for third- and second-degree relatives. To identify ancestry outliers, i.e. subjects of non-European ancestry, we performed principal component analysis (PCA) for the remaining QCed cases and controls including reference samples from the 1,000 Genomes Phase 3 reference panel.(2) We used the PCA method, as implemented in FlashPCA(6) on an LD-pruned subset of SNPs (see text above). Ancestry outliers not matching European populations were removed (**Supplementary Figures 1a-f**). After QC, PCA revealed no non-European ancestry outliers (**Supplementary Figures 1e-h)** when performing PCA.

***SNP genotype imputation***

The QCed Italian, Spanish, Norwegian and German GWAS datasets comprised 1,563 Italian COVID-19 cases, 4,759 Italian controls, 2,174 Spanish COVID-19 cases, 4,406 Spanish controls, 81 Norwegian cases, 283 Norwegian controls, 336 German COVID-19 cases and 3,303 German controls, and contained 567,131 (Italy), 564,856 (Spain), 525,836 (Norway) and 476,562 (Germany/Austria) variants after QC and filtering of SNPs with alleles AT or CG (the latter often leading to strand issues during imputation). Genotype imputation was conducted for chromosomes 1-22 and X data using the novel TOPMed Freeze 5 on genome build GRCh38 and the Michigan Imputation Server.(7) We provided the input data in "vcf.gz" format as GRCh38 build. We used the offered population panel "ALL" and applied the server-side option to filter by an imputation $R^2$ with threshold 0.1. The final imputed results contained 80,794,511 variants in the Italian dataset, 75,346,562 variants in the Spanish dataset, 20,195,513 variants in the Norwegian dataset and 53,164,135 variants in the German/Austrian dataset after TOPMed imputation. For the imputation of the X chromosome, we coded males as diploid in the non-pseudoautosomal (non-PAR) region. After quality control and imputation using TOPMed in total 8,910,172 variants were included for the Italian panel, 9,089,877 variants for the Spanish panel, 8,841,609 variants for the Norwegian panel and 9,019,898 variants for the German/Austrian panel with post imputation $R^2 \geq 0.6$ and MAF≥1%.

***Imputation of the ~0.9-Mb inversion polymorphism at 17q21.31***

In 2005, Stefansson and colleagues discovered a 900-kb inversion polymorphism at 17q21.31, a region that contains several genes, including those encoding corticotropin releasing hormone receptor 1 (*CRHR1*) and microtubule-associated protein tau (*MAPT*).(8) Chromosomes with the inverted segment in different orientations represent two distinct lineages, H1 and H2, that have diverged for up to 3 million years and show no evidence of recombination.(8) For the Italian, Spanish, Norwegian and German/Austrian GWAS discovery cohorts we inferred the 17q21.31 inversion status (H1 or H2) with IMPUTE v2.3.2(9) using genotype information and an imputation reference panel consisting of 109 individuals (from different continents [EUR, EAS, AFR]) from the 1,000 Genomes Project Phase 3(2), for which 17q21.31 inversion genotypes, obtained experimentally by FISH(10, 11) and droplet digital PCR(12), as well as SNP genotype data are available for the region of the inversion. Imputed inversion genotypes were determined according to the highest posterior probability and were further confirmed by examining consensus genotypes of known inversion tag SNPs in perfect LD ($r^2$=1) with the inversion in the imputation reference panel. H1 and H2 alleles were coded according to an additive model as 0,1 and 2.

We additionally imputed the 17q21.31 inversion for the COVID-19 HGI release 5 A2 and B2 analyses. Here, the inversion P-value was imputed from summary statistics using Fast and accurate P-value Imputation for genome-wide association study (FAPI).(13) The odds ratio (OR) and 95% confidence intervals (CIs) were estimated from the SNP rs62061809, which is in perfect LD with the inversion ($r^2$=1).

***Genome-wide and candidate-based association analysis***

Using genome-wide SNP information, we performed different types of statistical analyses as specified below. If not stated otherwise, all analyses were performed as follows: case-control allele-dose association tests of the genotyped and imputed SNPs in the Italian, Spanish, Norwegian and German/Austrian panels were performed separately using SAIGE (version 0.43.2).(14) Using age, biological sex, age*age, biological sex*age and the first 10 principal components (PCs) from PCA as covariates, we performed a logistic association analysis assuming additive effects. For chromosome X, association analyses were performed with SAIGE parameters --is_rewrite_XnonPAR_forMales=TRUE, the XPAR region given as --X_PARregion "10001-2781479,155701383-156030895" and --sampleFile_male=males.txt, with ids of males saved in males.txt, to account for sex in the analysis. For each of the following analyses, sample numbers are given in **Supplementary Table 1e**.

***Cohort-specific analyses***

Ia. *Genome-wide association analysis for first GWAS discovery cohorts*

For each individual case-control dataset, COVID-19 patients with respiratory failure (cases, respiratory support status 1-4) were compared with population controls (negative or unknown COVID-19 status). Statistical testing was performed as:

Case/Control ~ SNP + age + sex + age*age + age*sex + PC1 + PC2 + PC3 +PC4 + PC5 + PC6 + PC7 + PC8 + PC9 + PC10

Ib. *Genome-wide association analysis for second GWAS discovery cohorts*
For each individual case-control dataset, COVID-19 patients with respiratory failure (cases, respiratory support status 2-4) were compared with population controls (negative or unknown COVID-19 status). Here the Norwegian dataset was omitted due to a small case sample size ($N_{Cases} < 50$).

Ic. *Genome-wide association analysis for first GWAS discovery cohorts, excluding overlap with COVID-19 HGI release 5 B2 data.*
We performed the Ia analysis as above, omitting 1,700 (775 cases/925 controls) Spanish and 2,090 (835 cases/1,255 controls) Italian individuals as well as all German individuals from the BoSCO study overlapping with the B2 analysis of the COVID-19 HGI release 5 data.

Id. *Genome-wide association analysis for second GWAS discovery cohorts, excluding overlap with COVID-19 HGI release 5 A2 data.*
We performed the Ib analysis as above omitting 1,227 (302 cases/925 controls) Spanish and 1,953 (698 controls/1,255 cases) Italian individuals overlapping with the A2 analysis of the COVID-19 HGI release 5 data.

***Meta-analysis of cohort-specific summary statistics.***
The resulting summary statistics datasets (individual cohort post-imputation $R^2 \geq 0.6$) were meta-analyzed using fixed-effect meta-analysis based on METAL's (https://genome.sph.umich.edu/wiki/METAL_Documentation) inverse-variance weighted approach.(15) Post meta-analysis, all analyses were filtered for MAF≥1% and at least N=2 studies.

Meta-analyses were conducted as follows:
IIa) *Meta-analysis of Italian, Spanish, Norwegian and German/Austrian cohorts from Ia*
IIb) *Meta-analysis of Italian, Spanish and German/Austrian cohorts from Ib*
IIc) *Meta-analysis of Italian, Spanish, Norwegian and German/Austrian cohorts from Ic and COVID-19 HGI genetics consortium B2 release 5 data (COVID19_HGI_B2_ALL_leave_23andme_20210107)*

IId) *Meta-analysis of Italian, Spanish and German/Austrian cohorts from Id and COVID-19 HGI genetics consortium A2 release 5 data (COVID19_HGI_A2_ALL_leave_23andme_ 20210107)*

***Bayesian fine-mapping analysis***

Statistical fine-mapping analysis was conducted using FINEMAP(16) Version 1.4 for each of the loci of interest to calculate the posterior inclusion probability (PIP) for each lead SNP and every other SNP within 250kb flanking regions. In the case where the association signal extended over a larger region, as in the case of 17q21.21, the region was expanded to include this. FINEMAP determined the 95% credible set of SNPs assuming a single causal variant using shotgun stochastic search (--n-causal-snps 1 --sss), i.e. the minimum set of variants containing the causal variant with ≥ 95% certainty. The union of the genotypes of the Italian, Spanish, Norwegian and German/Austrian cohorts was used as LD reference population.

***Meta-Analysis Model-based Assessment of replicability***

We tested the replicability of our candidate variants from the meta-analysis using the Meta-Analysis Model-based Assessment of replicability(17) (MAMBA) method, as single outlier studies can drive a false positive meta-analysis association. MAMBA takes the genetic effects and standard deviations from participating studies to test each SNP for a true non-zero effect. This is achieved by fitting a two-level mixture model to genome-wide LD-pruned SNPs that reduces to a fixed effect meta-analysis in absence of outliers, in which case it has similar power. Since this is a likelihood model, the result is a posterior probability of replicability (PPR) that the SNP has a non-zero replicable effect. If the PPR is low, MAMBA tests for excess in variation of effects sizes and outputs the probability that each study is an outlier. For each of the first and second analyses we used the lead variants and a genome-wide set of LD-pruned SNPs (PLINK v1.90b6.16 64-bit, --indep-pairwise 500kb 1 0.1)(3, 4) from each participating study for the algorithm to estimate the null distribution. Suggestive variants and LD-pruned background SNPs are entered as a single table and the algorithm has no prior knowledge of which SNPs are considered significant. MAMBA was run with default settings and always terminated before 10,000 iterations.

***Association analysis and meta-analyses of candidate loci***

For candidate loci, we additionally performed stratified analyses based on disease severity, sex and age as detailed below.

III. Association analysis of severity

For each individual case dataset, COVID-19 patients were stratified based on their respiratory support status, which we used to define a new case-control status (control = respiratory support status 1; case = respiratory support status 2-4. Statistical testing was performed as:

Case only (respiratory support status=1 vs. respiratory support status=2-4) ~ SNP + age + sex + age*age + age*sex + PC1 + PC2 + PC3 +PC4 + PC5 + PC6 + PC7 + PC8 + PC9 + PC10**.**

IV. *Sex-stratified association analysis*

For each individual case-control dataset, COVID-19 patients with respiratory failure (cases) were compared with population controls (negative or unknown COVID-19 status). Sex-stratified analysis were performed for males and females separately. The resulting case-control numbers are shown in **Supplementary Table 1e**.

Sex-stratified analyses were performed according to

Case/Control ~ SNP + age + age*age + PC1 + PC2 + PC3 +PC4 + PC5 + PC6 + PC7 + PC8 + PC9 + PC10

V. *Age-stratified association analysis*

Individuals from analysis Ia/Ib were binned into age groups 0-20, 21-40, 41-60, 61-80 and >80 to calculate age-specific single-study and meta-analysis allele frequencies, ORs and P-values. Since sample numbers in the categories 0-20 and >80 were small (**Supplementary Table 1e**), association analyses was only conducted on ages 41-60 and 61-80. We additionally analysed age categories <=60 vs >60 years.

Age-stratified analyses were performed according to

Case/Control ~ SNP + sex + PC1 + PC2 + PC3 +PC4 + PC5 + PC6 + PC7 + PC8 + PC9 + PC10.

VI. Analysis of comorbidities

For each individual case dataset, COVID-19 patients were stratified based on whether they suffered from hypertension, coronary artery disease or diabetes. For each comorbidity we defined a new case/control status, which control individuals being COVID-19 patients not suffering from the respective comorbidity and case individuals being COVID-19 patients suffering from the respective comorbidity.

Analyses were carried out according to

Case only (comorbidity=”no” vs. comorbidity =”yes”) ~ SNP + age + sex + age*age + age*sex + PC1 + PC2 + PC3 +PC4 + PC5 + PC6 + PC7 + PC8 + PC9 + PC10**.**

All analyses on single loci were conducted using logistic regression in R (Version 3.6.1/3.6.2), according to the analysis plan described for analyses I-II. Meta-analyses were conducted using the metafor(18) package, weighted by the variance of the estimates. For the meta-analyses only cohorts with a $N_{Cases}$ & $N_{Controls}$ > 50 were considered.

##### ***Functional characterization of genome-wide significant lead variants and associated candidate genes***

***Phenome-wide association studies (PheWAS)***

We queried the NHGRI-EBI GWAS Catalog curated collection of established genome-wide significant disease/trait associations (http://www.ebi.ac.uk/gwas/; release 2020-12-02) for GWAS hits in high LD ($r^2 > 0.9$) with the inversion or rs1405655 (19q13.33) and a P-value < $10^{-7}$. Since each GWAS study is focused on populations from different origins, the LD patterns employed to evaluate the association between the inversion and GWAS signals were based on individuals with the corresponding ancestry or the closest one available from our inversion imputation panel (of 109 experimentally genotyped individuals), whereas the global LD was selected if populations from underrepresented ancestries were studied. 27 proxy variants were used for r1405655 and 2,904 proxy variants were used for the inversion.

***Tissue-specific expression and splicing quantitative trait loci (eQTL, sQTL)***

eQTLs and sQTLs were evaluated using publicly available data from the GTEx Project (GTEx Analysis Release V8)(19) in 49 different human tissues. Here we extracted analysis estimates (normalized effect score (NES)) and P-values of the two variants of interest: rs62055540 (tag SNP for the 17q21.31 inversion) and rs1405655 (19q13.33) as well as 27 proxy ($r^2$>0.9) variants for rs1405655 and 2,904 proxy variants for the inversion ($r^2$>0.9). LD was calculated based on the 1000 Genomes population European(2) population only, to account for the ethnic bias of GTEx donors. Based on these P-values we additionally investigated whether the two variants themselves or any proxy variants were the lead-SNP (variant with the lowest nominal  P-value in a tissue/gene) for an expression and splicing change.

***Selection and definition of candidate genes at 17q21.31 and 19q13.33***

To identify candidate genes most likely to play a causative role at 17q21.31 and 19q13.33, all protein-coding genes that are located within locus boundaries or that are candidates based on the lead cis-eQTL (eGenes) or sQTL (sGenes) association analysis were chosen. More precisely, the boundaries for the 19q13.33 locus were defined by Bayesian fine mapping (GRCh38: chr19:50344768-50379362) while extended boundaries (GRCh38: chr17:45495836-46707123)(20) were used for the 17q21.21.31 locus, since it lies within a ~0.9Mb inversion region of high LD that affects expression and splicing of numerous genes in GTEx.(19) To retrieve candidate genes that overlap the boundaries, GENCODE v36(21) annotations for genome build GRCh38 were used. A complete list of candidate protein-coding genes from both loci is provided in **Supplementary Table 20**.

***Gene expression analysis for candidate genes from genome-wide significant susceptibility loci***

Publicly available bulk tissue and immune cell type RNA-seq data for all available candidate genes were retrieved from GTEx v8(19) and from the Expression Atlas(22) (BLUEPRINT consortium data [accession E-MTAB-3827](23)) portals, respectively. Gene-level expression values (transcripts per million, TPM) by tissue or by cell type were obtained as median-summarized in the case of the GTEx data and as mean-summarized in the case of the BLUEPRINT data. The summarized TPM values were centered gene-wise, and z-score scaled for visualization using the ggplot2 R-package. The single-cell RNA-seq (scRNA-seq) data in COVID-19 relevant tissues from non-diseased individuals, such as lung and upper airways(24) or brain(25) were obtained from the COVID-19 Cell Atlas.(26) The pre-processed and cell type annotated scRNA-seq datasets were retrieved as AnnData objects in .h5ad format files. Log-normalized average expression values of available candidate genes by cell type were visualized using the scanpy v1.4.6 package.(27) For gene expression analysis of candidate genes in SARS-CoV-2 infected brain organoids, pre-processed and cell-annotated scRNA-seq data were obtained upon request from Song *et al.*(28) Differential gene expression analysis of scRNA-seq data was performed using the R package MAST.(29) More precisely, hurdle models were used to evaluate differentially expressed genes in each brain organoid cell type (neural progenitors, interneurons, neurons, and cortical neurons) comparing SARS-CoV-2 infected and mock (non-infected) cells. The models were fitted using the condition, sample identification, number of detected genes (centered) and total counts of SARS-CoV-2 transcripts (centered) as covariates, thus adjusting for the cellular detection rate, batch effects and viral load. Genes with PFDR<0.01 and absolute value of log2 fold change >0.1 were considered as significantly differentially expressed. Finally, the status, log2 fold change and P-values of candidate gene differential expression in COVID-19 infected lung cells were obtained from pseudo-bulk differential expression analysis performed by Delorey *et al.*(30)

***Mendelian Randomization analysis***

Mendelian Randomization (MR) analysis uses genetic variants, which are expected to be independent of confounding factors, as instrumental variables to test for causal relationships between various human traits and severe COVID-19 with respiratory failure. For this purpose, genetic variants from susceptibility loci 19q13.33 and 17q21.31 were subjected to two-sample summary data-based MR using the TwoSampleMR package(31) and the MRC IEU OpenGWAS database.(32) Whole-blood *cis*-eQTL results for the genes in the susceptibility loci were retrieved from the eqtl-a(33) dataset and used as 'exposure' to assess their effect on the COVID-19 summary statistics meta-analysis results which were used as 'outcome'. The analysis workflow followed the MR approach as described by Zhu *et al.*(34) Briefly, exposure trait summary data were filtered to retain only instrument variables with $P_{eQTL} \leq 1.6 \times 10^{-3}$ (equivalent to $X^2 > 10$) and MAF ≥%1. If no instrument in the analyzed trait fulfilled this criterion in the candidate loci, the trait was discarded. After harmonization of exposure and outcome instruments, Wald test statistics were calculated for all 3,408 variants within the candidate locus boundaries with summary statistics for the outcome (severe COVID-19 with respiratory failure) and exposure (*cis*-eQTL traits). The variant with the smallest P-value was defined as the lead variant. Based on the number of SNPs from the candidate loci (n=3,408), P =0.05/3408 =1.467 x $10^{-5}$ was defined as the study-wide significance threshold. For traits meeting study-wide significance, we further applied the HEIDI-outlier-detection approach from Zhu *et al.* to discriminate single genetic instruments (variants) that appear to have pleiotropic effects on both risk factor (i.e. traits that have been investigated here) and disease from heterogeneous signals, indicating effects that cannot only be attributed to the interaction of gene expression and severe COVID-19.

***17q21.31 inversion-related effects with respect to gene expression in infection-stimulated CD14+ monocytes***

To investigate a possible effect of the 17q21.31 inversion with respect to an altered condition of infection, we used publicly available RNA-seq and high-density genome-wide SNP genotyping data (accession EGAS:00001001895) from unstimulated and stimulated CD14+

monocytes treated with different bacterial and viral stimuli for a total of 100 individuals of European and 100 individuals of African origin, with a total of 970 expression experiments: 200 unstimulated, 184 bacterial lipopolysaccharide, 196 Pam3CSK4, 191 R848, and 199 human influenza A virus.(35) As before, inversion genotypes (H1 or H2) were imputed using IMPUTE v2.3.3(9) from genome-wide SNP genotyping for each individual, using our inversion imputation panel of 109 experimentally genotyped individuals from the 1000 Genome Population reference.(36) Next, we performed pseudoalignment of RNA-seq read data using Kallisto v0.46.0(37) and a reference transcriptome index derived from GENCODE v36(21) to quantify overall transcript abundance. Individual gene-level abundance for each of the 970 expression experiments was calculated as the sum of estimated counts for all transcripts of a gene using the tximport package(38) and transformed to TPM.

To detect eQTL associations at gene level, for each condition we filtered out non-expressed genes (0 TPMs in >75% of samples) and ran linear regression models of gene expression (TPM) by genotype using QTLtools Version 1.1.(39) The models were adjusted by the first three PCs from genotype data and a set of components calculated from expression TPMs to capture technical confounding factors. These factors were estimated using PCA and by taking up to 50 expression-derived PCs in intervals of 5. The number of components used was chosen to maximize the number of eQTL associations. The analysis was done including the 17q21.31 inversion and neighboring variants located within 1 Mb from the gene transcription start site of each selected gene to estimate the contribution of each polymorphism to expression variation and identify lead eQTLs. To adjust nominal P-values for multiple testing, we adopted the permutation approach implemented in QTLtools and significant gene-variant pairs were determined by the R/qvalue package with FDR of 5%.(40) For transcript level data, we carried out the same analysis as above using the relative expression of each gene alternative transcript isoforms as the testing phenotype for the association to detect changes in the splicing patterns.

##### *HLA locus fine mapping and association analysis*

##### *HLA allele, amino acid and SNP imputation*

Imputation of alleles at the histocompatibility leukocyte antigen (HLA) region was performed for the classical HLA class I loci HLA-*A*, -*B*, -*C*, the class II loci HLA-*DQA1*, -*DQB1, -DPA1, -DPB1, -DRB1* and the non-classical HLA class I locus HLA-*E* at 2-field G group resolution from quality-controlled SNP genotype data. Here, we extracted pre-imputation SNP genotypes from the extended HLA region (chromosome 6: 29-34Mb) and used them as input for HLA genotype prediction with the random-forest based machine learning tool HIBAG (version 1.20.0).(41–43) To optimally cover the genetic ancestry of all analyzed cohorts, we used different imputation reference panels for the Italian, Spanish, Norwegian and German/Austrian data: (1) For the Spanish and Italian dataset, we used a HIBAG model specifically trained for this study (details below). (2) As an imputation reference for all HLA loci except HLA-*E* within the Norwegian and German/Austrian dataset, we used the previously trained and publicly available multi-ethnic HLARES models for the "Illumina Infinium Global Screening Array v2.0" (available at: https://hibag.s3.amazonaws.com/index.html), For imputation of the HLA-*E* locus in the German/Austrian and Norwegian dataset, we used model (1). (3) Since neither of the models published by Zheng *et al.*(41) nor our new model allowed for imputation of *HLA-DPA1*, but both an alpha and a beta chain are necessary for the prediction of peptide binding affinity (see below), we imputed HLA-*DPA1* alleles using the multi-ethnic reference model published by Degenhardt *et al.*(42) which was modified to additionally use the variants available on the GSA. We additionally derived amino acid and additional SNP imputation and defined marginal posterior probabilities for single HLA alleles across all predicted HLA genotypes as described in Degenhardt *et al.* (43)

##### *Construction of an imputation panel for the Spanish and Italian cohorts*

For the Spanish and Italian datasets, we first constructed a specifically tailored HLA imputation reference panel, using available next-generation sequencing-(NGS) based HLA allele typing for a subset of 2,576 samples (836 cases from Spain; 838 cases and 897 controls from Italy) and quality-controlled SNP genotype information from the same individuals. The HIBAG model was trained using 100 individual classifiers for each of the loci *HLA-A, -B, -C, -DRB1, -DQA1, -DQB1, -DPB1 and -E*. For details on the NGS typing refer to Ellinghaus *et al.*(44)

##### *SNP- and amino-acid-wide association study*

Due to the high SNP density in the HLA region and excessive allelic variation at the classical HLA loci, we performed a dedicated HLA locus fine mapping analysis based on imputed SNP, amino acid and classical HLA allele data. Association analyses were performed as described in analyses I-III. Imputed SNPs, amino acids and classical HLA alleles with MAF<0.05% were filtered out; classical HLA alleles with a marginal posterior probability from imputation <0.3 were further excluded. Association analyses were conducted for all four cohorts separately (Italy, Spain, Norway and Germany/Austria) using PLINK's logistic framework; fixed-effects meta-analysis was performed using METAL as described in analyses IIa/IIb).(3, 4, 15) Only alleles present in at least two of the cohorts (after filtering) were included in the meta-analysis.

##### *Peptidome-wide association study (PepWAS)*

To screen for disease-relevant peptides that may present a possible functional link between severe COVID-19 and variation at classical HLA loci, a peptidome-wide association study(45) was conducted. Here, the goal of the PepWAS approach was to identify associations between HLA-presented viral peptides and COVID-19 susceptibility and severity by examining similarities and differences in peptide binding affinities among HLA alleles across samples. The PepWAS approach has the potential to identify HLA-presented peptides that are more abundantly or less abundantly presented in cases compared to controls, potentially conferring risk or protection. Briefly, the reference proteome of SARS-CoV-2 (UP000464024 [accessed on 12/17/20]) was downloaded from UniProt.(46) For HLA class I and class II alleles, the proteome was digested into 9mer and 15mer peptides, respectively, using a sliding window approach with a step-size of one amino acid. Next, the binding of the generated peptides to HLA class I and class II alleles was predicted using NetMHCpan-4.1 and NetMHCIIpan-4.0(47), respectively. For the main analysis, we focused on strong binders, i.e. peptides having a predicted affinity percentile rank less than 0.5 for HLA-I and less than 1 for HLA-II. We also repeated the analysis for HLA class I with a more stringent threshold (rank less than 0.1 and absolute affinity values below 50nM). We further repeated the analyses with two other peptide binding prediction algorithms, MHCflurry(48) for HLA class I and MHCnuggets(49) for HLA class II alleles. Two different COVID-19 phenotype contrasts were tested for association with HLA-presented peptides, the disease risk and severity as defined in the main analysis. We used the same logistic regression models as for the genetic association (GWAS) analysis, including the full set of covariates. As a threshold for statistical significance, we applied

Bonferroni correction using the number of peptides bound by at least one of the HLA alleles of a given HLA locus.

***Analysis of quantitative HLA parameters***

As quantitative aspects of individual HLA variability (e.g. heterozygosity) are known to associate with the risk or severity of infectious diseases(50), we investigated the potential role of several such quantitative parameters, also following up on previous analyses described in Ellinghaus *et al.*(44)

1. We calculated allele numbers across the classical class I (HLA-*A,* -*B*, -*C*) and class II (HLA-*DRB1*, -*DQB1*) loci as a measure of multilocus heterozygosity (ranging from 3 and 2 alleles for complete homozygosity to 6 and 4 for complete heterozygosity, respectively).
2. Additionally, a more nuanced measure of allelic variability, the amino acid sequence divergence among alleles, was calculated for each of the five HLA loci separately and across all three class I or two class II loci, using the GranthamDist tool. The rationale behind this is described in detail by Pierini & Lenz.(51)
3. Another quantitative compound measure of HLA variability is the total number of peptides bound by an individual's HLA variants. This has for instance been shown to predict control of HIV replication.(52) We therefore used the HLA-peptide binding prediction algorithms NetMHCpan-4.1 and NetMHCIIpan-4.0 for HLA class I and class II alleles, respectively.(47) We used the same reference proteome as for the PepWAS analysis to infer all possible potentially relevant peptides (9mers for class I and 15mers for class II). Default %Rank_EL thresholds as defined in Reynisson *et al.*(47) were used to define strong (0.5% for NetMHCpan and 2% for NetMHCIIpan) and both weak and strong (2% for NetMHCpan and 10% for NetMHCIIpan) binders. The total number of bound peptides per individual was calculated for each locus as well as for all class I and class II variants together.
4. According to Grifoni *et al.*(53) CD4+ T-cell responses concentrate on M, N and Spike proteins and CD8+ T cell responses on M, NSP6 and Spike proteins. Therefore, we also calculated specifically the number of bound peptides for these subsets of proteins (for class I alleles from M, NSP6 and Spike proteins and for class II alleles from M, N and Spike). Finally, the calculations were repeated by using only the Spike protein, given its importance in the transmission of SARS-CoV-2.

Association between these compound parameters of genetic variability and disease risk (cases vs. general population; analogous to analysis Ia) as well as disease severity (analogous to analysis III), i.e. substituting the SNP dosage by the HLA parameter in Ia and III. We also tested for a quadratic relationship using parameter$^2$ as an additional variable. We performed a combined analysis of all data according to Ia and III, including cohort ID (Italy, Spain, Norway and Germany/Austria) as a covariate to account for differences between the datasets.

5. Eventually, we also converted HLA-*B* alleles into supertypes, following Sidney *et al.*(54), and tested their association with disease risk and severity in the same way as for the other parameters.
6. KKDKKKKAD and SSRSSSRSR, both originate from the Nucleocapsid (N) protein and present exact matches to human self-peptides (see below). Therefore we investigated

As an analysis at the population level, we also tested whether the association effect obtained from the HLA fine mapping (see above) correlated with the predicted number of bound peptides across allele of a given HLA locus, as it has been observed for HIV-1 control where HLA-*B* alleles that bound more HIV-1 peptides were associated with reduced viral load.(45)

***HLA-presentation of shared peptides ('molecular mimicry')***

Cross-reactivity of T cell-mediated immunity from viral epitopes to human self-epitopes has been considered as a potential trigger of excess inflammatory responses associated with severe forms of COVID-19. Such cross-reactivity would be most likely in the case of viral peptides that share protein sequence similarity with human self-peptides ('molecular mimicry'). We therefore compared all possible 9mer (for HLA class I) and 15mer (for HLA class II) peptides between the SARS-CoV-2 reference proteome (accession UP000464024) and the human reference proteome (accession UP000005640) and identified identical matches. We then used the same peptide binding prediction approach as above to predict the presentation of these peptides in cases and controls given the individual HLA genotypes, to test for associations between HLA-presented shared peptides and COVID-19 risk and severity.

We also considered the possibility that molecular mimicry might play a role in COVID-19 severity independent of HLA variation. We therefore identified SARS-CoV-2 peptides that were predicted to be bound by all common alleles (frequency>1% in current dataset) of a given HLA class I locus (using the NetMHCpan tool as above) and compared them to proteins over-expressed in the human lung (as the main organ experiencing COVID-19-related inflammation). These lung-expressed proteins were retrieved from the Human Protein Atlas(55) (n=231 as of 12/2020). The SARS-CoV-2 peptides were blasted against human lung proteins using command line blastp tool(56) and fragments of length ≥8 amino acids and identity ≥60% were selected as regions of molecular mimicry between COVID-19 and human lung proteins.

Gene set enrichment analysis (GSEA) for the genes exhibiting peptide similarity with SARS-CoV-2 (as defined above) was performed for Gene Ontology (GO) terms (biological processes, molecular functions and cellular compartments) taken from The Molecular Signatures Database Version 7.1.(57, 58) Only terms with at least 3 genes in the present gene set were considered. We calculated enrichment (ORs) using Fisher's exact test against the background of all human genes (minus the present gene set). Statistical significance of enrichment was derived from 100,000 permutations. In each permutation, as many genes as in the present gene set were sampled from background human genes, and their overlap with the given pathway was calculated. The P-value was taken as the number of times the sampled gene sets had equal or higher overlap than the original gene set. The empirical P values were finally Bonferroni corrected for the total number of tested pathways.

***Association analysis of Y-chromosomal haplogroups***

***Calling of genotypes***

We produced high quality Y-chromosome genotypes by manually calling and visually inspecting Y-chromosome SNPs in the male fraction of the cohorts only. We used Illumina's Genome Studio for clustering of genotyping intensities. Heterozygous calls were set to missing.

***Y-chromosome haplogroup characterization by genotyping***

By using 22 Y-chromosomal SNP genotypes extracted from the raw genotype data, we performed a SNP based analysis of haplogroups E, G, H, I, J, L, N, O, Q, R and T including analysis of sub-clades of G, I, J, and R.

SNPs were identified to cover the most common haplogroups in the four different populations (Italian, Spanish, Norwegian and German/Austrian) harmonized across the different genotyping arrays (**Online Methods**). We extracted the genotypes of 22 SNPs: rs2032654, rs13447378, rs34134567, rs2032673, rs2032597, rs13447352, rs3911, rs34442126, rs16981290, rs8179021, rs2032658, rs20320, rs9341296, rs35547782, rs9341313, rs2032604, rs3908, rs9786184, rs9786194, rs16981293, rs11799226 and rs1236440. We inferred the haplogroup status based on these SNPs as described below.

*Level 0 haplogroups*

We used SNP rs2032654 (phylogenetic marker M215) specific for subclade E1b1b to infer individuals belonging to haplogroup E. We used genotypes for SNP rs13447378 (phylogenetic marker M285) specific for subclade G1 and SNP rs34134567 (phylogenetic marker L30) specific for subclade G2a2 to infer individuals belonging to haplogroup G. We used genotypes from SNP rs2032673 (phylogenetic marker M69) specific for subclade H1a to infer individuals belonging to haplogroup H. We used SNP rs2032597 (phylogenetic marker M170) to infer haplogroup I. We used rs13447352 (phylogenetic marker M304) to infer haplogroup J. Genotypes of SNP rs3911 (phylogenetic marker M20) where used to infer haplogroup L. Genotypes of SNP rs34442126 (phylogenetic marker M46) specific for N1c1 were used to infer individuals belonging to haplogroup N. SNP rs16981290 (phylogenetic marker P186) was used to infer individuals belonging to haplogroup O. Genotypes of SNP rs8179021 (phylogenetic marker M242) where used to infer haplogroup Q. Haplogroup R was inferred based on genetic variation in SNP rs2032658 (phylogenetic marker M207). We used SNP rs20320 (phylogenetic marker M184) to infer haplogroup T.

*Level 1 haplogroups*

Information from inferred haplogroups were used as basis to further infer subclades. To infer subclades, we used haplogroups as inclusion criteria and sub-grouped within haplogroups. We used genotypes for SNP rs13447378 (phylogenetic marker M285) to infer subclade G1. Genotypes of SNP rs34134567 (phylogenetic marker L30) specific for subclade G2a2 were used to infer individuals belonging to subclade G2. We used rs9341296 (phylogenetic marker M253) to infer subclade I1a. Genotypes of SNP rs35547782 (phylogenetic marker L68) where used to infer subclade I2. We used SNP rs9341313 (phylogenetic marker M267) to infer subclade J1. Genotypes of SNP rs2032604 (phylogenetic marker M172) where used to infer subclade J2. Haplogroup R was sub-grouped into deeper subclade levels to follow the phylogenetic three of haplogroup R branches in the pandemic. Genotypes of SNP (phylogenetic marker M207) were used to infer subclades of R. We used genetic variation in SNP rs3908 (phylogenetic marker M17) specific for subclade R1a1 to infer individuals belonging to R1a. Genotypes of SNP rs9786184 (phylogenetic marker M343) were used to infer subclade R1b.

*Level 2 haplogroups*

Subclade R1b was further sub-grouped by genotypes carrying SNP rs9786076 (phylogenetic marker L11; R1b1a1b1a1a) into the two subclades of U106 and P312. These markers constitute mostly Germanic (phylogenetic marker U106; R1b1a1b1a1a1) and mostly Spanish, Italian (phylogenetic marker P312; R1b1a1b1a1a2). The individuals included in subclade R1b1a1b1a1a were assigned to P312 by exclusion from ancestral genetic variation in SNP rs16981293 (phylogenetic marker U106).

*Level 3 haplogroups*

Subclade P312 (also known as S116) was further subdivided into subclades L21 using genetic variation in SNP rs11799226 and U152 using genetic variation in SNP rs1236440. A larger group constituted individuals of P312 not including L21 and U152. This group contain one of the Italian and Spanish branches of R1b that initiated the interest in haplogroup R1b.

Based on the analyses, SNP rs9786076 (phylogenetic marker L11; R1b1a2a1a) representing the level above P312 and U106 made it possible to trace the association signal in those having marker L11 (including P312 and U106) and compare to those with subclades that branched of before L11 (lacking P312 and U106) (annotated: R1b_BranchBefore (L11)).

***Y-chromosome association analysis***

Association analyses were performed as described in analyses I-III using logistic regression in R as well as meta-analysis with the R-package metafor.(18) Additional analysis was performed as described in Section "*Association analysis and meta-analyses of candidate loci*".

VI. *Y-chromosome association analysis on mortality*

For each individual case dataset, COVID-19 patients were stratified based on their survival status (**Supplementary Table 1e**), which we used to define a new case-control status (control=alive; case=deceased). Statistical testing was performed as:

Case/Control ~ SNP + age + sex + age*age + age*sex + PC1 + PC2 + PC3 +PC4 + PC5 + PC6 + PC7 + PC8 + PC9 + PC10

Analysis on survival status was performed for the Y-chromosome to test the specific hypothesis that Y-chromosome haplogroups are associated with COVID-19 mortality.

#### Supplementary Information, Results and Discussion

##### *Functional characterization of genome-wide significant lead variants and associated candidate genes*

###### *Expression analysis*

On the tissue level, expression of candidate genes from 19q13.33 locus, including *NAPSA* and *KCNC3*, show high tissue specificity. For example, *NAPSA* is enhanced in lung, while *KCNC3* is highly expressed in brain and thyroid gland tissues (**Figure 1** and **Supplementary Figure 12**). Meanwhile, the *NR1H2* gene is broadly expressed among human tissues. Among immune cell types, all candidate genes of the 19q13.33 show low specificity and expression, only the *NR1H2* gene shows higher expression in eosinophils. These observations from bulk RNA-seq data are consistent with sc-RNA-Seq data of healthy lung cells where *NAPSA* is specifically expressed in type 1 and type 2 alveolar cells, while *NR1H2* is broadly expressed among lung cell types **Supplementary Figure 12b**). Notably, the *NR1H2* gene is significantly down-regulated in some parenchymal (basal, ciliated, club) and endothelial (pericyte) lung cells and is up-regulated in monocytes of COVID-19 patients compared to healthy controls. Interestingly, *NAPSA* shows significantly increased expression in type 1, but not in type 2 alveolar cells of COVID-19 patients (**Supplementary Figure 12c**).

For candidate genes of the 17q21.31 inversion locus, tissue gene expression data show their strong enrichment in the neural system (**Supplementary Figure 12a**). For example, protein-coding genes, including *ARL17B*, *CRHR1*, *KANSL1*, *LRRC37A2*, *MAPT*, *NSF*, *PLEKHM1* and *STH*, show high expression in brain tissues and the pituitary gland. Other genes, such as A*RHGAP27* and *FMNL1*, display broader expression among tissues and are enriched in whole blood. Expression data of bulk immune cell types show that the majority of the 17q21.31 locus candidate genes are highly enriched in myeloid cells such as mature eosinophils (including *ARHGAP27*, *FMNL1*, *KANSL1*, *LRRC37A*, LRRC37A2 and *PLEKHM1*) and neutrophils (including *FMNL1* and PLEKHM1). However, some of them (including *ARL17A*, *ARL17B*, *KANSL1*, *FMNL1* and *NSF*) are broadly distributed among myeloid and lymphoid immune cells. The expression levels are consistent with the ones in sc-RNA-Seq data of healthy upper airway cells, where the *KANSL1* gene is expressed in tissue-resident luminal macrophages and lymphatic cells, while *FMNL1* and *PLEKHM1* are expressed in neutrophils and *FMNL1* alone is expressed in dendritic, T and NK cells (**Supplementary Figure 12b**).

The expression levels in brain tissues in bulk (**Supplementary Figure 12a**) as well as in sc-RNA-seq data (**Supplementary Figure 12b**) showed consistent and high expression of *MAPT*, *NFS* and *KANSL1* genes, while other candidate genes from the 17q21.31 locus did not show noticeable expression in the brain sc-RNA-seq dataset. Unsurprisingly, candidate genes that are normally expressed in immune cells were also found to be deregulated in the lung tissue-resident immune cells of COVID-19 patients compared to heathy controls (**Supplementary Figure 12c**).

For example, genes including *PLEKHM1*, *NSF*, *FMNL1*, *LRRC37A2*, *ARL17A* and *ARHGAP27* are found to be up-regulated in myeloid as well as lymphoid cells such as monocytes, neutrophils, CD8+ T cells, NK cells, Treg cells, etc. Interestingly, some candidate genes from the 17q21.31 locus were also differentially expressed in parenchymal and endothelial cells. For example, besides being up-regulated in immune cells, *ARHGAP27* is significantly deregulated in ciliated, club and goblet cells. Another candidate gene, *KANSL1* is significantly up-regulated in vascular endothelial cells (**Supplementary Figure 12c**).

The 17q21.31 inversion is a lead cis-eQTL for 11 genes in unstimulated and/or stimulated CD14+ monocytes (**Supplementary Figure 13**). For a few genes, including *AC126544.1*, *AC126544.2*, *FAM215B*, *KANSL1*, *KANSL1-AS1* and *LINC02210*, the inverted allele H2 has the strongest eQTL associations under monocyte stimuli with bacterial or viral agents. In particular the protein-coding gene *KANSL1* shows increased expression after treatment with human influenza A virus (IAV) in the presence of the H2 allele. Interestingly, this H2 allele-linked increase of *KANSL1* expression levels turn out to be related to a significant increase of its protein-coding and non-coding isoforms (**Supplementary Figure 14**). For other genes, including *AC005829.2*, *DND1P1*, *LRRC37A*, *LRRC37A2*, and *LRRC37A2*, the inversion is a lead eQTL in unstimulated, as well as in stimulated, monocytes (**Supplementary Figure 13**). In general, most inversion-linked genes tend to be down-regulated after monocyte activation, while the inverted H2 allele is associated with increased expression of those genes (except for *FAM215B*, *LRRC37A* and *LRRC37A4P*), thus suggesting compensative role of linked-gene down-regulation under monocyte activation.

In brain organoids, only the *NR1H2* and *KCNC3* genes are found to be expressed among protein-coding candidate genes of the 19q13.33 locus (**Supplementary Figure 15**). *NR1H2* is expressed in all major cell types of brain organoids, while *KCNC3* is expressed in cortical neurons. As in the sc-RNA-seq dataset of non-diseased human brain tissues(25), candidate genes of the 17q21.31 locus, including *MAPT*, *KANSL1* and *NFS*, showed high expression in neural cells (neural progenitors, interneurons, neurons, and cortical neurons) of human brain organoids (**Supplementary Figure 15a**). Other genes such as *ARL17B*, *FMNL1* and *PLEKHM1* were expressed at much lower rates, while the *CRHR1* candidate gene was not noticeably expressed in brain organoids. From the inversion locus, only the *MAPT* gene is differentially expressed in the SARS-CoV-2 infected cells. Its down-regulation is highly consistent after 96 hours of infection among premature and mature neuronal cells, namely interneurons, neurons, and cortical neurons (**Supplementary Figure 15b**).

##### *HLA locus fine mapping and association analysis*

###### *HLA typing and imputation*

The imputation of the 6,322 Italian, 6,580 Spanish, 364 Norwegian and 3,639 German/Austrian samples resulted in a total of 279 different 2-field alleles with the two main-panels (new Spanish-Italian panel and multi-ethnic HLARES, respectively). The number of alleles per locus and cohort are shown and the marginal probability for the imputation at 2-field resolution is presented in **Supplementary Table 16a/b**. No allele with an allele frequency above 0.05% showed a marginal probability below 0.3.

###### *HLA fine mapping of association*

There were no association signals meeting either the genome-wide significance threshold of $P=5x10^{-8}$ or the suggestive association significance threshold of $P=1x10^{-5}$, neither for disease risk nor for disease severity (**Supplementary Figure 16, Supplementary Table 16c).** We further investigated specific HLA alleles that had been suggested to potentially play a role in SARS-CoV-2 susceptibility or COVID-19 severity.(59–72) For each of the potential alleles, we explored if our data showed at least a nominally significant trend in the direction as proposed in the different studies, but for none of the potential alleles, nominal significance and same effect direction was observed with the association analysis closest to the one performed in the different studies across all our cohorts. Overall, we would like to point out, that all reported associations were based on less than 250 patients in the affected group and there is no consistency between the reported signals. Further, some findings do not stay significant after correcting for multiple testing and others do not report any P-values corrected for multiple testing. To our knowledge the largest so far published study on this topic was performed on 6,413 Israelian COVID-19 patients and, in agreement with our study, does not show any significant association.(62)

***Peptidome-wide association study (PepWAS)***

The total number of unique 9mer and 15mer peptides from the reference proteome of SARS-CoV-2 was 9,910 and 9,814, respectively. After correcting for multiple testing using Bonferroni correction, no statistically significant association between HLA-presented SARS-CoV-2 peptides and COVID-19 could be established neither for disease risk (all cases vs. the general population) nor for disease severity (mild cases vs. severe cases). Exemplary results for the largest, Spanish cohort for the two different association tests are shown in **Supplementary Figures 17-18.** The complete results and peptide lists are shown in **Supplementary Table 17.**

***Analysis of quantitative HLA parameters***

We found no robust statistically significant associations between any of the tested HLA compound parameters that exceeded the statistical significance thresholds for disease risk or disease severity, in line with previous results on a smaller dataset.(44) Neither the number of classical HLA alleles of an individual nor the HLA allele divergence at specific loci or across multiple loci differed significantly between cases and controls or between groups of cases with different respiratory support. Similarly, when computationally predicting individual HLA-binding of SARS-CoV-2 peptides, we found no robust statistically significant association between the different peptide values and either disease risk (case vs. control) or disease severity (different levels of respiratory support). A few of the tested parameters showed nominally significant associations but did not replicate across cohorts (**Supplementary Table 16**) and are thus likely statistical artifacts or caused by unaccounted population stratification with no consequence for COVID-19 risk. We also tested for quadratic associations, investigating the possibility of an optimal HLA diversity, but found no support for this either. The correlation analysis between risk/severity association and number of bound SARS-CoV-2 peptides across HLA alleles did also not yield any robust significant associations (**Supplementary Table 17**).

***HLA-presentation of shared peptides ('molecular mimicry')***

Out of all unique 9mer and 15mer peptides presented in the SARS-CoV-2 reference proteome, we found exact matches to human self-peptides for two and zero peptides, respectively. The two 9mer peptides with a perfect match, KKDKKKKAD and SSRSSSRSR, both originate from the Nucleocapsid (N) protein. Intriguingly, the peptide SSRSSSRSR was significantly more likely to be presented by HLA of severe cases compared to the general population, yielding an overall nominally significant association with higher risk for severe COVID-19 (P=0.034). However, albeit the effect direction for this peptide was the same across all three cohorts, its association with disease risk was only significant in the German/Austrian cohort. It therefore does not seem to represent a general risk factor for COVID-19.

When screening for peptides that are predicted to be bound by all common alleles of a given locus, we identified a set of 75 viral 9mer peptides that were presented by all common HLA-*C* alleles of the present dataset (no such peptides were found for HLA-*A* or HLA-*B*). Of these peptides, 24 showed enhanced sequence similarity to human self-peptides following the criteria outlined in the method section. However, the human lung-expressed proteins to which these peptides mapped, were not significantly enriched for any GO-terms

*Supplementary details on the Y-chromosomal haplogroup analysis*

***Introduction***

The COVID-19 pandemic spread unequally across regions and countries. Specific regions, like Northern Italy, have been particularly affected, e.g. Bergamo had an increased COVID-19 mortality. This region has a high frequency of Y-chromosome haplogroup R1b (80.8%)(73) the haplogroup that also dominates in Western Europe and North and South America.(74–79) During late 2020, the pandemic shifted and affected countries eastwards in Europe more severely while the global R1b correlation lost strength. South-east Asia, with a high dominance of other Y-haplogroups, still have very low COVID-19 mortality.(80) We found a significant correlation between COVID-19 mortality and haplogroup R1b in two regression correlation studies conducted in regions of Italy in addition to 34 European countries (plus China and India) in May and September 2020.(81) These findings, corroborate one other study(82), and in addition to knowledge regarding increased mortality in males versus females indicates a potential impact of Y chromosomal genetic mechanisms on COVID-19 severity. There are several genes on the Y chromosome involved in mechanisms of androgen receptor regulation, as well as in immunity and inflammation like *KDM5D*, *UTY*, *DDX3Y*, *MSL3* and *USB9Y* that could influence both virus transmission and immune response to virus.(83) To this end the Y-linked KDM5D is an important regulator of androgen receptor levels and involved in regulation of TMPRSS2 known to enable SARS-CoV-2 binding to ACE2.(84–87) Genetic variation in *KDM5D* across Y chromosome haplogroups suggest a possibility for ‘fine tuning' of androgen receptor levels varying across haplogroups and for R1b specific variations of *KDM5D* to partly explain COVID-19 severity and mortality.

This is consistent with previous research finding androgen mechanisms involved in the pathological pathways of COVID-19 e.g., androgen sensitive prostate cancer.(88–90)
Based on the observed correlations our hypothesis became that the mortality of COVID-19 was partly associated with haplogroups on the Y chromosome, particularly haplogroup Y-R1b as a marker of mortality during the first wave.

***Methods***
Stratified analyses were carried out according to analyses III-VI on male individuals

***Results***
Results are shown in **Supplementary Table 19**. We observe suggestive association (P<0.05) with increased risk for severe respiratory COVID-19 with members of the R haplogroup (M207) in males primarily in the Italian population. This is specifically observed within the R1b haplogroups (as opposed to R1a1). Not carrying the haplogroup R was observed to be protective. The strongest effects in the first and second analysis are observed in the age group > 60 years in the Italian population, with effect estimates, albeit not significant, being lower in younger age groups. The main R (M207) haplogroup (level 0) showed an increasing risk for severe respiratory COVID-19 (first analysis: age > 80 years: P=0.0017, OR=4.13, 95%CI=1.70-10.00). The strongest association was observed for haplogroup R1b1a2a2 (P312) in the Italian population and age group > 80 years ($P=1.151 \times 10^{-4}$, OR=14.88, 95%CI=3.78-58.65). Signals observed in the R1b group U152 are most likely reflecting batch effects by different genotyping platforms, since a large coefficient of variation is observed here. Careful follow-up analyses are therefore warranted. Apart from the association of mortality with Level2:R1b1a2a1:(U106) in the Spanish and Italian population (**Main Manuscript)**, none of the signals, however replicate across our cohorts.

***Discussion***

Analysis of the Y-chromosome in COVID-19 revealed no consistent signals of across our different cohorts. However, we observed an association between haplogroup R1b and severe respiratory COVID-19 as well as severe respiratory COVID-19 mortality in the Italian cohort. The results indicate an increased risk of severe respiratory COVID-19 for R1b. These results are from the first wave of the pandemic (until September 2020), when the pandemic affected Western and not Eastern Europe to the same degree. Further research on different viral mutations and their potential unequal susceptibility for different haplogroups in later waves of the pandemic, would be of interest regarding this.

Limitations in the study are the low sample sizes in the stratified groups and the lack of consistency between the populations in the study when differentiating the data, this requires a careful interpretation of the results. In addition to different frequencies of haplogroups in the Italian and Spanish data, this might explain the lack of consistency between the populations

#### Supplementary Figures

**Supplementary Figure 1. Principal components analysis of COVID-19 cases and controls.**

Scatter plots of the principal component analysis (PCA) for cases and controls using the PCA method as implemented in FlashPCA(6), using an LD-pruned subset of SNPs (**Supplementary Methods**). Ancestry outliers not matching European populations were removed (a, b, c and d). After QC, PCA revealed no non-European ancestry outliers (e, f, g and h) when performing PCA including reference samples from the 1,000 Genomes reference panel.(2)

(a) Italian cases and controls before exclusion of ancestry outliers.
(b) Spanish cases and controls before exclusion of ancestry outliers.
(c) German/Austrian cases and controls before exclusion of ancestry outliers
(d) Norwegian cases and controls before exclusion of ancestry outliers.
(e) Italian cases and controls after exclusion of ancestry outliers.
(f) Spanish cases and controls after exclusion of ancestry outliers.
(g) German/Austrian cases and controls after exclusion of ancestry outliers.
(h) Norwegian cases and controls after exclusion of ancestry outliers.

The grey crosses represent COVID-19 cases and controls. The colored points represent the five super populations retrieved from the 1,000 Genomes Phase 3 data.(2)
African (AFR, purple), Ad Mixed American (AMR, turquoise), South Asian (SAS, green), East Asian (EAS, blue), (EUR, yellow). Population code (super population code): CHB (EAS) Han Chinese in Beijing, China; JPT (EAS) Japanese in Tokyo, Japan; CHS (EAS) Southern Han Chinese; CDX (EAS) Chinese Dai in Xishuangbanna, China; KHV (EAS) Kinh in Ho Chi Minh City, Vietnam; CEU (EUR) Utah Residents (CEPH) with Northern and Western European Ancestry; TSI (EUR) Toscani in Italia; FIN (EUR) Finnish in Finland; GBR (EUR) British in England and Scotland; IBS (EUR) Iberian Population in Spain; YRI (AFR) Yoruba in Ibadan, Nigeria; LWK (AFR) Luhya in Webuye, Kenya; GWD (AFR) Gambian in Western Divisions in the Gambia; MSL (AFR) Mende in Sierra Leone; ESN (AFR) Esan in Nigeria; ASW (AFR) Americans of African Ancestry in SW USA; ACB (AFR) African Caribbeans in Barbados; MXL (AMR) Mexican Ancestry from Los Angeles USA; PUR (AMR) Puerto Ricans from Puerto Rico; CLM (AMR) Colombians from Medellin, Colombia; PEL (AMR) Peruvians from Lima,

Peru; GIH (SAS) Gujarati Indian from Houston, Texas; PJL (SAS) Punjabi from Lahore, Pakistan; BEB (SAS) Bengali from Bangladesh; STU (SAS) Sri Lankan Tamil from the UK; ITU (SAS) Indian Telugu from the UK.

(a) Italian cases and controls before exclusion of ancestry outliers.

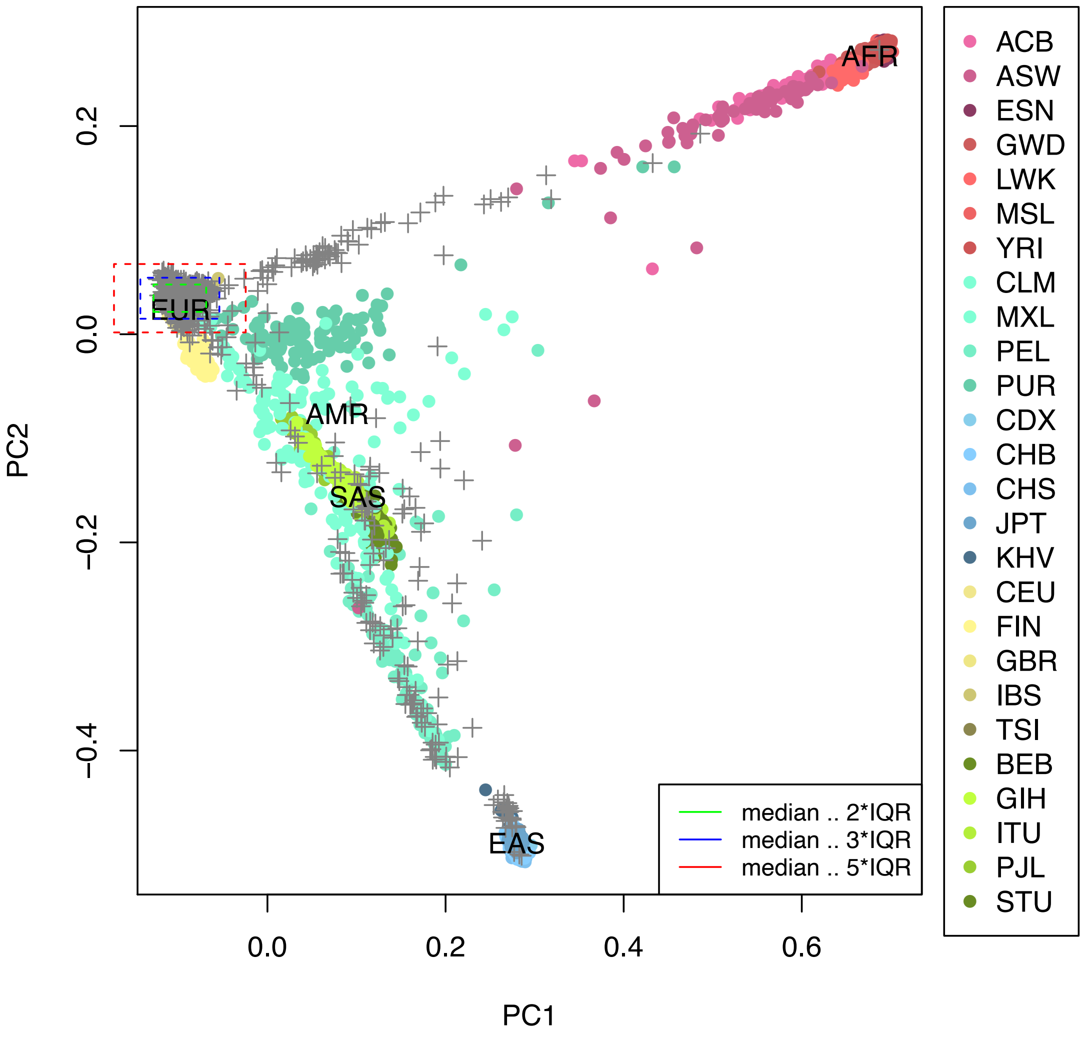

PC1: principal component 1, PC2: principal component 2

(b) Spanish cases and controls before exclusion of ancestry outliers.

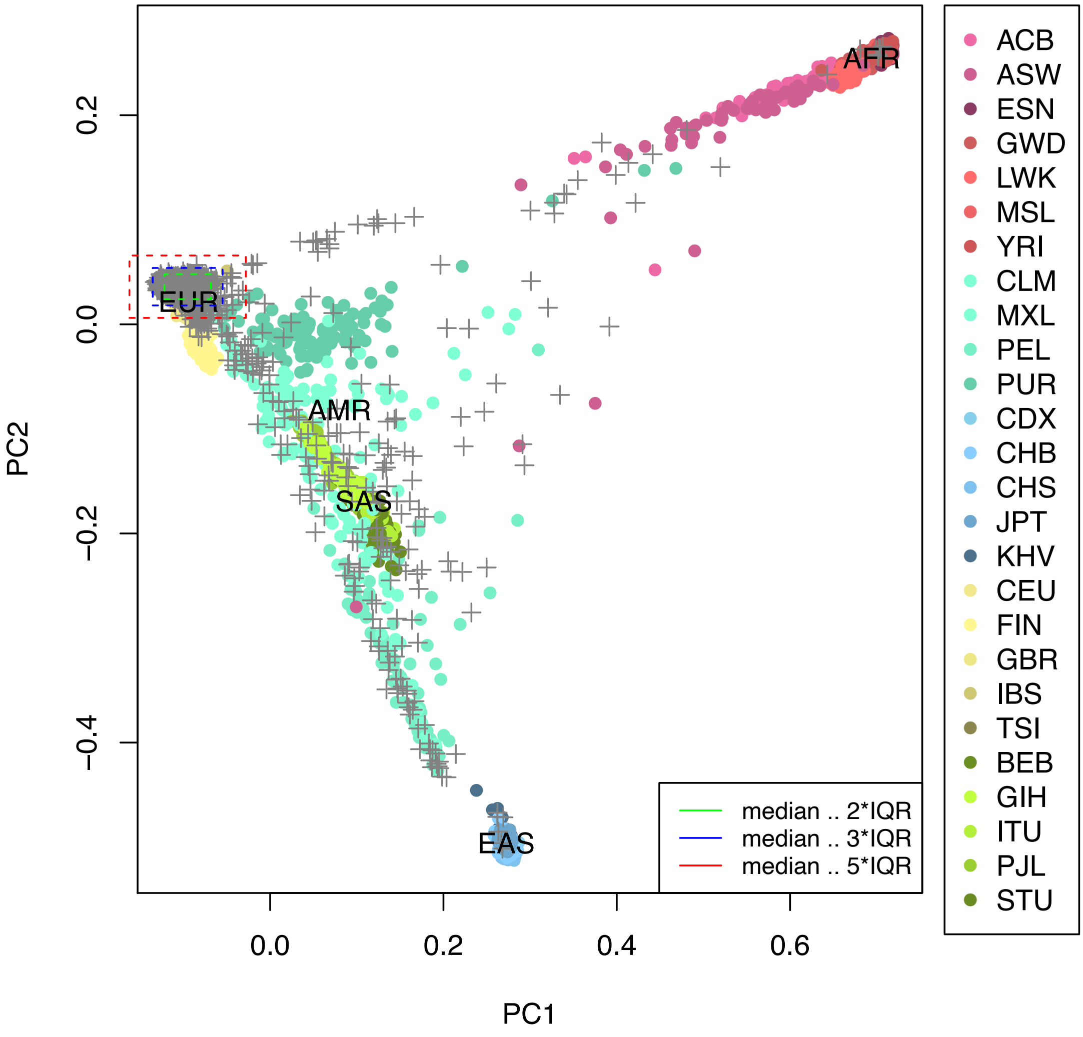

PC1: principal component 1, PC2: principal component 2

(c) German/Austrian cases and controls before exclusion of ancestry outliers.

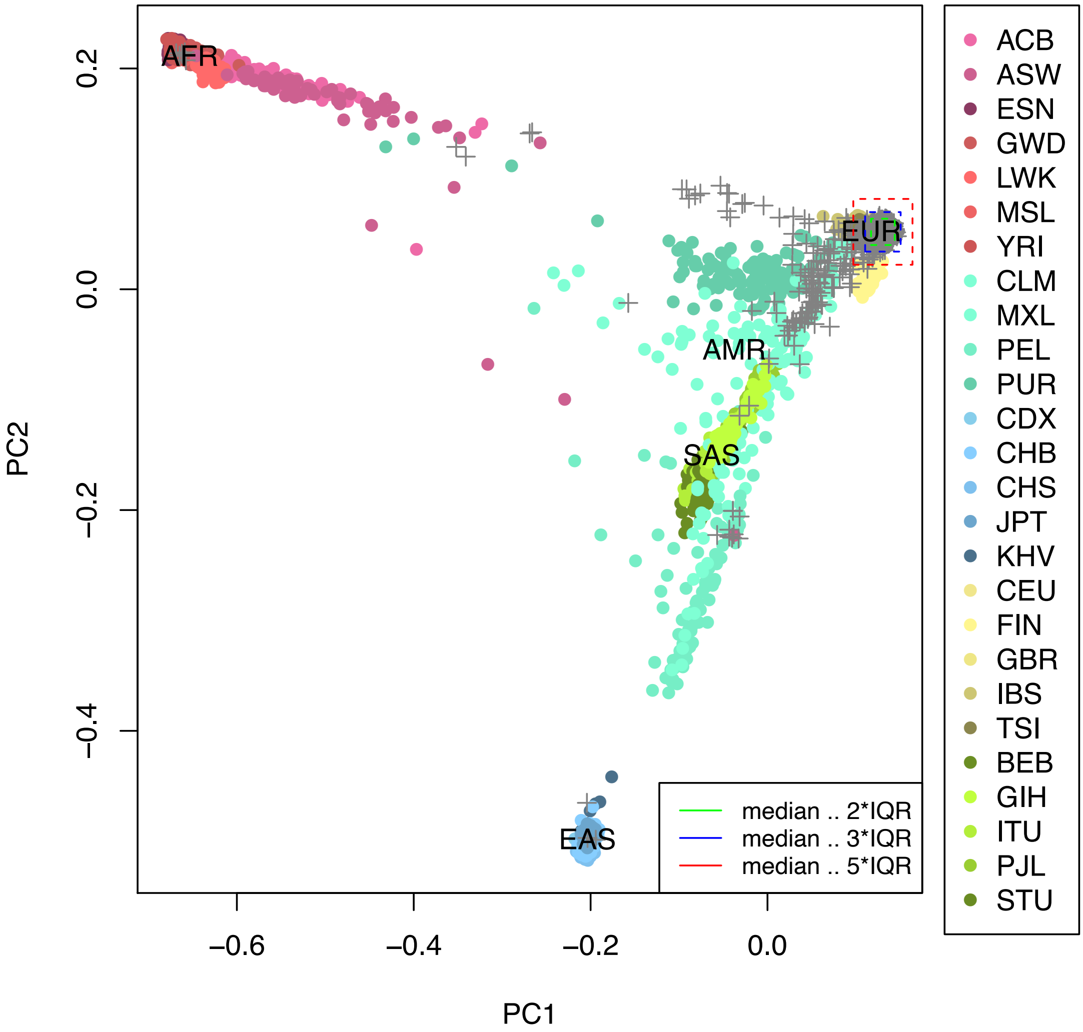

PC1: principal component 1, PC2: principal component 2

(d) Norwegian cases and controls before exclusion of ancestry outliers.

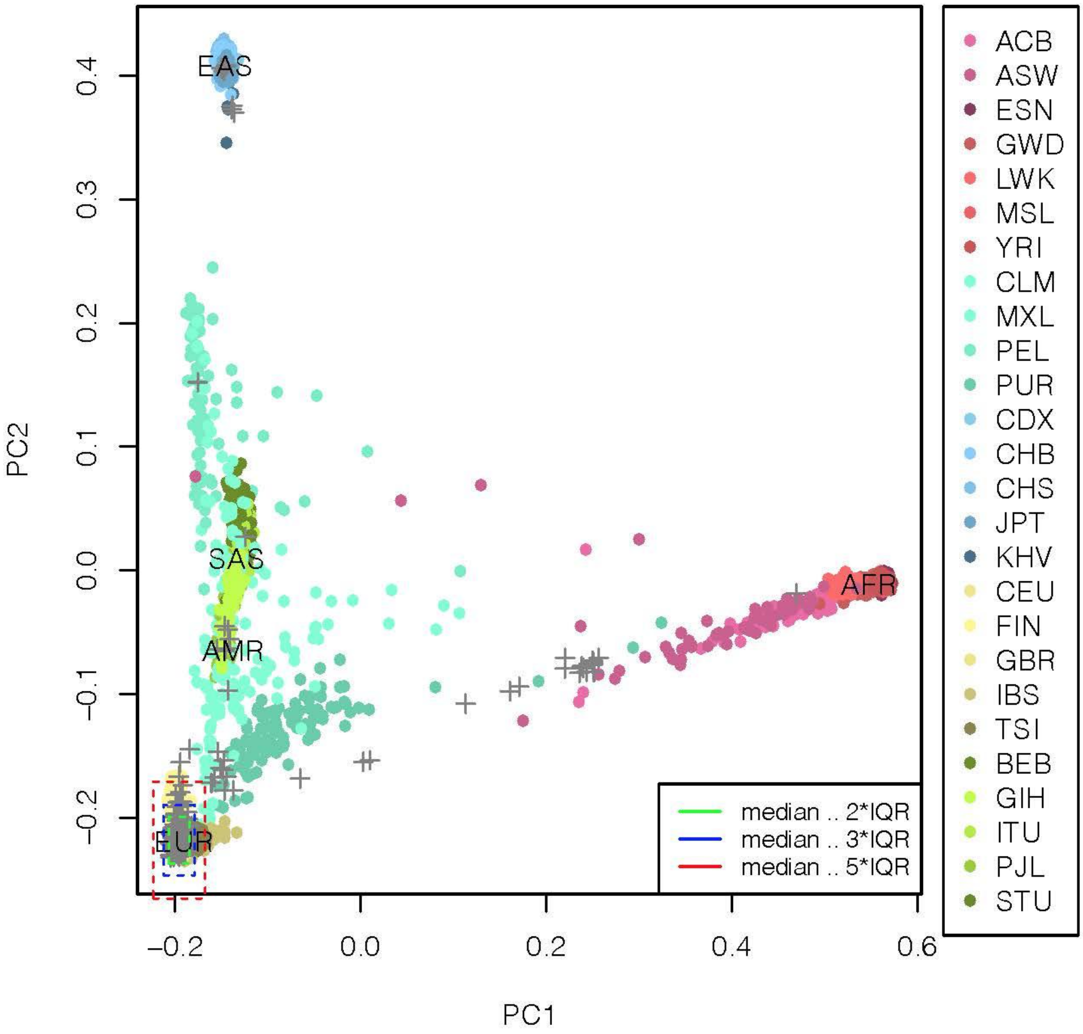

PC1: principal component 1, PC2: principal component 2

(e) Italian cases and controls after exclusion of ancestry outliers.

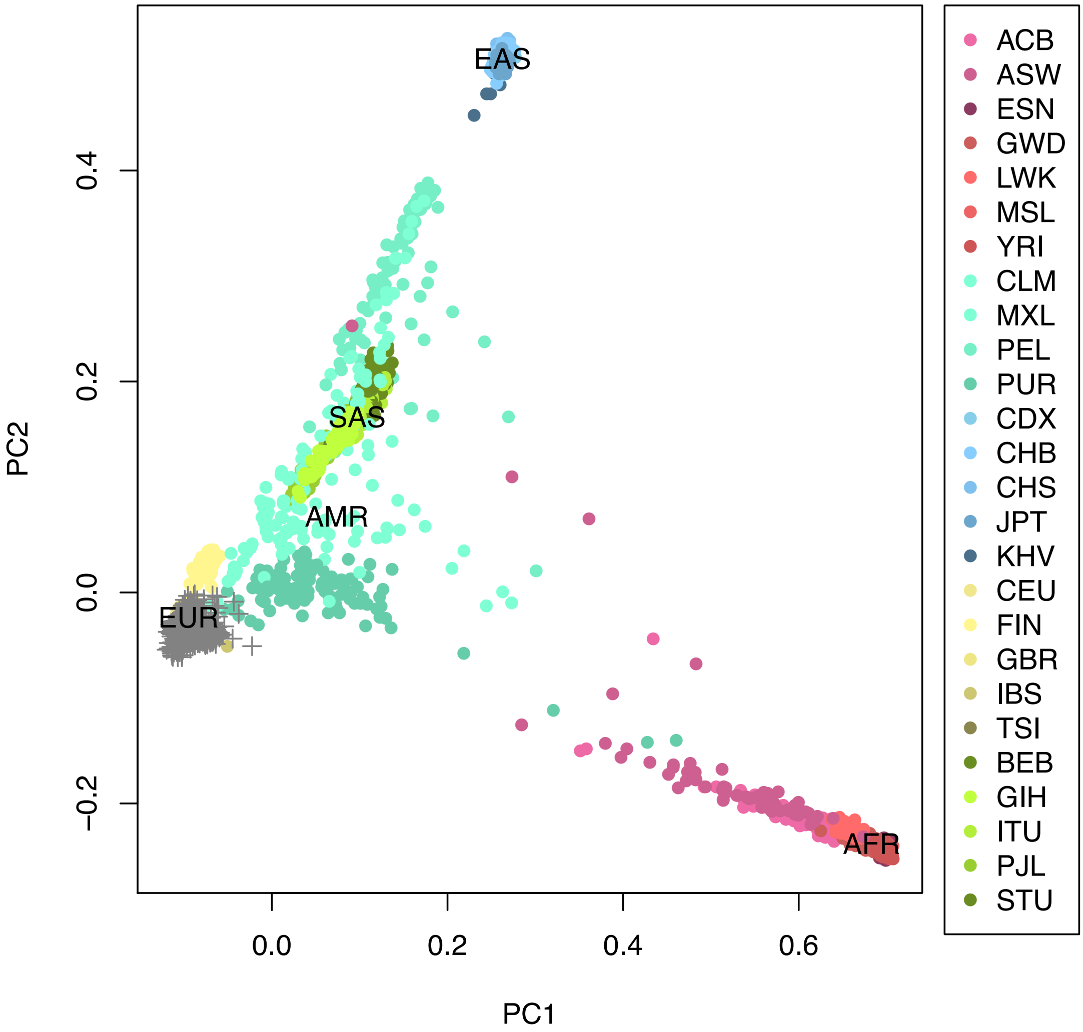

PC1: principal component 1, PC2: principal component 2

(f) Spanish cases and controls after exclusion of ancestry outliers.

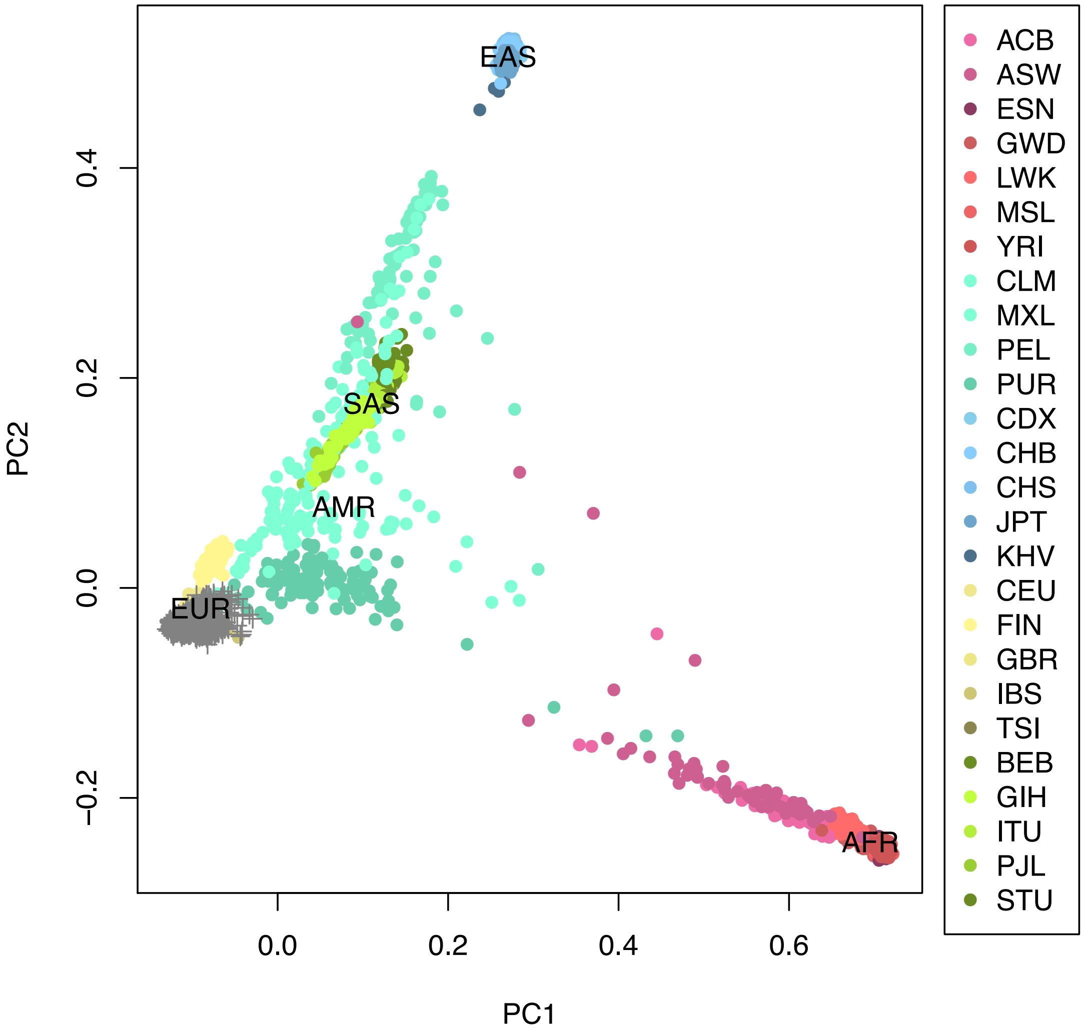

PC1: principal component 1, PC2: principal component 2

(g) German/Austrian cases and controls after exclusion of ancestry outliers.

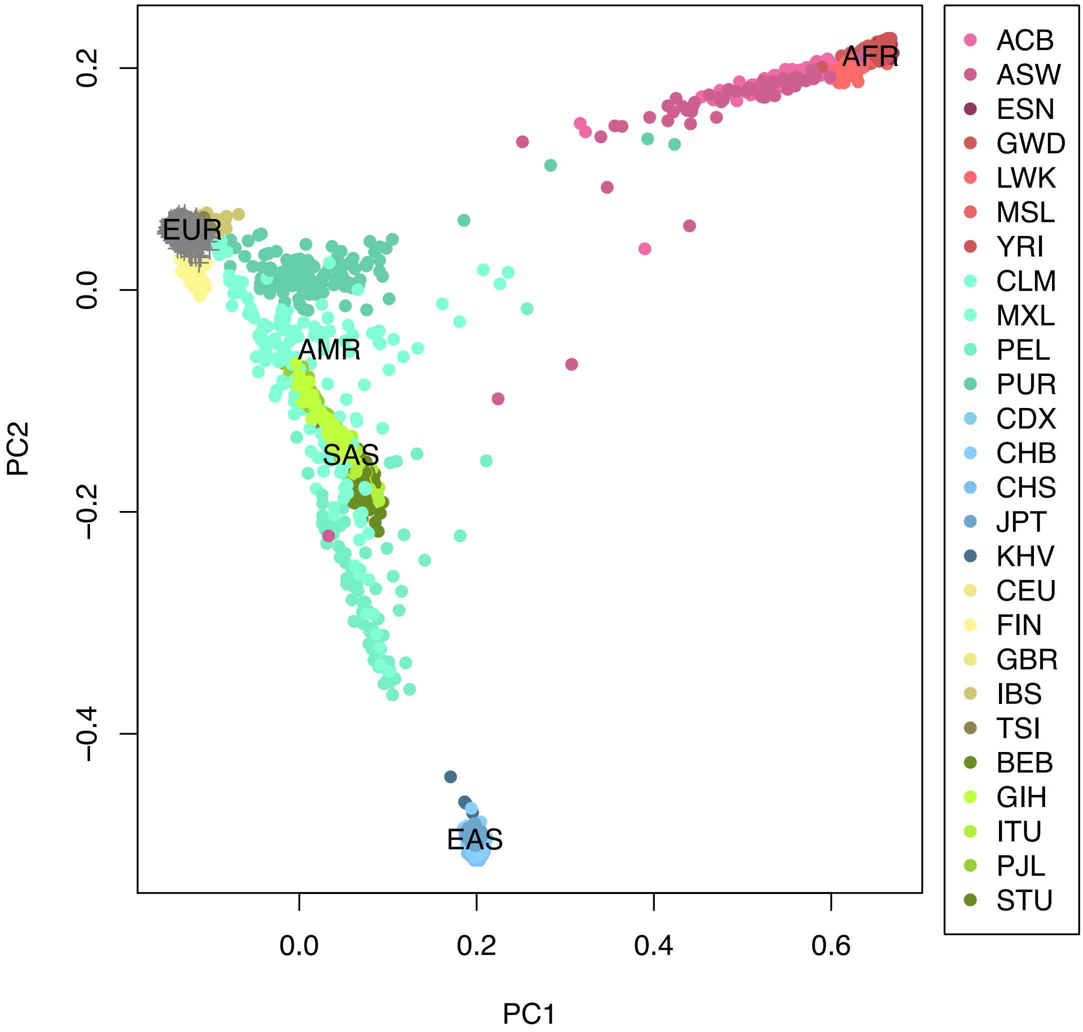

PC1: principal component 1, PC2: principal component 2

(h) Norwegian cases and controls after exclusion of ancestry outliers.

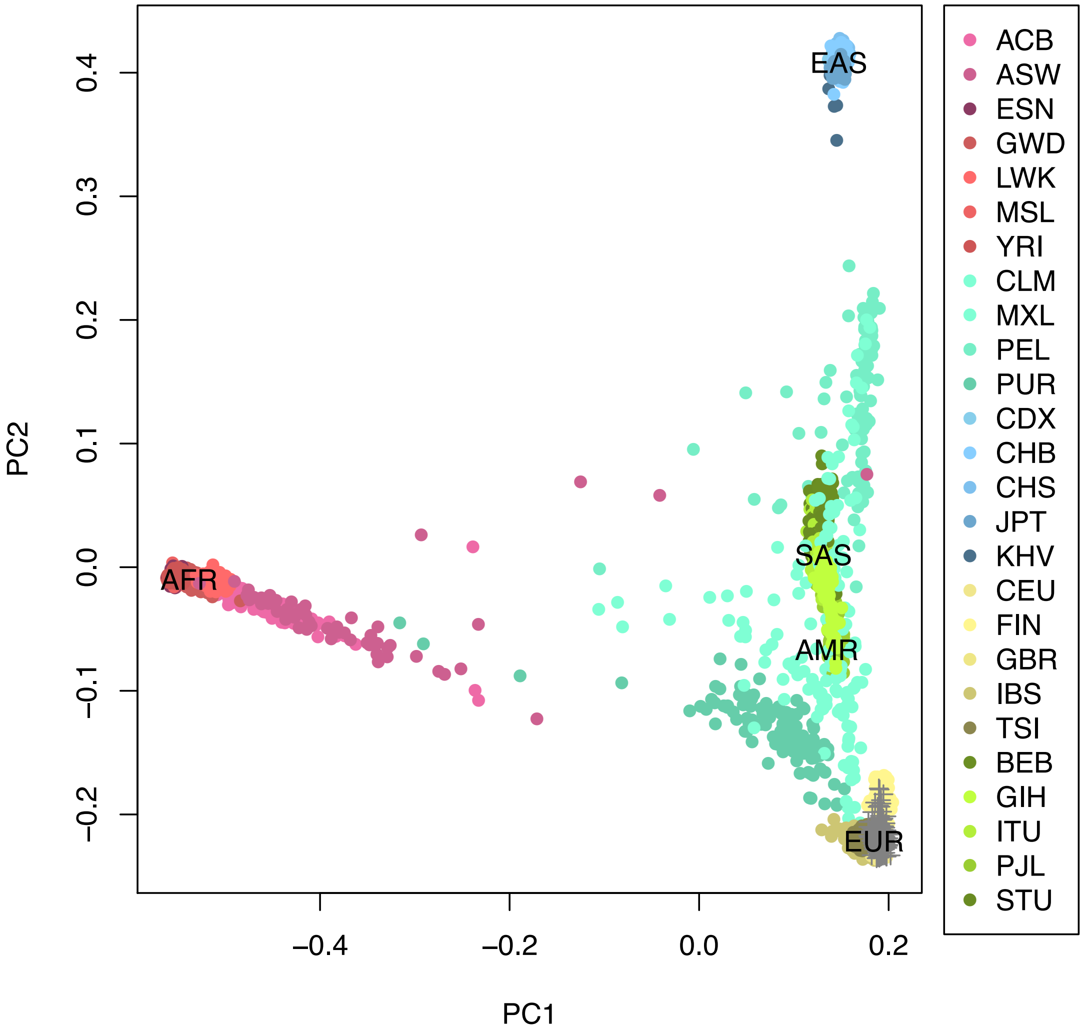

PC1: principal component 1, PC2: principal component 2

**Supplementary Figure 2. Manhattan and quantile-quantile plots from meta-analysis of Italian, Spanish, Norwegian and German/Austrian GWAS summary association statistics IIa**.

Shown are **(a)** Manhattan and **(b)** quantile-quantile (QQ) plots of the association statistics from our main analysis IIa (cases with respiratory support codes 1-4 vs. population controls; Supplementary Methods). The red dashed line indicates the genome-wide significance threshold of a P $<5\times10^{-8}$, the blue dashed line indicates a suggestive threshold of P $<10^{-6}$. Only markers that passed the imputation score $R^2\geq0.6$ and had a MAF≥1% were used for plotting. In the QQ plot, the 2.5th and 97.5th centiles of the distribution under random sampling and the null hypothesis form the 95% concentration band. The genomic inflation factor lambda (λ) is defined as the ratio of the medians of the sample χ2 test statistics and the 1-d.f. χ2 distribution (0.455).(91) Red dots indicate variants with meta-analysis heterogeneity P-value of < $10^{-5}$.

**(a)**

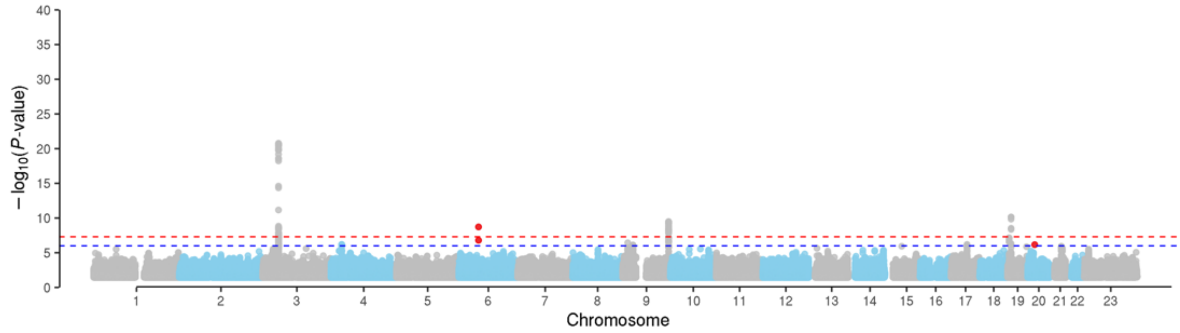

**(b)**

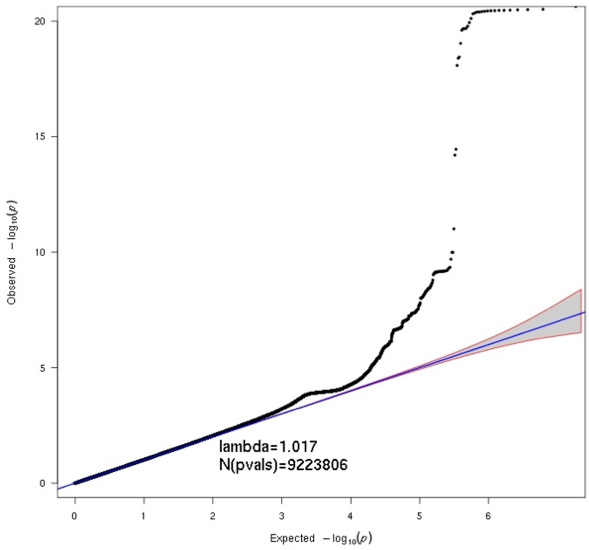

**Supplementary Figure 3. Manhattan plots quantile-quantile from meta-analysis of Italian, Spanish and German/Austrian GWAS summary association statistics (IIb).** Shown are **(a)** Manhattan and **(b)** quantile-quantile (QQ) plots of the association statistics from our main analysis IIb (cases with respiratory support codes 2-4 vs. population controls) Supplementary Methods). The red dashed line indicates the genome-wide significance threshold of a $P < 5 \times 10^{-8}$, the blue dashed line indicates a suggestive threshold of $P < 10^{-6}$. Only markers that passed the imputation score $R^2 \geq 0.6$ and had a MAF≥1% were used for plotting. In the QQ plot, the 2.5th and 97.5th centiles of the distribution under random sampling and the null hypothesis form the 95% concentration band. The genomic inflation factor lambda (λ) is defined as the ratio of the medians of the sample $\chi 2$ test statistics and the 1-d.f. $\chi 2$ distribution (0.455).(91) Red dots indicate variants with meta-analysis heterogeneity P-value of < $10^{-5}$.

**(a)**

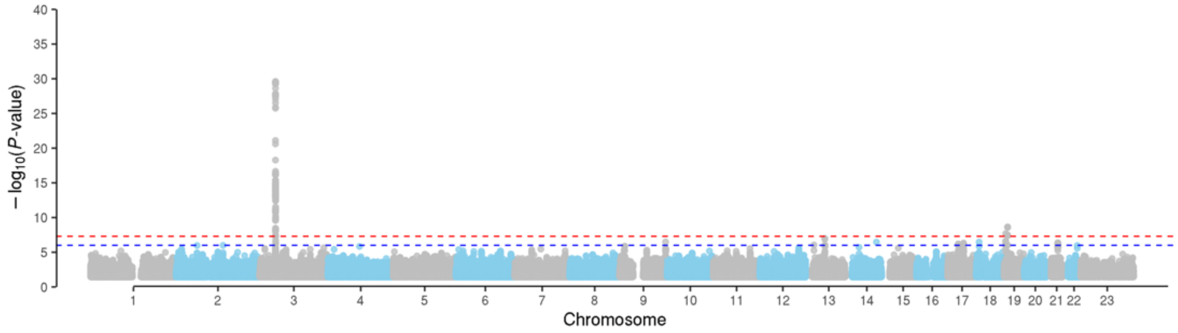

**(b)**

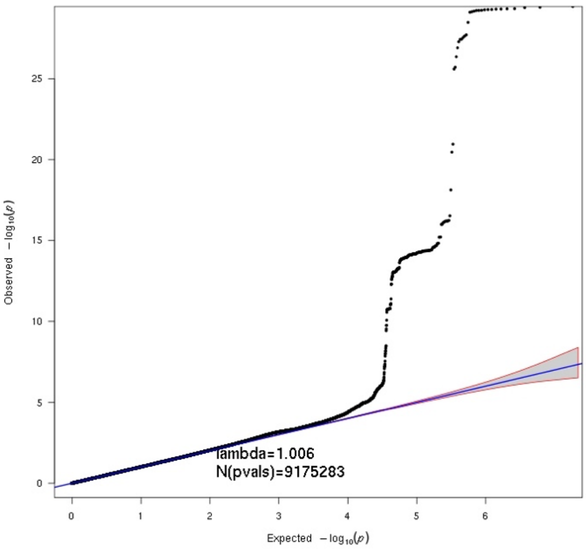

**Supplementary Figure 4. Regional association plots for suggestive loci from the first analysis.**

Plot was created using the LocusZoom tool.(92) LD values were calculated based on genotypes of the merged Italian/Spanish/Norwegian/German/Austrian dataset derived from TOPMed imputation (**Online Methods**). Genome build hg38 positions are plotted. The recombination rate is shown in centimorgans (cM) per million base pairs (Mb). The plot shows the names and locations of the genes; the transcribed strand is indicated with an arrow. Genes are represented with intronic and exonic regions. The purple diamond in each panel represents the variant most strongly associated with severe COVID-19 and respiratory failure. Set1 shows the 95% credible set from Bayesian fine mapping (**Online Methods**).

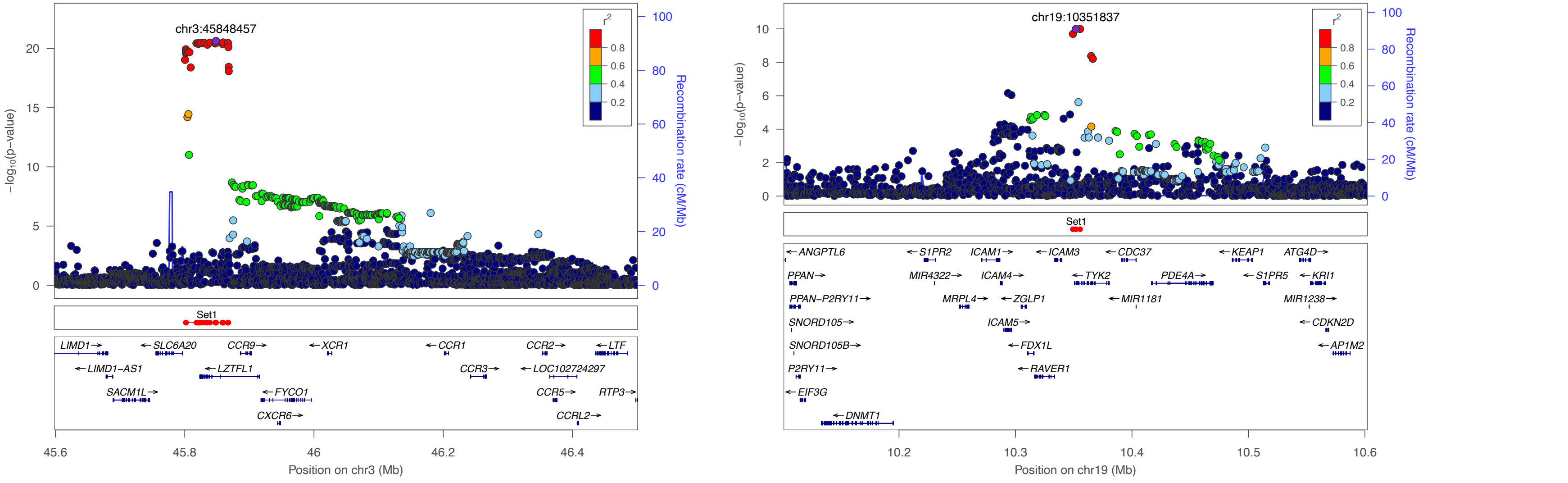

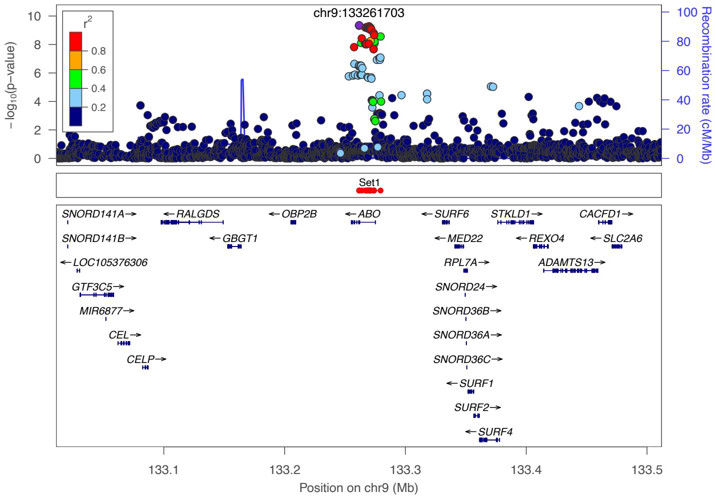
chr9:133261703
$r^2$
0.8
0.6
0.4
0.2
−log10(p−value)
Recombination rate (cM/Mb)
Set1
SNORD141A→
←RALGDS
←OBP2B
←ABO
←SURF6
STKLD1→
CACFD1→
SNORD141B→
←GBGT1
←MED22
←REXO4
←SLC2A6
←LOC105376306
RPL7A→
ADAMTS13→
GTF3C5→
SNORD24→
MIR6877→
SNORD36B→
CEL→
SNORD36A→
CELP→
SNORD36C→
←SURF1
SURF2→
←SURF4
133.1
133.2
133.3
133.4
133.5
Position on chr9 (Mb)

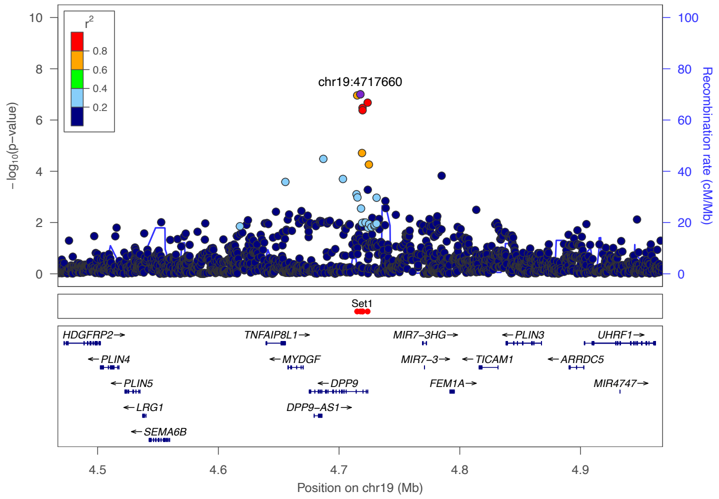
chr19:4717660
$r^2$
0.8
0.6
0.4
0.2
−log10(p−value)
Recombination rate (cM/Mb)
Set1
HDGFRP2→
TNFAIP8L1→
MIR7−3HG→
←PLIN3
UHRF1→
←PLIN4
←MYDGF
MIR7−3→
←TICAM1
←ARRDC5
←PLIN5
←DPP9
FEM1A→
MIR4747→
←LRG1
DPP9−AS1→
←SEMA6B
4.5
4.6
4.7
4.8
4.9
Position on chr19 (Mb)

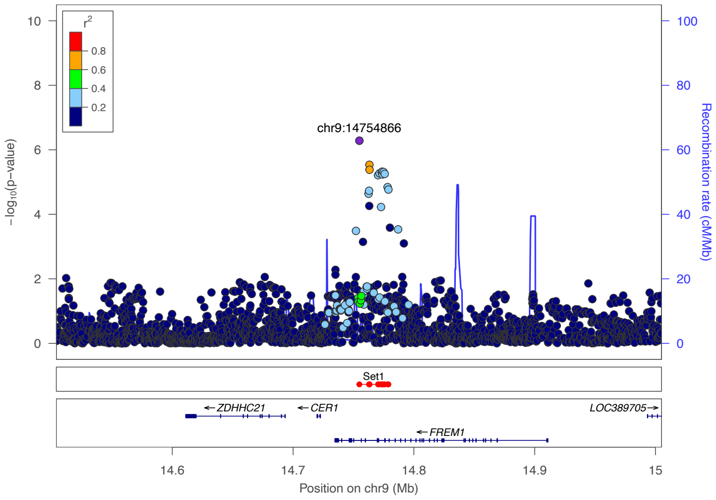
chr9:14754866
$r^2$
0.8
0.6
0.4
0.2
−log10(p−value)
Recombination rate (cM/Mb)
Set1
←ZDHHC21
←CER1
LOC389705→
←FREM1
14.6
14.7
14.8
14.9
15
Position on chr9 (Mb)

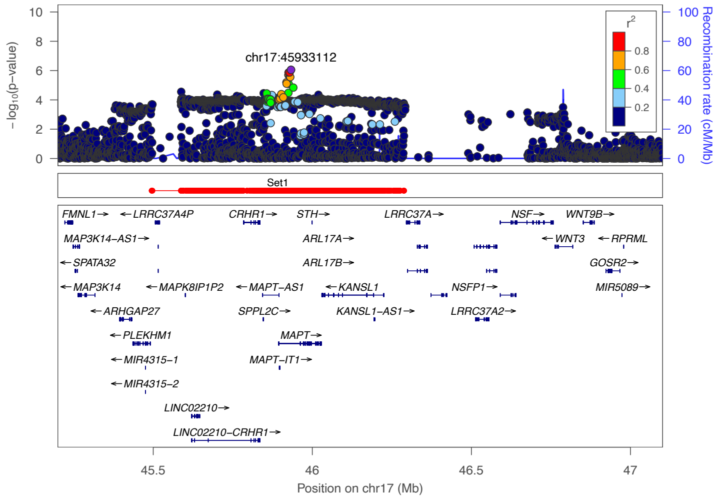
chr17:45933112
$r^2$
0.8
0.6
0.4
0.2
−log10(p−value)
Recombination rate (cM/Mb)
Set1
FMNL1→
←LRRC37A4P
CRHR1→
STH→
LRRC37A→
NSF→
WNT9B→
MAP3K14−AS1→
ARL17A→
←WNT3
←RPRML
←SPATA32
ARL17B→
GOSR2→
←MAP3K14
←MAPK8IP1P2
←MAPT−AS1
←KANSL1
NSFP1→
MIR5089→
←ARHGAP27
SPPL2C→
KANSL1−AS1→
LRRC37A2→
←PLEKHM1
MAPT→
←MIR4315−1
MAPT−IT1→
←MIR4315−2
LINC02210→
LINC02210−CRHR1→
45.5
46
46.5
47
Position on chr17 (Mb)

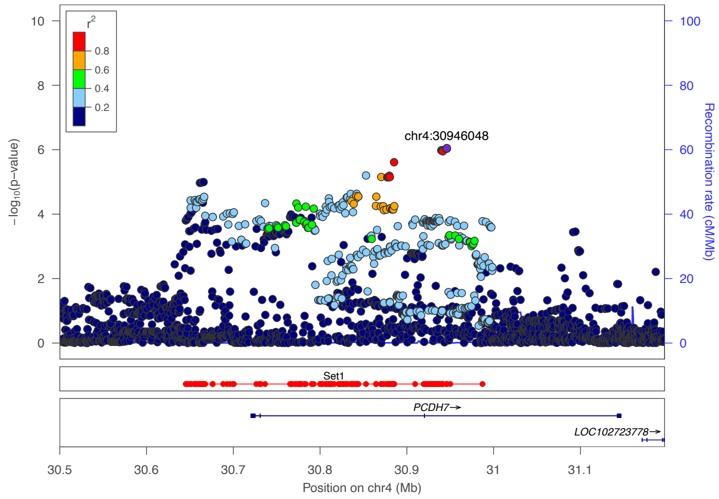

r2
0.8
0.6
0.4
0.2
chr4:30946048
−log10(p−value)
Recombination rate (cM/Mb)
Set1
PCDH7→
LOC102723778→
Position on chr4 (Mb)

**Supplementary Figure 5. Regional association plots for suggestive loci from the second analysis**.

Plot was created using the LocusZoom tool.(92) LD values were calculated based on genotypes of the merged Italian/Spanish/Norwegian/German/Austrian dataset derived from TOPMed imputation (**Online Methods**). Genome build hg38 positions are plotted. The recombination rate is shown in centimorgans (cM) per million base pairs (Mb). The plot shows the names and locations of the genes; the transcribed strand is indicated with an arrow. Genes are represented with intronic and exonic regions. The purple diamond in each panel represents the variant most strongly associated with severe COVID-19 and respiratory failure. Set1 shows the 95% credible set from Bayesian fine mapping (**Online Methods**).

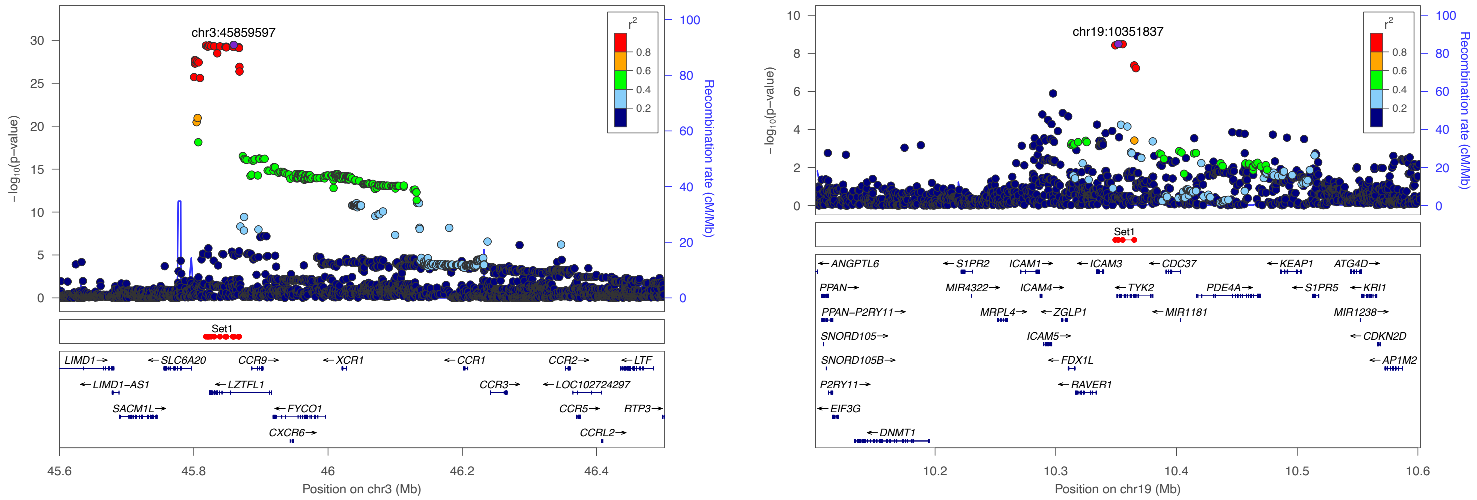

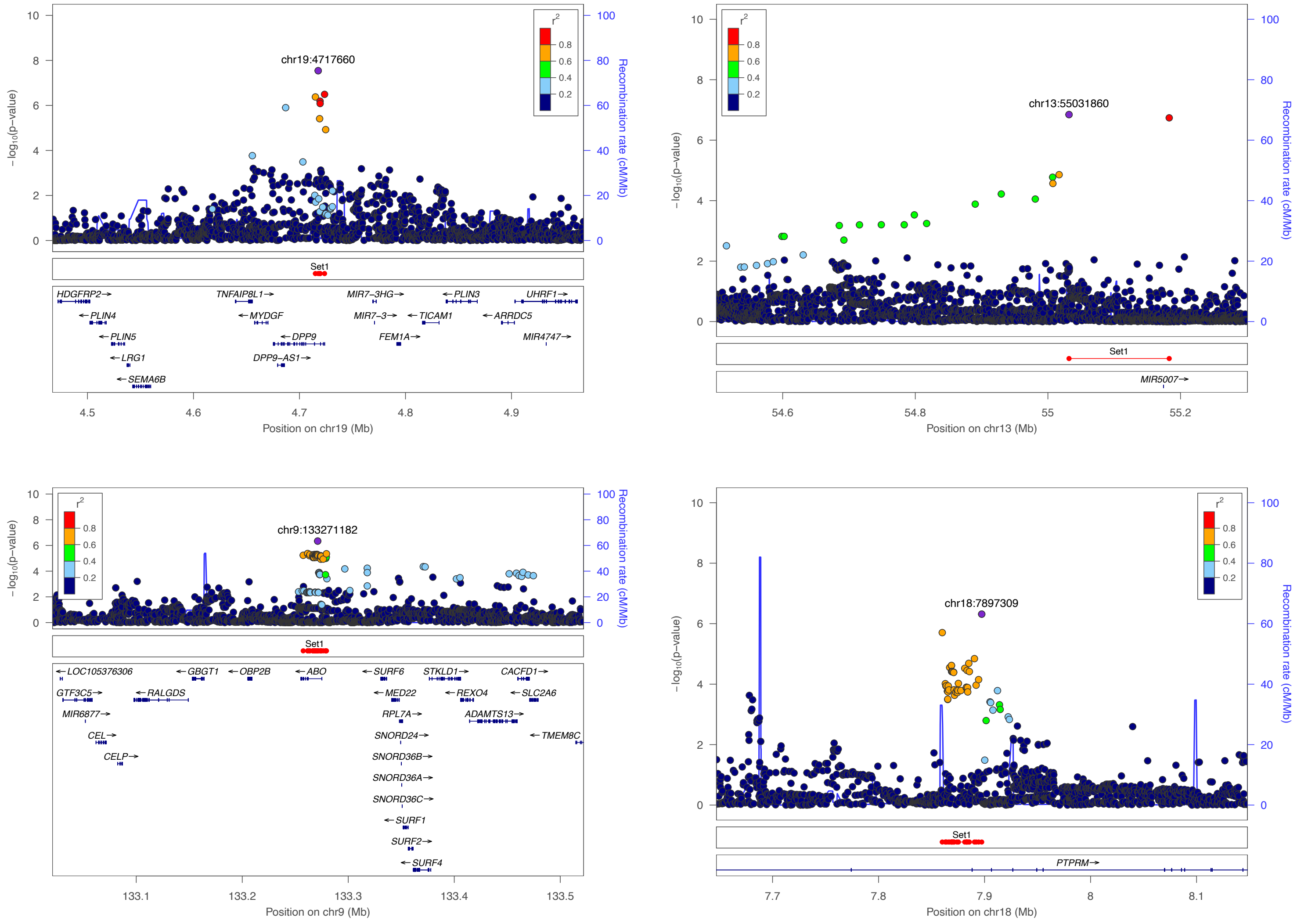
chr19:4717660
−log10(p−value)
Recombination rate (cM/Mb)
r2
0.8
0.6
0.4
0.2
Set1
HDGFRP2
PLIN4
PLIN5
LRG1
SEMA6B
TNFAIP8L1
MYDGF
DPP9
DPP9−AS1
MIR7−3HG
MIR7−3
FEM1A
PLIN3
TICAM1
ARRDC5
UHRF1
MIR4747
Position on chr19 (Mb)
chr13:55031860
MIR5007
Position on chr13 (Mb)
chr9:133271182
LOC105376306
GTF3C5
MIR6877
CEL
CELP
RALGDS
GBGT1
OBP2B
ABO
SURF6
MED22
RPL7A
SNORD24
SNORD36B
SNORD36A
SNORD36C
SURF1
SURF2
SURF4
STKLD1
REXO4
ADAMTS13
CACFD1
SLC2A6
TMEM8C
Position on chr9 (Mb)
chr18:7897309
PTPRM
Position on chr18 (Mb)

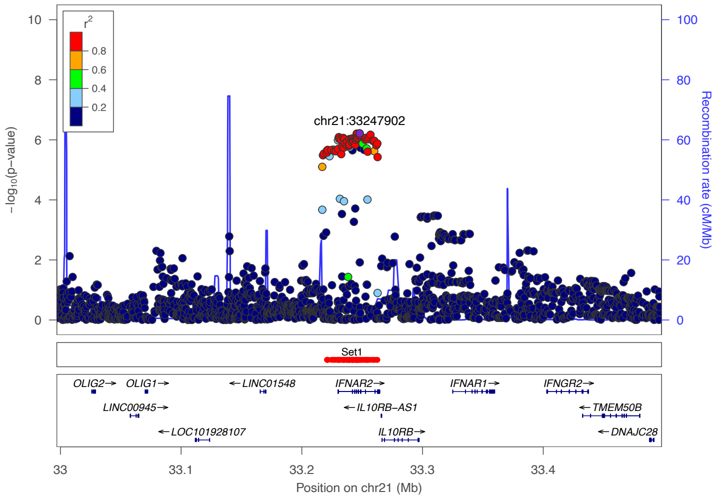

chr21:33247902
−log10(p−value)
Recombination rate (cM/Mb)
Set1
OLIG2
OLIG1
LINC00945
LOC101928107
LINC01548
IFNAR2
IL10RB−AS1
IL10RB
IFNAR1
IFNGR2
TMEM50B
DNAJC28
Position on chr21 (Mb)

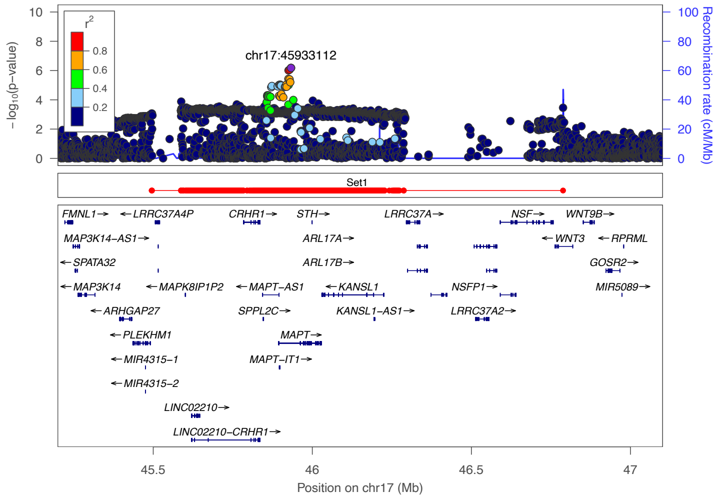

chr17:45933112
−log10(p−value)
Recombination rate (cM/Mb)
Set1
FMNL1
LRRC37A4P
MAP3K14−AS1
SPATA32
MAP3K14
ARHGAP27
PLEKHM1
MIR4315−1
MIR4315−2
LINC02210
LINC02210−CRHR1
MAPK8IP1P2
CRHR1
STH
ARL17A
ARL17B
MAPT−AS1
SPPL2C
MAPT
MAPT−IT1
KANSL1
KANSL1−AS1
LRRC37A
NSFP1
LRRC37A2
NSF
WNT3
WNT9B
RPRML
GOSR2
MIR5089
Position on chr17 (Mb)

**Supplementary Figure 6. Manhattan and quantile-quantile plots from the meta-analysis of the first analysis and the COVID-19 HGI B2 analysis release 5 summary association statistics (IIc).** Shown are Manhattan **(a)** and **(b)** quantile-quantile (QQ) plots of the association statistics from the meta-analysis (IIc) of the first analysis and the COVID-19 HGI B2 analysis release 5 (**Supplementary Methods**). The red dashed line indicates the genome-wide significance threshold of a P $<5\times10^{-8}$, the blue dashed line indicates a suggestive threshold of P $<10^{-6}$. Only markers that passed the imputation score $R^2\geq0.6$ and had a MAF≥1% were used for plotting. In QQ plot, the 2.5th and 97.5th centiles of the distribution under random sampling and the null hypothesis form the 95% concentration band. The genomic inflation factor lambda (λ) is defined as the ratio of the medians of the sample χ2 test statistics and the 1-d.f. χ2 distribution (0.455).(91) Red dots indicate variants with meta-analysis heterogeneity P-value of < $10^{-5}$ or variants with a meta-analysis heterogeneity P-value < 0.001 in the COVID-19 HGI genetics consortium B2 analysis release 5.

**(a)**

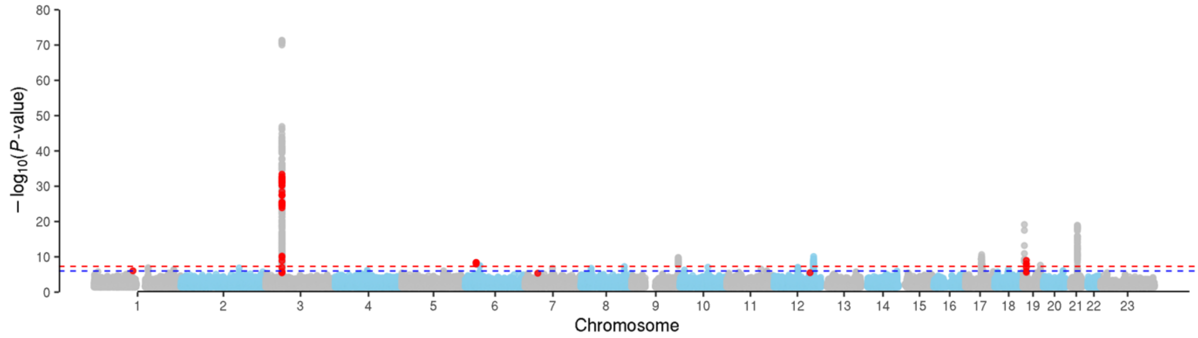

**(b)**

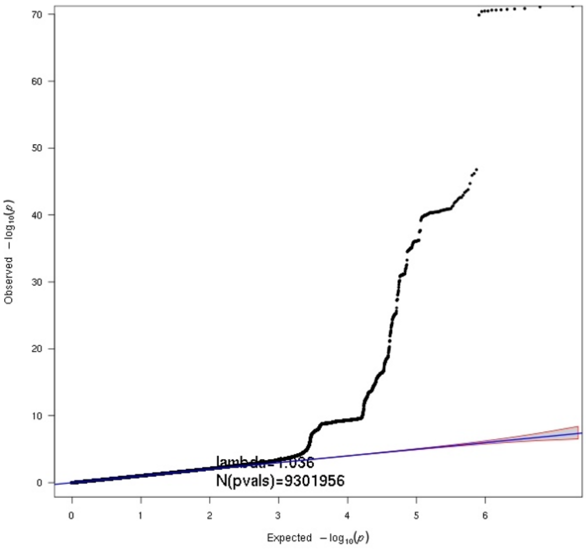

**Supplementary Figure 7. Manhattan and quantile-quantile plots from the meta-analysis of the second analysis and the COVID-19 HGI A2 analysis release 5 summary association statistics (IId).** Shown are Manhattan **(a)** and **(b)** quantile-quantile (QQ) plots of the association statistics from the meta-analysis (IId) of the second analysis and the COVID-19 HGI A2 analysis release 5 (**Supplementary Methods**). The red dashed line indicates the genome-wide significance threshold of a P <5×10$^{-8}$, the blue dashed line indicates a suggestive threshold of P <10$^{-6}$. Only markers that passed the imputation score R²≥0.6 and had a MAF≥1% were used for plotting. In QQ plot, the 2.5th and 97.5th centiles of the distribution under random sampling and the null hypothesis form the 95% concentration band. The genomic inflation factor lambda (λ) is defined as the ratio of the medians of the sample χ2 test statistics and the 1-d.f. χ2 distribution (0.455).(91) Red dots indicate variants with meta-analysis heterogeneity P-value of < 10$^{-5}$ or variants with a meta-analysis heterogeneity P-value < 0.001 in the COVID-19 HGI genetics consortium A2 analysis release 5.

**(a)**

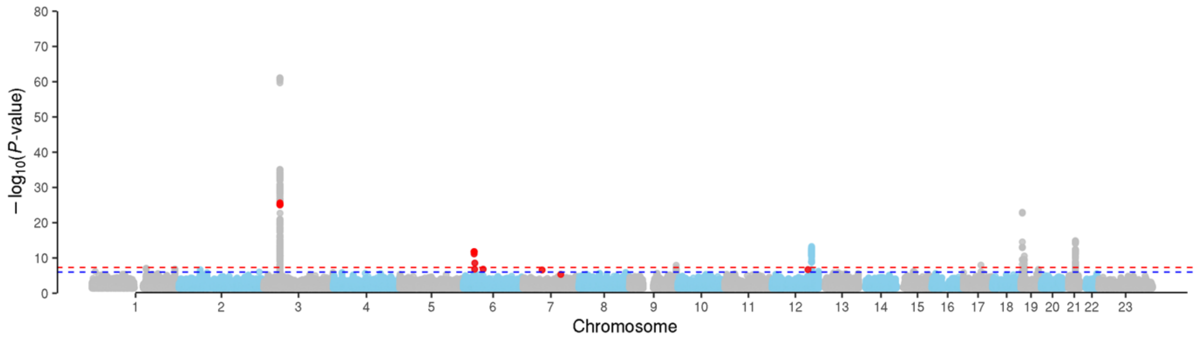

**(b)**

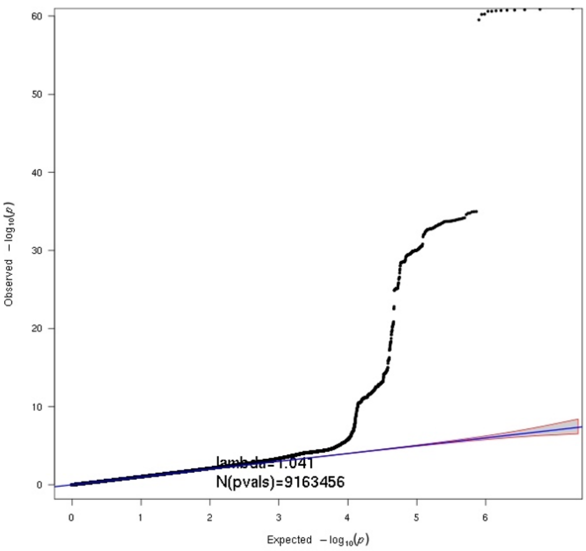

**Supplementary Figure 8. Regional association plot for the 19q13.33 locus.**

Plot was created using the LocusZoom tool.(92) LD values were calculated based on genotypes of the merged Italian/Spanish/Norwegian/German/Austrian dataset derived from TOPMed imputation (**Online Methods**). Genome build hg38 positions are plotted. The recombination rate is shown in centimorgans (cM) per million base pairs (Mb). The plot shows the names and locations of the genes; the transcribed strand is indicated with an arrow. Genes are represented with intronic and exonic regions. The purple diamond in each panel represents the variant most strongly associated with severe COVID-19 and respiratory failure. Set1 shows the 95% credible set from Bayesian fine mapping (**Online Methods**).

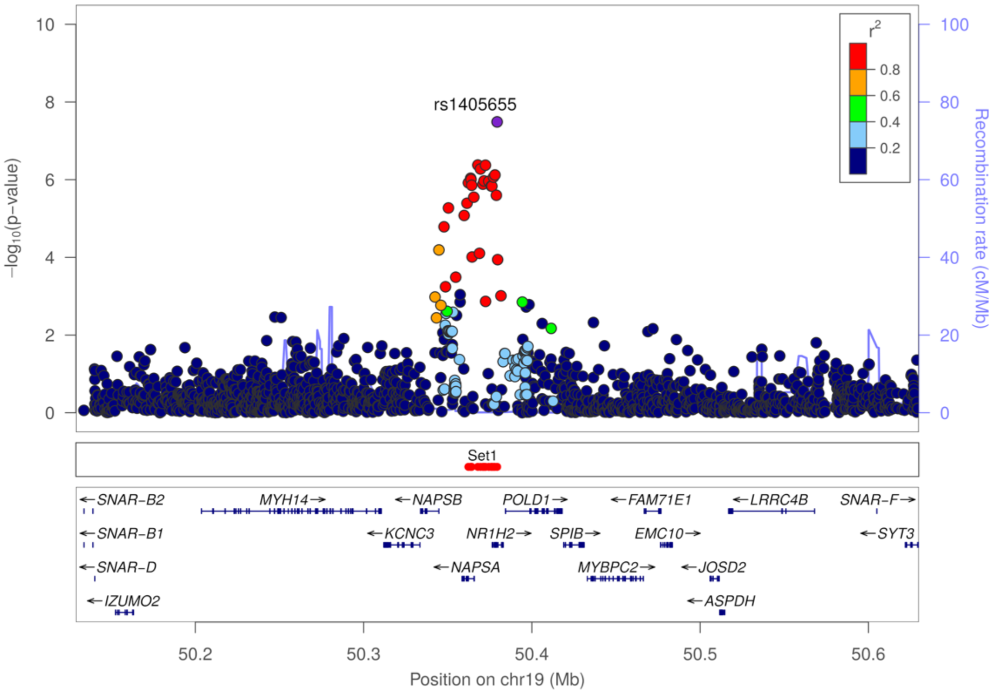

**Supplementary Figure 9: Forest plot of candidates from the in-depth stratified analysis.** Odds ratio (OR) and 95% confidence intervals (CIs) of the main (MAIN) analysis and the stratified analyses, each substratified by sex. Only cohorts in which N>50 in both cases and controls are shown. Only the first analysis is shown.

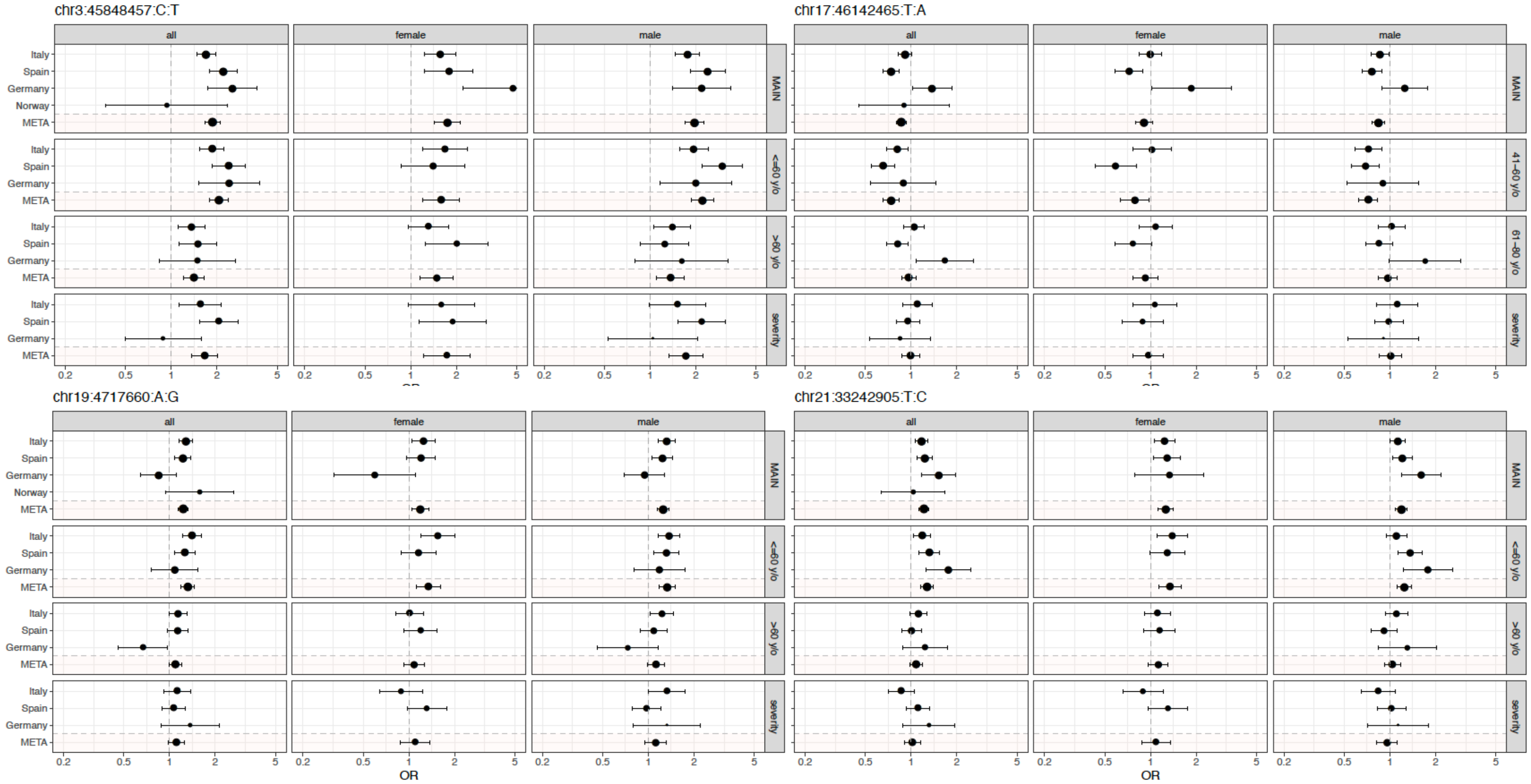

**Supplementary Figure 10: Forest plot of candidates from the in-depth stratified analysis.** Odds ratio (OR) and 95% confidence intervals (CIs) of the main (MAIN) analysis and the stratified analyses, each substratified by sex. Only cohorts in which N>50 in both cases and controls are shown. Only the second analysis is shown.

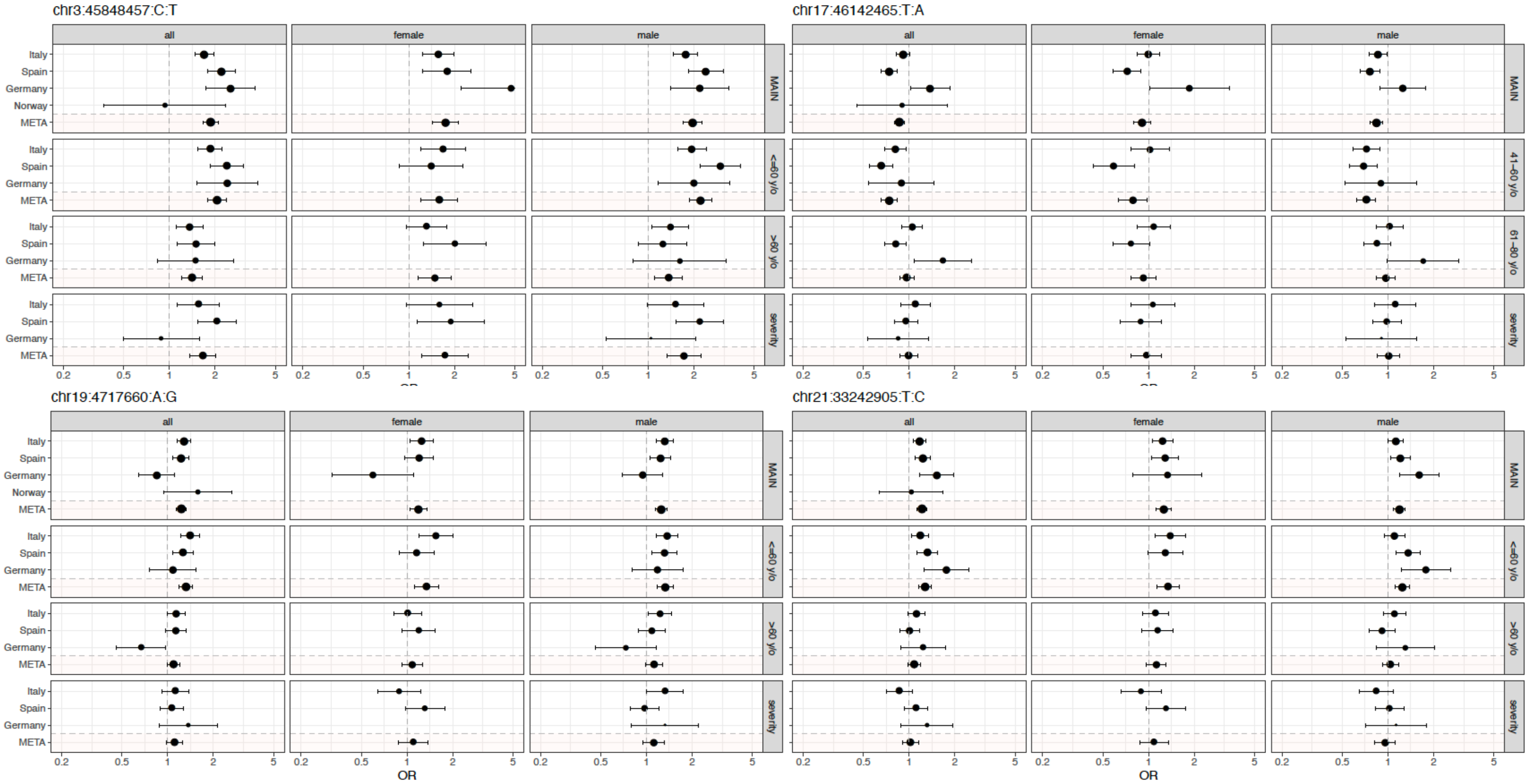

**Supplementary Figure 11. Summary of the 17q21.31 and 19q13.33 loci tissue-specific effects on gene-expression (eQTL) and splicing (sQTL) from GTEx data.**

Extended figure in analogy to **Figure 2a** in the main manuscript. eQTL and sQTL analysis of all candidate genes associated with the 17q.21.31 and 19q.13.33 loci in GTEx(19) tissues. Black rectangles indicate genes for which the inversion haplotype (here using the tag SNP rs62055540-C, LD ($r^2$)=1), the 19q.13.33 allele (rs1405655-C) or any proxy variants ($r^2$>0.9) of these two SNPs are lead QTL variants in that tissue (**Supplementary Table 11**). If no box is shown other variants (i.e. not rs62055540-C, rs1405655-C or their proxy variants) are lead QTLs. Estimates and P-values were taken from the GTEx Catalog.(19) Direction of eQTL and sQTL NES are represented by color intensities, while statistical significance is represented by dot size. Both loci are lead *cis*-eQTL and/or sQTL of several genes in different tissues, thus have the strongest effect on their expression (eGenes) and splicing (sGenes), respectively.

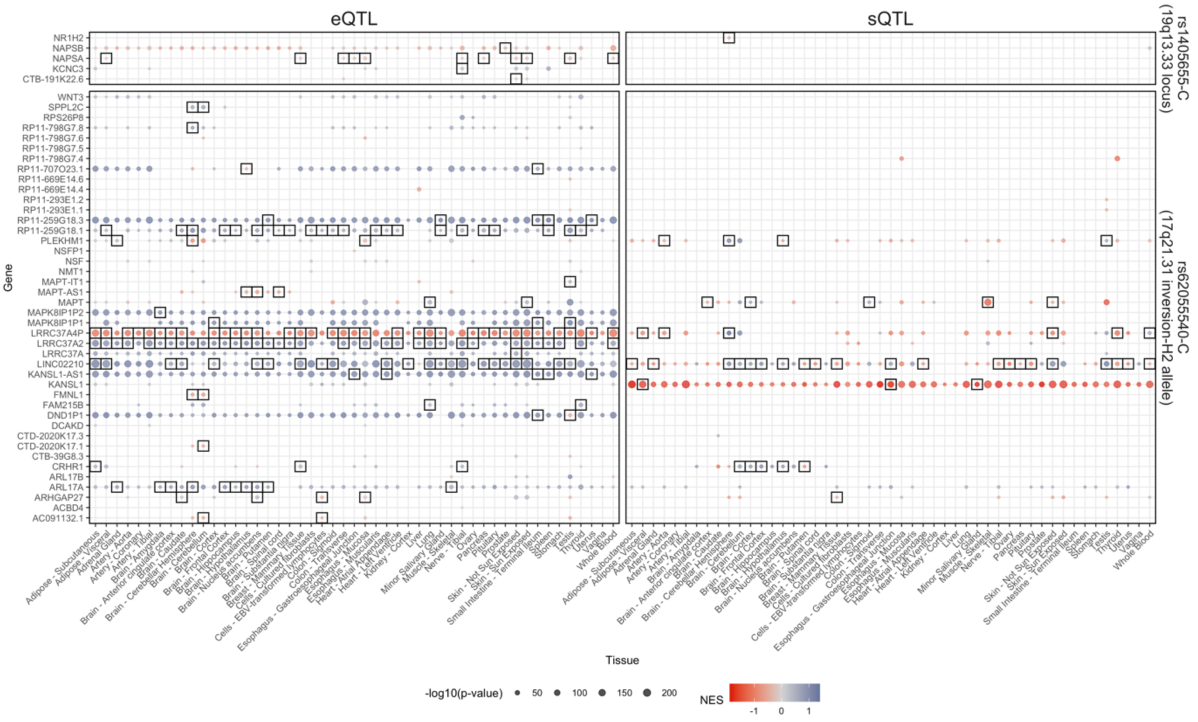

**Supplementary Figure 12. Expression levels of candidate genes of genome-wide significant loci in different tissues, immune cell types and lung as well as brain single cell data.**

Chromosome regions 17q21.31 and 19q13.33 span or/and are associated with expression changes of several protein-coding genes. To identify most plausible candidates, we performed exploratory gene expression analysis on publicly available bulk as well as sc-RNA-Seq datasets of main organ and COVID-19 relevant tissues and cells. Description of how candidate genes were chosen is described in the **Supplementary Methods**. **(a)** Figure shows immune cell-type and tissue bulk mRNA gene expression results from the BLUEPRINT and GTEx consortia, respectively. Visualized expression values were gene-wise centered, and z-score normalized, thus showing in which tissues a particular gene is mostly enriched; **(b)** Figure panels represent log-normalized mean expression (visualized by color) of candidate genes and fraction of cells expressing those genes (visualized by the size of the dot). Lung and upper airway sc-RNA-Seq data from Vieira Braga *et al.*(24) contain mRNA expression levels in healthy nasal, bronchi, alveoli and parenchyma cells, whereas brain sc-RNA-Seq data from Habib *et al.*(25) contain mRNA expression levels in adult human brain cells from recently deceased, non-diseased donors. Processed and cell type annotated sc-RNA-Seq datasets from both studies were retrieved from COVID-19 Cell Atlas.(26) **(c)** Figure shows differential expression of candidate genes in COVID-19 lung cells compared to healthy controls. Log2 fold change (log2FC) values are presented as color gradient, while the nominal P-values in –log10 scale are shown proportionally to a dot size. Black-bordered circles indicate significantly differentially expressed genes after FDR correction. The results were obtained from pseudo-bulk differential expression analysis performed by Delorey *et al.*(30) Genes missing from one of the plots in b) and c) were not available for the respective datasets. The *MAPT-IT* gene is here named *STH.*

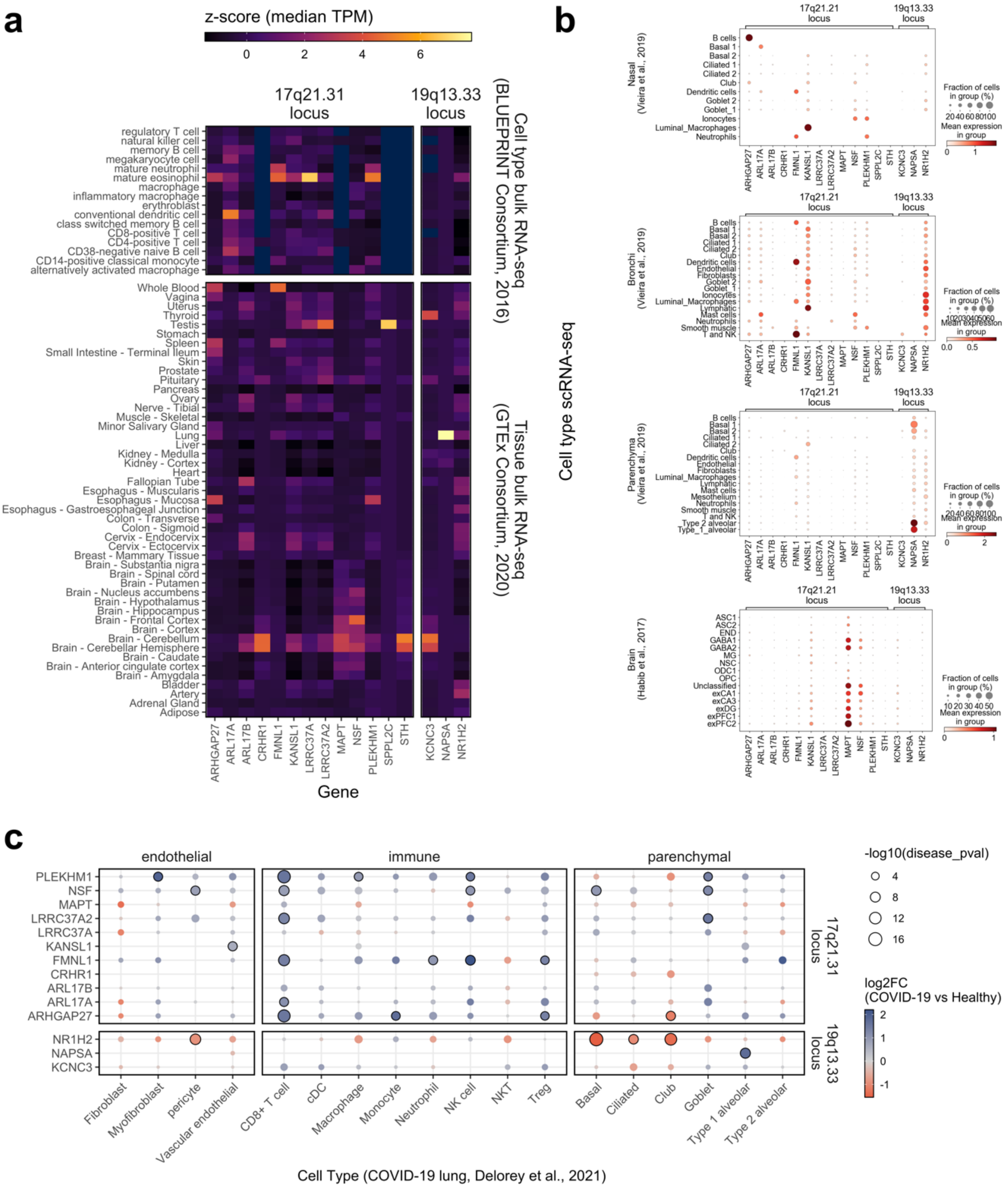
a
z-score (median TPM)
0
2
4
6
17q21.31 locus
19q13.33 locus
Cell type bulk RNA-seq (BLUEPRINT Consortium, 2016)
Tissue bulk RNA-seq (GTEx Consortium, 2020)
regulatory T cell
natural killer cell
memory B cell
megakaryocyte cell
mature neutrophil
mature eosinophil
macrophage
inflammatory macrophage
erythroblast
conventional dendritic cell
class switched memory B cell
CD8-positive T cell
CD4-positive T cell
CD38-negative naive B cell
CD14-positive classical monocyte
alternatively activated macrophage
Whole Blood
Vagina
Uterus
Thyroid
Testis
Stomach
Spleen
Small Intestine - Terminal Ileum
Skin
Prostate
Pituitary
Pancreas
Ovary
Nerve - Tibial
Muscle - Skeletal
Minor Salivary Gland
Lung
Liver
Kidney - Medulla
Kidney - Cortex
Heart
Fallopian Tube
Esophagus - Muscularis
Esophagus - Mucosa
Esophagus - Gastroesophageal Junction
Colon - Transverse
Colon - Sigmoid
Cervix - Endocervix
Cervix - Ectocervix
Breast - Mammary Tissue
Brain - Substantia nigra
Brain - Spinal cord
Brain - Putamen
Brain - Nucleus accumbens
Brain - Hypothalamus
Brain - Hippocampus
Brain - Frontal Cortex
Brain - Cortex
Brain - Cerebellum
Brain - Cerebellar Hemisphere
Brain - Caudate
Brain - Anterior cingulate cortex
Brain - Amygdala
Bladder
Artery
Adrenal Gland
Adipose
ARHGAP27
ARL17A
ARL17B
CRHR1
FMNL1
KANSL1
LRRC37A
LRRC37A2
MAPT
NSF
PLEKHM1
SPPL2C
STH
KCNC3
NAPSA
NR1H2
Gene
b
Cell type scRNA-seq
17q21.21 locus
19q13.33 locus
Nasal (Vieira et al., 2019)
Bronchi (Vieira et al., 2019)
Parenchyma (Vieira et al., 2019)
Brain (Habib et al., 2017)
Fraction of cells in group (%)
Mean expression in group
c
endothelial
immune
parenchymal
-log10(disease_pval)
4
8
12
16
log2FC (COVID-19 vs Healthy)
2
1
0
-1
17q21.31 locus
19q13.33 locus
PLEKHM1
NSF
MAPT
LRRC37A2
LRRC37A
KANSL1
FMNL1
CRHR1
ARL17B
ARL17A
ARHGAP27
NR1H2
NAPSA
KCNC3
Fibroblast
Myofibroblast
pericyte
Vascular endothelial
CD8+ T cell
cDC
Macrophage
Monocyte
Neutrophil
NK cell
NKT
Treg
Basal
Ciliated
Club
Goblet
Type 1 alveolar
Type 2 alveolar
Cell Type (COVID-19 lung, Delorey et al., 2021)

**Supplementary Figure 13. 17q21.31 inversion is a lead eQTL variant for several genes in activated monocytes.** Figure shows gene expression by genotype of the inversion in differently stimulated monocyte populations (NS – non-stimulated; Pam3CSK4 – Pam3CysSerLys4-stimulated. Pam3CysSerLys4 mimics the acylated amino terminus of bacterial lipopeptides; LPS – lipopolysaccharide; R848 – Resiquimod (Resiquimod is an agonist of Toll-like receptors 7 and 8); IAV – human influenza A virus). Barplots represent averages and of log-normalized TPM values across at total 200 NS samples, 184 LPS samples, 196 Pam3CSK4 samples, 191 R848 samples, and 199 IAV samples respectively, derived from 100 European and 100 African individuals in total.(35) Barplots are grouped by genotype (indicated by color). Standard errors are shown as error bars. Significance levels of lead associations: $**P_{FDR} < 0.01$; $***P_{FDR} < 0.001$ and $****P_{FDR} < 0.0001$. The dataset was obtained from Quach *et al.*(35) (accession EGA:EGAS00001001895). Based on the inversion genotype and the expression data, we performed an eQTL analysis (**Supplementary Methods**).

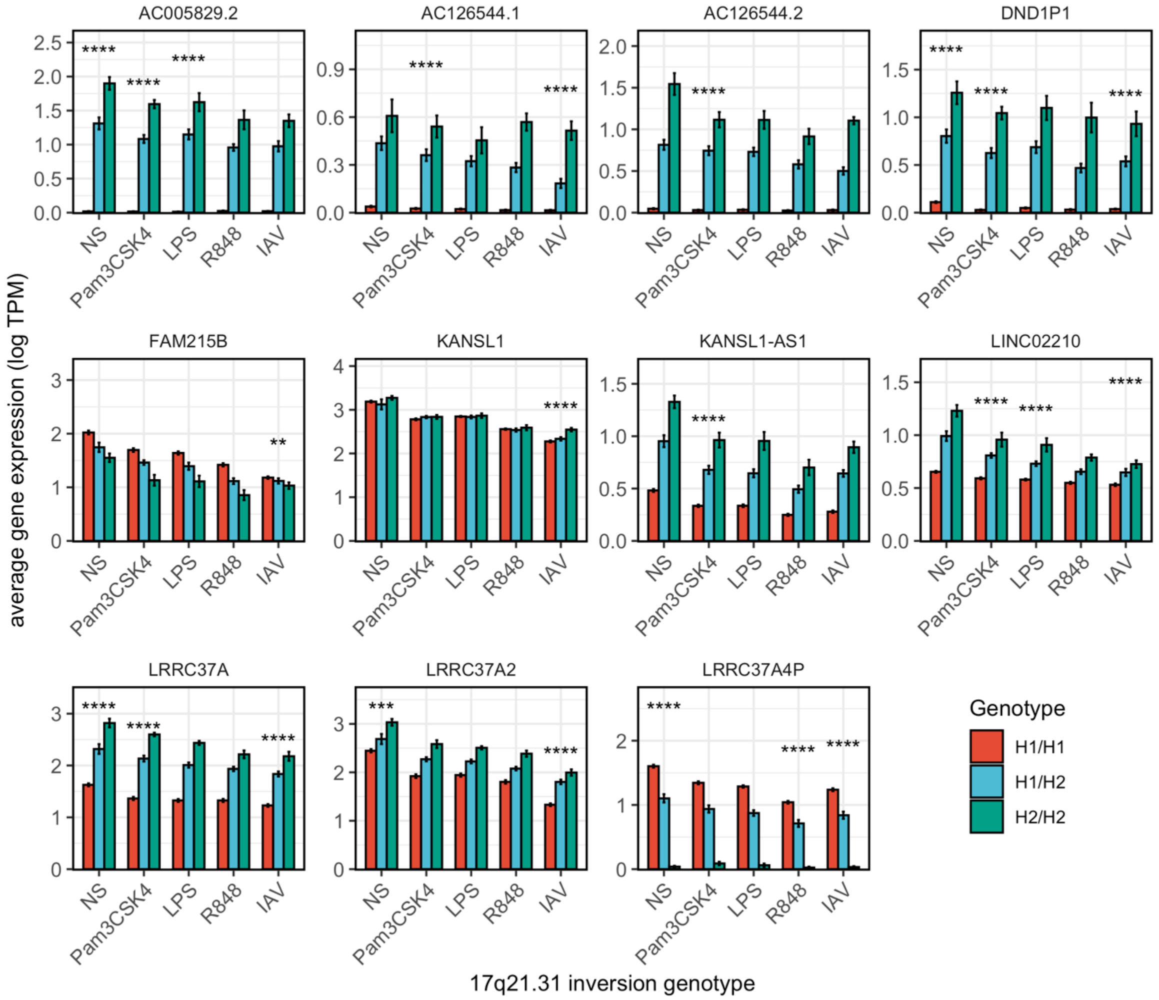

**Supplementary Figure 14. 17q21.31 inversion impact on *KANSL1* expression in monocytes.**

**(a)** Schematic representation of two *KANSL1* isoforms out of the 36 described by the GENCODE Project (v36). ENST00000572904.6 is the most expressed protein-coding transcript, whereas ENST00000639356.1 is the non-coding isoform that most change its expression in inverted H2 chromosomes. UTR exons are depicted as light blue boxes and coding sequence in dark blue, with the arrow indicating the transcription direction. Legend shows the different scale used to represent the size of exons and introns. **(b)** Boxplots of expression levels of different protein-coding and non-coding *KANSL1* isoforms by inversion genotype in non-stimulated and stimulated monocytes with Influenza A virus (IAV) from ~200 individuals from European and African origin. Significance levels of lead associations: *$P_{FDR} < 0.05$; **$P_{FDR} < 0.01$ and ***$P_{FDR} < 0.001$. The dataset was obtained from Quach *et al.*(35) (accession EGA:EGAS00001001895).

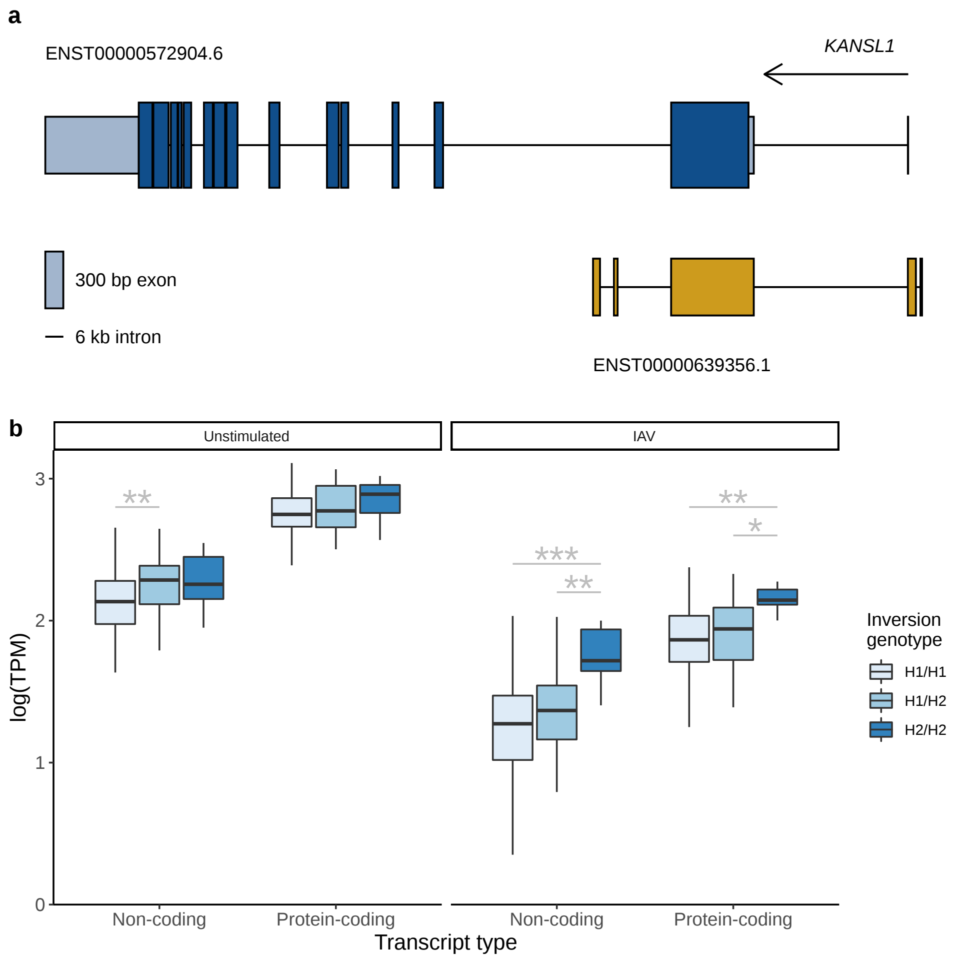

**Supplementary Figure 15. Differential expression analysis of GWAS candidate genes in SARS-CoV-2 infected human brain organoid scRNA-seq data.** Due to the fact that SARS-CoV-2 is capable to infect neural cells and that many of the GWAS candidate genes are enriched in neural tissues and cells, differential expression of candidate genes was explored in SARS-CoV-2 infected human brain organoid cell data, obtained from Song *et al.*(28) **(a)** Figure shows log-normalized mean expression (presented by color) of candidate genes by cell type and cell fraction expressing those genes (presented by the size of a dot), where x-axis displays time (in hours) after brain cell infection with SARS-CoV-2, while y-axis represents candidate genes of each GWAS locus. **(b)** The figure displays results of differential expression analysis in the SARS-CoV-2 infected neural cells compared to mock (non-infected) neural cells after 96 hours of infection. Values on the x-axis depict gene expression fold changes in logarithmic scale (log2 FC), while values on the y-axis present statistical significance (negative base 10 logarithm of p-value) of a change in gene expression. Only the candidate genes that were differentially expressed (PFDR<0.01 and |log2FC| > 0.1) are annotated using gene symbols in red.

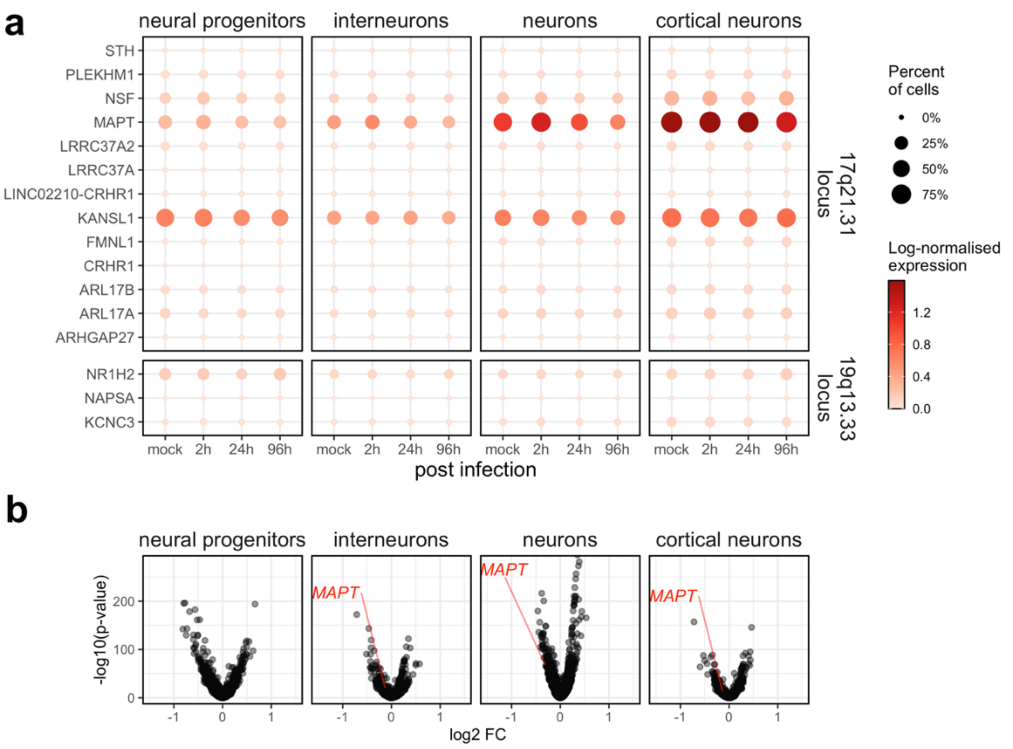

s

**Supplementary Figure 16. Regional association plot of the extended HLA region (chr6:25-35Mb).**

Regional association plot of the extended HLA region for the meta-analysis between cases and the general population **(a)** and disease severity **(b)**, and between severe cases and the general population **(c)** across the four cohorts. SNPs from the genome-wide array are shown as light-blue circles, imputed amino acids and nucleotide variants at the HLA loci as orange diamonds, and classical HLA alleles (at both 1st and 2nd field resolution) are shown as dark blue triangles. There were no association signals meeting either the genome-wide (red dashed line) or the suggestive association significance threshold of $P=1x10^{-5}$ (orange dashed line). The location of the classical HLA class I and class II genes are shown, together with the non-classical loci HLA-*E* and HLA-*G.*

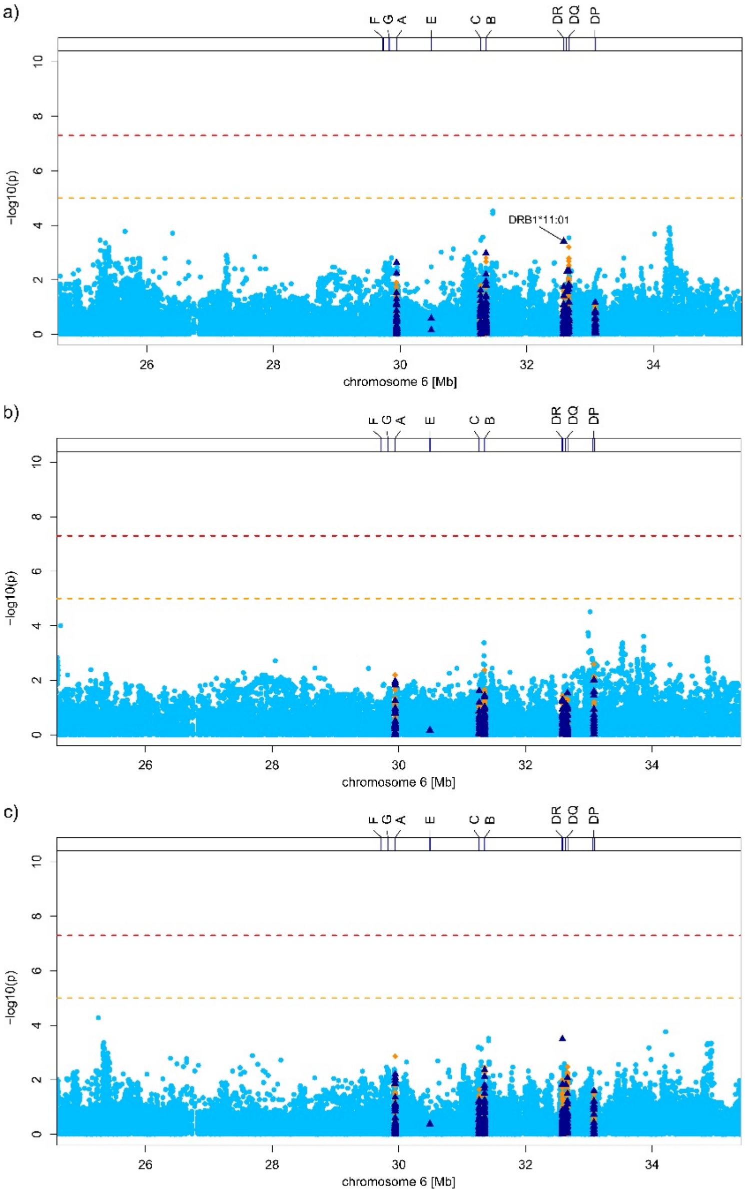

**Supplementary Figure 17. PepWAS results for disease risk, i.e. all cases vs. the general population, shown for the Spanish cohort ($N_{Cases}$=1,416, / $N_{Controls}$=4,382).**

Dashed red lines represent the peptidome-wide significance threshold using Bonferroni correction. For better visualization, a few peptides with outlier effect sizes were removed during plotting, none of which had a P-value that exceeded the significance threshold.

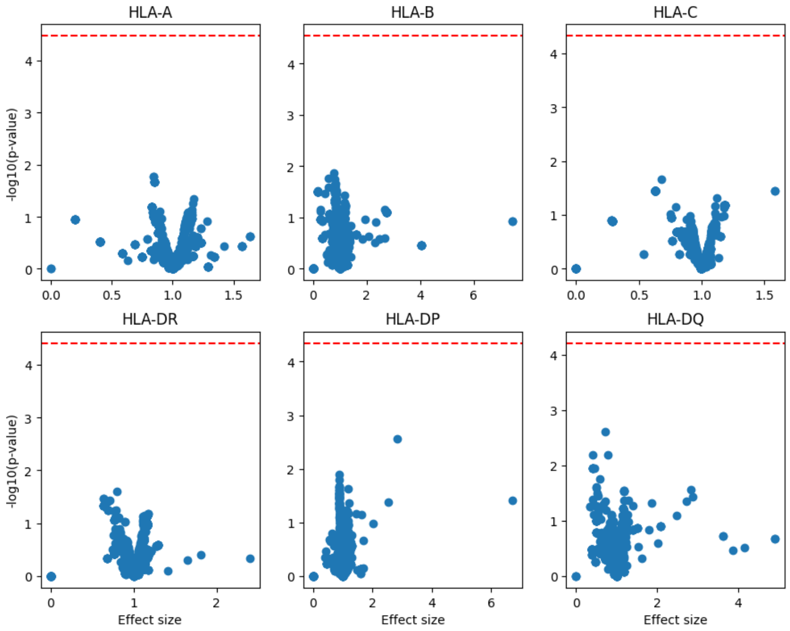

**Supplementary Figure 18. PepWAS results for disease severity, i.e. mild cases vs. severe cases, shown for the Spanish cohort ($\mathbf{N_{resiratory\_support\_status1}}$=897/ $\mathbf{N_{respiratory\_support\_status2\text{-}4}}$=519).**

Dashed lines represent the peptidome-wide significance threshold using Bonferroni correction. For better visualization, a few peptides with outlier effect sizes were removed during plotting, none of which had a P-value that exceeded the significance threshold.

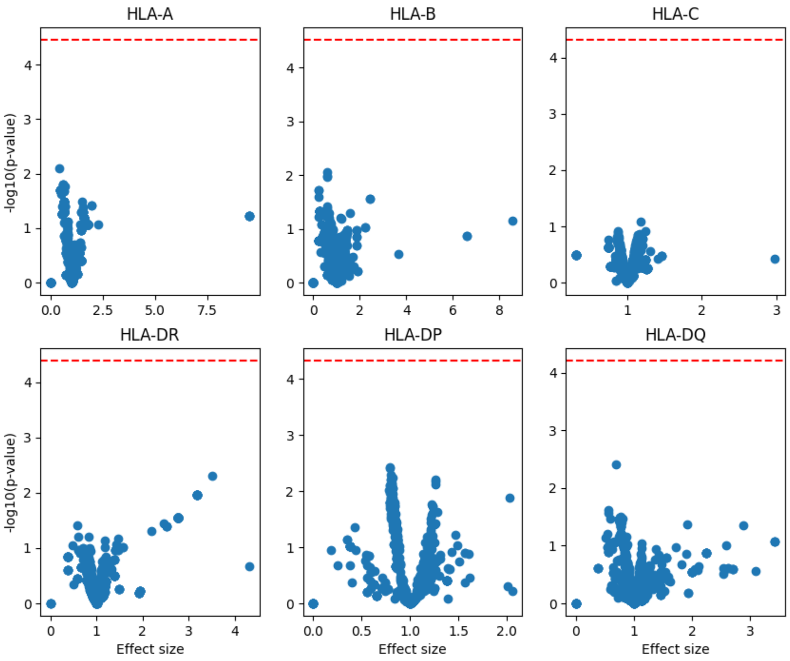

#### Supplementary Tables

**Supplementary Table 1**. Patient and control GWAS panels before and after quality control. Institutional review board and ethnics approval ids for each center.

a) Total number of patients from each center with defined case/control status before QC

| | Hospitals or research institution/project | patients/center | |
|---|---|---|---|
| | | cases | controls |
| NORWAY | TOTAL | 127 | 288 |
| | 1. Oslo University Hospital, Oslo | | |
| | 2. Vestre Viken Hospital Trust, Drammen | | |
| | 3. Østfold Hospital Trust, Kalnes | | |
| | 4. University Hospital of North Norway, Tromsø | | |
| | 5. St. Olav's University Hospital, Trondheim | | |
| | 6. Nord-Trøndelag Hospital Trust, Levanger | | |
| | 7. Møre og Romsdal Hospital Trust, Molde | | |
| | 8. Møre og Romsdal Hospital Trust, Ålesund | | |
| ITALY | TOTAL | 1,857 | 5,247 |
| | 1. Fondazione IRCCS Casa Sollievo della Sofferenza, San Giovanni Rotondo | - | 396 |
| | 2. Fondazione IRCCS Ca'Granda Ospedale Maggiore Policlinico, Milan | 1,232 | 3,198 |
| | 3. UNIMIB School of Medicine, San Gerardo Hospital, Monza | 350 | - |
| | 4. E.O. Galliera Hospital, Genoa | 48 | - |
| | 5. Humanitas Clinical Research Center, IRCCS, Milan | 227 | 1,653 |
| SPAIN | TOTAL | 2,795 | 4,552 |
| | 1. Donostia University Hospital, Donostia and Donostia Basque Biobank, Donostia | 1,169 | 994 |
| | 2. Hospital Clínic and IDIBAPS, Barcelona | 161 | - |
| | 3. Hospital Ramón y Cajal, Madrid | 298 | - |
| | 4. Hospital Universitario Vall d'Hebron, Barcelona | 489 | - |
| | 5. Hospital Universitario Virgen del Rocío, Sevilla & Hospital Universitario San Cecilio, Granada | 166 | - |
| | 6. GCAT. Genomes For Life. Germans Trias i Pujol Research Institute (IGTP), Barcelona | 512 | 2,368 |
| | 7. Vall d'Hebron Research Institute, Barcelona | - | 1,190 |
| GERMANY/AUSTRIA | TOTAL | 449 | 3,582 |
| | 1. Charité Unversitätsmedizin Berlin, Berlin, Germany | 66 | - |
| | 2. University Hospital Frankfurt, Frankfurt, Germany | 46 | - |
| | 3. Technical University Munich, Munich, Germany; München Klinik Schwabing, Munich, Germany; (COMRI study) | 54 | - |
| | 4. Research Center Borstel, Borstel, Germany | 5 | - |
| | 5. University Medical Center Schleswig-Holstein, Kiel, Germany | 20 | - |
| | 6. University Medical Center, Schleswig-Holstein, Lübeck, Germany | - | 3,582 |
| | 7. University Hospital Regensburg, Regensburg, Germany | 65 | - |
| | 8. Medical University of Innsbruck, Innsbruck, Austria | 35 | - |
| | 9. University Hospital Bonn, Bonn, Germany; (BoSCO study) | 148 | - |
| | 10. University Hospital Cologne, Cologne, Germany | 10 | - |

b) Recruiting Centers and Ethics Committee approval IDs.

| Center | Review Board | Reference |
|---|---|---|
| Donostia University Hospital, Donostia and Donostia, Basque Biobank, Donostia | Ethics Committee for research with Medicines of the Basque Country (CEIm-E) | PI2020064 |
| Hospital Clinic and IDIBAPS, Barcelona | Ethics Committee of Hospital Clinic, Barcelona, Spain | HCB/2020/0405 and HCB/2020/1300 |
| Hospital Ramón y Cajal, Madrid | Ethics Committees for Investigation (CEI) Hospital Ramón y Cajal, Madrid | 093/20 |
| Hospital Universitari Vall d'Hebron, Barcelona | Ethics Committee of Vall Hebron Hospital, Barcelona, Spain | PR[AG]244/2020 |
| Hospital Universitario Virgen del Rocío, Sevilla & Hospital Universitario San Cecilio, Granada | Ethics Committees for Investigation (CEI) de los Hospitales Universitarios Virgen Macarena y Virgen del Rocío | 1954-N-20 & 0886-N-20 |
| GCAT. Genomes For Life. Germans Trias i Pujol Research Institute (IGTP), Barcelona | Germans Trias i Pujol University Hospital Research Ethics Committee | IRB00002131- / PI-13-020 / PI-20-182 |
| Vall d'Hebron Research Institute, Barcelona | Ethics Committee of Vall Hebron Hospital, Barcelona, Spain | 20/0022 |
| Fondazione IRCCS Casa Sollievo della Sofferenza, San Giovanni Rotondo | Ethical Committee of the Fondazione IRCCS Casa Sollievo della Sofferenza, San Giovanni Rotondo | 12701/08 |
| Fondazione IRCCS Ca'Granda Ospedale Maggiore Policlinico, Milan | Ethics Committee MILANO AREA 2, Fondazione IRCCS Ca' Granda Ospedale Maggiore Policlinico, Milan | 342_2020 and 342_2020bis for cases, 334_2020 and 334_2020bis for controls |
| UNIMIB School Of Medicine, San Gerardo Hospital, Monza | Ethics Committee of the National Institute of Infectious Diseases Lazzarro Spallanzani, Monza, Italy | 84/2020 |
| E.O. Galliera Hospital, Genoa | Ethics Committee of the Liguria Region (CER), Italy | 237/2020-DB id 10580 |
| Humanitas Clinical and Research Center, IRCCS, Milan | Independent Ethics Committee of the IRCCS Istituto Clinico Humanitas, Rozzano (Milan),<br>Ethics Committee of MILANO AREA 2 (ASST Centro Specialistico Ortopedico Traumatologico Gaetano Pini-CTO, Milan) | 316/20, 483 |
| Charité, Universitätsmedizin Berlin, Berlin, Germany | Ethics Committee of Charite Universitätsmedizin Berlin, Berlin, Germany | EA2/066/20 |
| University Hospital Frankfurt, Frankfurt, Germany | Institutional Review board of the University Hospital Frankfurt, Frankfurt, Germany | 11/17 |

| | | |
|---|---|---|
| Technical University Munich, Munich, Germany; München Klinik Schwabing, Munich, Germany | Ethics Committee of the Technical University Munich, Munich, Germany | TUM 217/20S, TUM 221/20S, TUM 440/20S |
| Research Center Borstel, BioMaterialBank Nord, Germany | Ethics Committee of the University of Lübeck, Lübeck, Germany | AZ 20-384 |
| University Medical Center, Schleswig Holstein, Kiel, Germany | Institutional Review board of the Medical Faculty of Kiel University, Kiel, Germany | B231/98 & Broad Consent |
| University Medical Center, Schleswig Holstein, Lübeck, Germany | Institutional Review board of the Medical Faculty of Kiel University, Kiel, Germany | AZ A103/14 |
| University Hospital Regensburg, Regensburg, Germany | Ethics Committee at the University of Regensburg, Regensburg, Germany | 20-1785-101 |
| Medical University and University Hospital of Innsbruck, Innsbruck Austria | Ethics Committee of the Medical University of Innsbruck, Innsbruck, Austria | AN1107/2020 |
| University Hospital Bonn and School of Medicine, University of Bonn, Bonn, Germany (part of BoSCO) | Ethics Committee of the Medical Faculty Bonn, Bonn, Germany | 171/20 |
| Aachen study on COVID-19 (part of BoSCO) | Ethics Committee of the RWTH Aachen, Aachen, Germany | EK 080 / 20 (CTC-A-Nr. 20-085) |
| CORSAAR study (part of BoSCO) | Ethics Committee of the Medical Board of the Saarland, Germany | 61/20 |
| Department of Anaesthesiology and Intensive Care Medicine, University Hospital Essen, University Duisburg-Essen, Germany (part of BoSCO) | Ethics Committee of the University Duisburg-Essen, Germany | 21-9900-BO |
| Düsseldorf Biobank, Department Gastroenterology, Hepatology and Infectious Diseases (part of BoSCO) | Ethics Committee of the Medical Faculty Duesseldorf, Duesseldorf, Germany | 3530 |
| Hannover Unified Biobank (part of BoSCO) | Ethics Committee of the Hannover Medical School (MHH), Hannover, Germany | 9001_BO_K |
| Recovery Cohort (part of BoSCO) | Ethics Committee of the University Hospital of Cologne, Cologne, Germany | 16–054/ 20–1187 |
| University Hospital of Cologne, Cologne, Germany | Ethics Committee of the University Hospital of Cologne, Cologne, Germany | 20-1295 |
| Oslo University Hospital, Oslo; Vestre Viken Hospital Trust, Drammen; Østfold Hospital Trust, Kalnes; University; Hospital of North Norway, Tromsø; St. Olav's University Hospital, Trondheim; Nord-Trøndelag Hospital Trust, Levanger; Møre og Romsdal Hospital Trust, Molde; Møre og Romsdal Hospital Trust, Ålesund | Regional Committee for Medical and Health Ethics in South-Eastern Norway, Norway | 132550 |

c) Description of control panels

| Center | PMID | Comment |
|---|---|---|
| Italy 1 | 21102463 | Controls were selected from healthy volunteers and from gastroenterology outpatients recruited in IRCCS-Casa Sollievo della Sofferenza Hospital, San Giovanni Rotondo, Italy. Symptomatic controls with IBS and non IBD inflammation were also collected. |
| Italy 2 | 32558485 | Randomly recruited blood donors. |
| Italy 5 | 33209983 | Controls recruited among partners and caregivers of Parkinson's disease patients at the Parkinson Institute of Milan. Negative for neurodegenerative disorders and denied any family history for movement disorders in first-degree relatives. |
| Spain 1 | | Randomly recruited donors from the Basque Biobank. |
| Spain 6 | 30166351, 29593016 | Cases and controls included in the study belong to the GCAT cohort, a population-based cohort of adult people aged 45-65, from Catalonia, in the North-East of Spain. Cohort Protocol and Genetic characterization have been previously reported. Cases and controls were defined by the linked Electronic Medical Records, from the Public Healthcare system, based on July's 2020 update. |
| Spain 7 | 30552173 | Vall d'Hebron Research Institute, Barcelona. Adult (>18yr old) healthy control individuals among blood bank donors, from >10 university hospitals from diverse provinces in Spain. Sampled for Spanish health control group in GWAS in autoimmune diseases. |
| Germany 6 | | Randomly recruited blood donors. |
| Norway 1 | | Healthy controls from Norway were randomly selected from the Norwegian Bone Marrow Donor registry. |

d) Overview of genotyped cases and controls from each country with reasons for exclusion during quality control (QC)

| | Italy | Spain | Norway | Germany/Austria |
|---|---|---|---|---|
| Pre-QC* totals | 7,104 | 7,347 | 415 | 4,067 |
| Pre-QC *(cases/controls) | 1,857/5,247 | 2,795/4,552 | 127/288 | 449/3,618 |
| *Sex mismatch/deleted in QC Ellinghaus *et al.*(44) | 323 | 412 | 2 | 55 |
| Pre-QC** totals | 6,791 | 6,935 | 413 | 4,012 |
| Pre-QC **(cases/controls) | 1,720/5,071 | 2,431/4,504 | 126/287 | 439/3,573 |
| QC details | | | 3 | |
| Missingness outliers | 27 | 6 | 4 | 14 |
| Heterozygosity outliers | 10 | 13 | 3 | 14 |
| PCA outliers | 239 | 262 | 45 | 150 |
| Duplicates | 42 | 24 | 0 | 94 |
| Relatives | 181 | 74 | 1 | 123 |
| Total unique removed*** | 306 | 293 | 49 | 257 |
| Post-QC totals | 6,322 | 6,583 | 364 | 3,639 |
| Post-QC (cases/controls) | 1,563/4,759 | 2,181/4,402 | 81/283 | 336/3,303 |

*Total number of available individuals including all individuals (pre-QC) from Ellinghaus, Degenhardt *et al.*(44) **Number of available subjects, including all individuals (post-QC) from post-QC Ellinghaus, Degenhardt et al. (44) and new samples. All quality control parameters now refer to **, ***The total number of unique samples removed from analysis is smaller than the sum of reasons for exclusion since some samples may have several.

**NOTE:** Post-QC individuals include: 85 German/Austrian, 12 Italian, 706 Spanish and 19 Norwegian individuals with a mild disease (defined as no respiratory support needed) or missing the information, 194 German, 26 Italian, 24 Spanish and 21 Norwegian individuals missing age information, 2 Norwegian individuals with missing sex. These were excluded for the analysis of severe respiratory failure

e) Detailed description of numbers of individuals included in this study.

SEE EXCEL TABLE

**Supplementary Table 2. Age and sex characteristics of controls**

| | Italy | Spain | Norway | Germany |
|---|---|---|---|---|
| First analysis & second analysis | | | | |
| Median age (IQR) — yr | 53 (23) | 52 (13) | 53 (11) | 50 (22) |
| Female sex — (%) | 45.12 | 42.15 | 74.43 | 36.43 |

**Supplementary Table 3.** Genome-wide significant (P<5×10E-8) and suggestive findings (P<10E-6) in the first and second analyses for the meta-analysis (Sheet 1: META-ANALYSES) and the respective single cohort analyses (Sheet 2: SINGLE ANALYSES).

SEE EXCEL TABLE

**Supplementary Table 4:** Linkage disequilibrium (LD) between variants from the first and second analysis and COVID-HGI release 5 variants.

SEE EXCEL TABLE

**Supplementary Table 5**: Replication analysis of the 13 COVID-19 HGI release 5 hits (PMID: 34237774).

SEE EXCEL TABLE

**Supplementary Table 6.** Genome-wide significant hits (P<5×10E-8) in the meta-analyses with HGI A2 and B2 summary statistics.

SEE EXCEL TABLE

**Supplementary Table 7.** FINEMAP results of the genome-wide significant and suggestive loci of the first analysis (Sheet 1: First analysis), second analysis (Sheet 2: Second analysis) and 19q13.33 (Sheet 3: 19q13.33).

SEE EXCEL TABLE

**Supplementary Table 8.** Candidate-based association statistics and stratified analyses of genome-wide variants from the first and second analyses, HGI variants, novel variant at 19q13.33 and the inversion at 17q21.31. Meta-analysis results and (Sheet 1: META-ANALYSES) and the respective single cohort analyses (Sheet 2: SINGLE ANALYSES) are shown.

SEE EXCEL TABLE

**Supplementary Table 9.** Association statistics of inversion allele H2. Meta-analysis results and (Sheet 1: META-ANALYSES) and the respective single cohort analyses (Sheet 2: SINGLE ANALYSES) are shown.

SEE EXCEL TABLE

**Supplementary Table 10.** Phenome-wide association study (PheWAS) of the 17q21.31 inversion.

SEE EXCEL TABLE

**Supplementary Table 11.** Lookup of eQTLs and sQTLs from GTex for the inversion (eQTL; Sheet 1: GTEx_eQTLs.chr17invers; sQTL; Sheet 2: GTEx_sQTLs.chr17invers) and rs1405655 (eQTL; Sheet 1: GTEx_eQTLs.rs1405655; sQTL; Sheet 2: GTEx_sQTLs.rs140565).

SEE EXCEL TABLE

**Supplementary Table 12:** Inversion effects on gene expression and splicing changes in monocytes across immune stimulations.

SEE EXCEL TABLE

**Supplementary Table 13:** Differential gene expression analysis of SARS-CoV-2 infected human brain organoids.

SEE EXCEL TABLE

**Supplementary Table 14:** Analysis results of Mendelian Randomization.

SEE EXCEL TABLE

**Supplementary Table 15:** Candidate-based association statistics and stratified analyses of the ABO blood types. Meta-analysis results and (Sheet 1: META-ANALYSES) and the respective single cohort analyses (Sheet 2: SINGLE ANALYSES) are shown.

SEE EXCEL TABLE

**Supplementary Table 16.** HLA association in COVID-19.

a)

| **Number of imputed 2-field alleles for the different HLA loci and cohorts.** In brackets we show numbers remaining after removing alleles with an allele frequency <0.05% and a marginal probability <0.3. | | | | | |
|---|---|---|---|---|---|
| | Germany | Italy | Norway | Spain | ALL |
| *A* | 29 (26) | 37 (31) | 21 (21) | 33 (28) | 39 (32) |
| *B* | 58 (45) | 68 (49) | 35 (35) | 68 (47) | 83 (54) |
| *C* | 25 (21) | 33 (25) | 20 (20) | 29 (23) | 36 (26) |
| *DPB1* | 21 (19) | 26 (19) | 18 (18) | 25 (20) | 33 (22) |
| *DQA1* | 15 (15) | 9 (8) | 13 (13) | 11 (8) | 19 (15) |
| *DQB1* | 17 (17) | 17 (16) | 17 (17) | 16 (15) | 19 (18) |

| | | | | | |
|---|---|---|---|---|---|
| *DRB1* | 38 (31) | 42 (36) | 27 (27) | 38 (32) | 47 (38) |
| *E* | - | 3 (2) | - | 3 (2) | 3 (2) |
| ALL | 203 (174) | 235 (186) | 151 (151) | 223 (175) | 279 (207) |

b)

| **Marginal Probability of HLA allele imputation.** The mean marginal probability(43) (measure for accuracy) and its standard deviation is shown for each locus and cohort. | | | | |
|---|---|---|---|---|
| | Germany | Italy | Norway | Spain |
| *A* | 0.948 ± 0.070 | 0.946 ± 0.084 | 0.962 ± 0.053 | 0.961 ± 0.060 |
| *B* | 0.888 ± 0.124 | 0.906 ± 0.104 | 0.921 ± 0.107 | 0.905 ± 0.105 |
| *C* | 0.970 ± 0.039 | 0.957 ± 0.075 | 0.940 ± 0.146 | 0.966 ± 0.049 |
| *DPB1* | 0.913 ± 0.103 | 0.979 ± 0.017 | 0.931 ± 0.082 | 0.9480 ± 0.092 |
| *DQA1* | 0.895 ± 0.124 | 0.985 ± 0.017 | 0.943 ± 0.070 | 0.988 ± 0.016 |
| *DQB1* | 0.932 0.120 | 0.965 ± 0.06 | 0.951 ± 0.087 | 0.982 ± 0.027 |
| *DRB1* | 0.839 ± 0.167 | 0.856 ± 0.142 | 0.857 ± 0.169 | 0.883 ± 0.119 |
| *E* | - | 0.995 ± 0.001 | - | 0.995 ± 0.001 |

c) HLA association statistics. Meta-analysis results and (Sheet 1: META-ANALYSES) and the respective single cohort analyses (Sheet 2: SINGLE ANALYSES) are shown.

SEE EXCEL TABLE

**Supplementary Table 17.** PepWAS analysis in COVID-19. The analysis results are split into multiple tabs for the overall analysis (unstratified = general) and the analysis of severity within each of the 4 cohorts.

SEE EXCEL TABLE

**Supplementary Table 18.** Analysis of HLA parameters in COVID-19. The analysis results are split into 5 tabs. CaseControl_Linear, CaseControl_Quadratic, Severity_Linear, Severity_Quadratic and CorrelationAcrossAlleles. The analyses behind these tabs are described in detail in the **Supplementary Methods section** of the HLA analyses.

SEE EXCEL TABLE

**Supplementary Table 19.** Detailed statistics of main and stratified analysis of the first and second as well as mortality analyses for the Y-chromosome haplotypes. Meta-analysis results and (Sheet 1: META-ANALYSES) and the respective single cohort analyses (Sheet 2: SINGLE ANALYSES) are shown.

SEE EXCEL TABLE

**Supplementary Table 20.** Ensembl v102 annotations of protein-coding candidate genes of 17q21.31 and 19q13.33 loci.

SEE EXCEL TABLE
